# Cross-Phenotype GWAS Supports Shared Genetic Susceptibility to Systemic Sclerosis and Primary Biliary Cholangitis

**DOI:** 10.1101/2024.07.01.24309721

**Authors:** Yiming Luo, Atlas Khan, Lili Liu, Cue Hyunkyu Lee, Gabriel J Perreault, Sydney F Pomenti, Pravitt Gourh, Krzysztof Kiryluk, Elana J Bernstein

## Abstract

**Objective:** An increased risk of primary biliary cholangitis (PBC) has been reported in patients with systemic sclerosis (SSc). Our study aims to investigate the shared genetic susceptibility between the two disorders and to define candidate causal genes using cross-phenotype GWAS meta-analysis.

**Methods:** We performed cross-phenotype GWAS meta-analysis and colocalization analysis for SSc and PBC. We performed both genome-wide and locus-based analysis, including tissue and pathway enrichment analyses, fine-mapping, colocalization analyses with expression quantitative trait loci (eQTL) and protein quantitative trait loci (pQTL) datasets, and phenome-wide association studies (PheWAS). Finally, we used an integrative approach to prioritize candidate causal genes from the novel loci.

**Results:** We detected a strong genetic correlation between SSc and PBC (rg = 0.84, p = 1.7 × 10^-6^). In the cross-phenotype GWAS meta-analysis, we identified 44 non-HLA loci that reached genome-wide significance (p < 5 × 10^-8^). Evidence of shared causal variants between SSc and PBC was found for nine loci, five of which were novel. Integrating multiple sources of evidence, we prioritized *CD40, ERAP1, PLD4, SPPL3*, and *CCDC113* as novel candidate causal genes. The *CD40* risk locus colocalized with trans-pQTLs of multiple plasma proteins involved in B cell function.

**Conclusion:** Our study supports a strong shared genetic susceptibility between SSc and PBC. Through cross-phenotype analyses, we have prioritized several novel candidate causal genes and pathways for these disorders.

**Key messages:** 

**What is already known on this topic:**

- Systemic sclerosis (SSc) and primary biliary cholangitis (PBC) are autoimmune disorders that exhibit overlapping clinical and histological features.
- The prevalence of PBC is higher in patients with SSc compared to the general population.
- Multiple susceptibility genomic loci have been identified for SSc and PBC through genome-wide association studies (GWAS).

**What this study adds:**

- There is a strong genetic correlation between SSc and PBC, comparable in magnitude to the genetic correlation between SSc and systemic lupus erythematosus (SLE). This shared genetic susceptibility aligns with the observed increased relative risk of developing PBC and SLE in individuals with SSc.
- Using cross-phenotype GWAS and colocalization analysis, we have discovered nine genomic loci that account for the shared genetic etiology. Five of the nine loci were novel.
- Using an integrative approach, we have prioritized five novel candidate causal genes: *CD40, ERAP1, PLD4, SPPL3* and *CCDC113*.
- The *CD40* risk allele for SSc and PBC is paradoxically associated with reduced CD40 levels. Causal inference analyses indicate that this reduction in CD40 levels, due to *CD40* locus polymorphism, leads to an increase in various plasma proteins involved in B cell activation, including the CD40 ligand.

**How this study might affect research, practice or policy:**

- Mechanistic studies are needed to confirm the candidate causal genes prioritized by our *in silico* analyses.
- Our study advocates for heightened awareness among rheumatologists regarding the possibility of concurrent PBC in patients with SSc.

## Introduction

Systemic sclerosis (SSc) is a multi-system autoimmune disease characterized by a complex interplay of fibrosis, vasculopathy, and inflammation. Unlike in other systemic autoimmune rheumatic diseases, the therapeutic response to immunosuppressive medications in SSc is organ-dependent. Certain organ involvement, including gastrointestinal tract fibrosis, has not been found to be responsive to immunosuppressive therapy.^1^

Primary biliary cholangitis (PBC) is an autoimmune liver disease characterized by inflammation of the intrahepatic bile ducts leading to liver fibrosis. Similar to the gastrointestinal involvement of SSc, the efficacy of immunosuppressive therapies in PBC has not yet been established.^2^ The prevalence of PBC in patients with SSc is 2-2.5%, substantially higher than its prevalence of 0.4% in the general population.^3,4^ Thus, there is likely an overlap of etiopathogenesis between SSc and PBC.

Genome-wide association studies (GWAS) have been conducted in both SSc and PBC, identifying numerous genomic loci associated with these two disorders.^5,6^ Cross-phenotype GWAS analytic approaches that leverage existing GWAS summary statistics have emerged as a powerful new strategy for identifying shared mechanisms and novel risk loci.^7^ We used this approach to systematically assess overlapping susceptibility and identify novel candidate causal genes that contribute to the common etiopathogenesis of the two disorders.

## Methods

### Study Design

An overview of the study design is shown in **Figure 1**.

**Figure 1.**
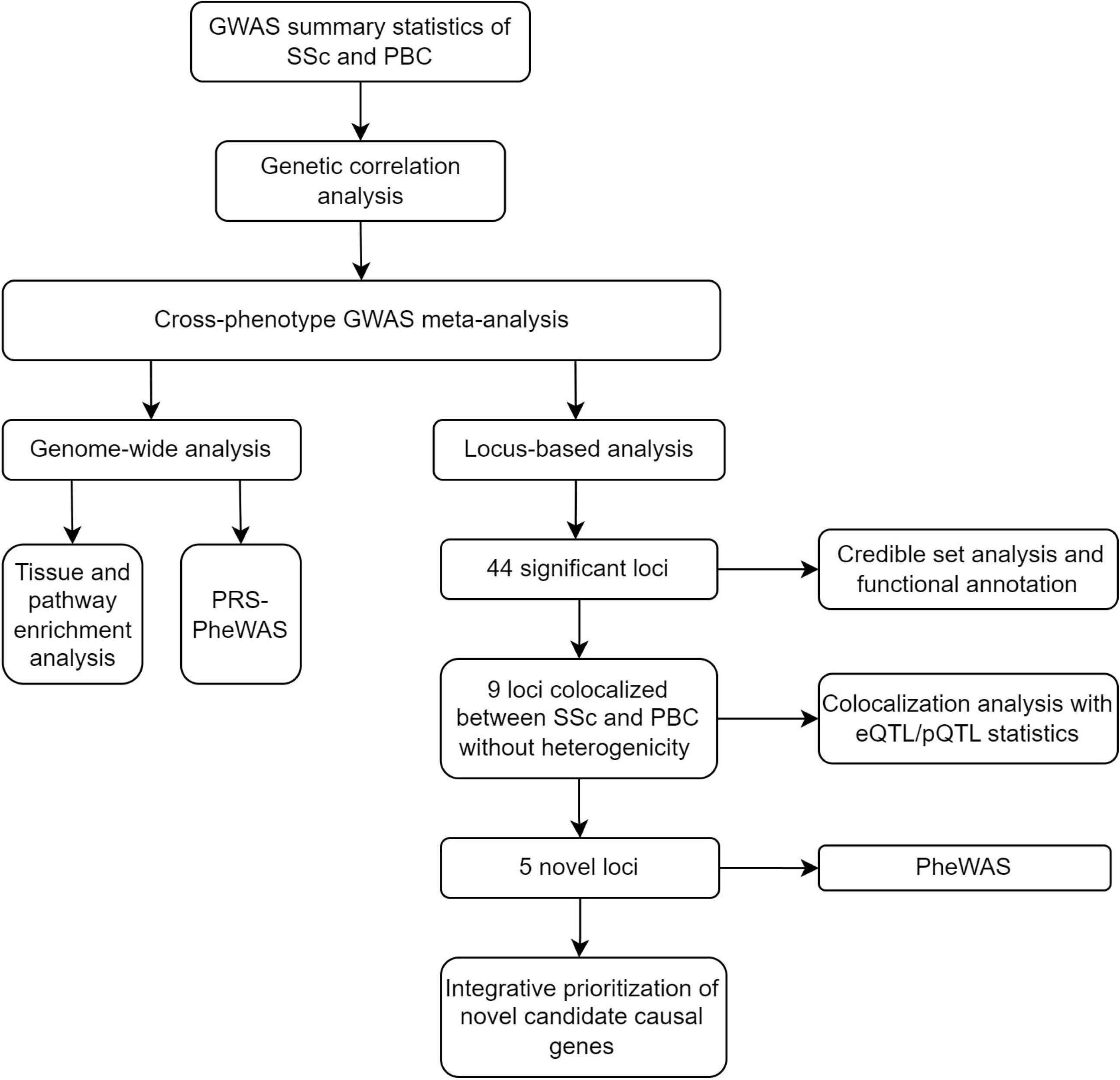
Flowchart for the overview of the study. We performed multiple cross-phenotype GWAS analyses to identify the shared genetic susceptibility between SSc and PBC. Additionally, we also performed a single-center retrospective chart review to evaluate the prevalence of PBC, including potentially undiagnosed cases, in patients with SSc. SSc: systemic sclerosis. PBC: primary biliary cholangitis. GWAS: genome-wide association studies. PLEIO: Pleiotropic Locus Exploration and Interpretation using Optimal test. eQTL: expression quantitative trait loci. PheWAS: phenome-wide association studies.

### GWAS Summary Statistics

We obtained summary statistics of SSc and PBC from their recent GWAS meta-analyses. The GWAS for SSc was comprised of 26,679 individuals (9,095 cases and 17,584 controls),^5^ while the GWAS for PBC was comprised of 24,510 individuals (8,021 cases and 16,489 controls).^6^ For comparison, we also obtained GWAS summary statistics for rheumatoid arthritis (RA)^8^ and systemic lupus erythematosus (SLE)^8,9^ since both are prevalent in patients with SSc. We also obtained GWAS summary statistics from expression quantitative trait locus (eQTL) datasets, including the eQTLGen, Genotype-Tissue Expression (GTEx, for skin, liver, and lung) and Correlated Expression and Disease Association Research (CEDAR), as well as plasma proteomics quantitative trait locus (pQTL) datasets from the UK Biobank.^10-13^ The GWAS summary statistics were harmonized using reference data from the 1000 Genome Project (phase 3) and underwent quality control with MungeSumstats.^14,15^ The included GWAS datasets are summarized in **Table 1**.

**Table 1.**
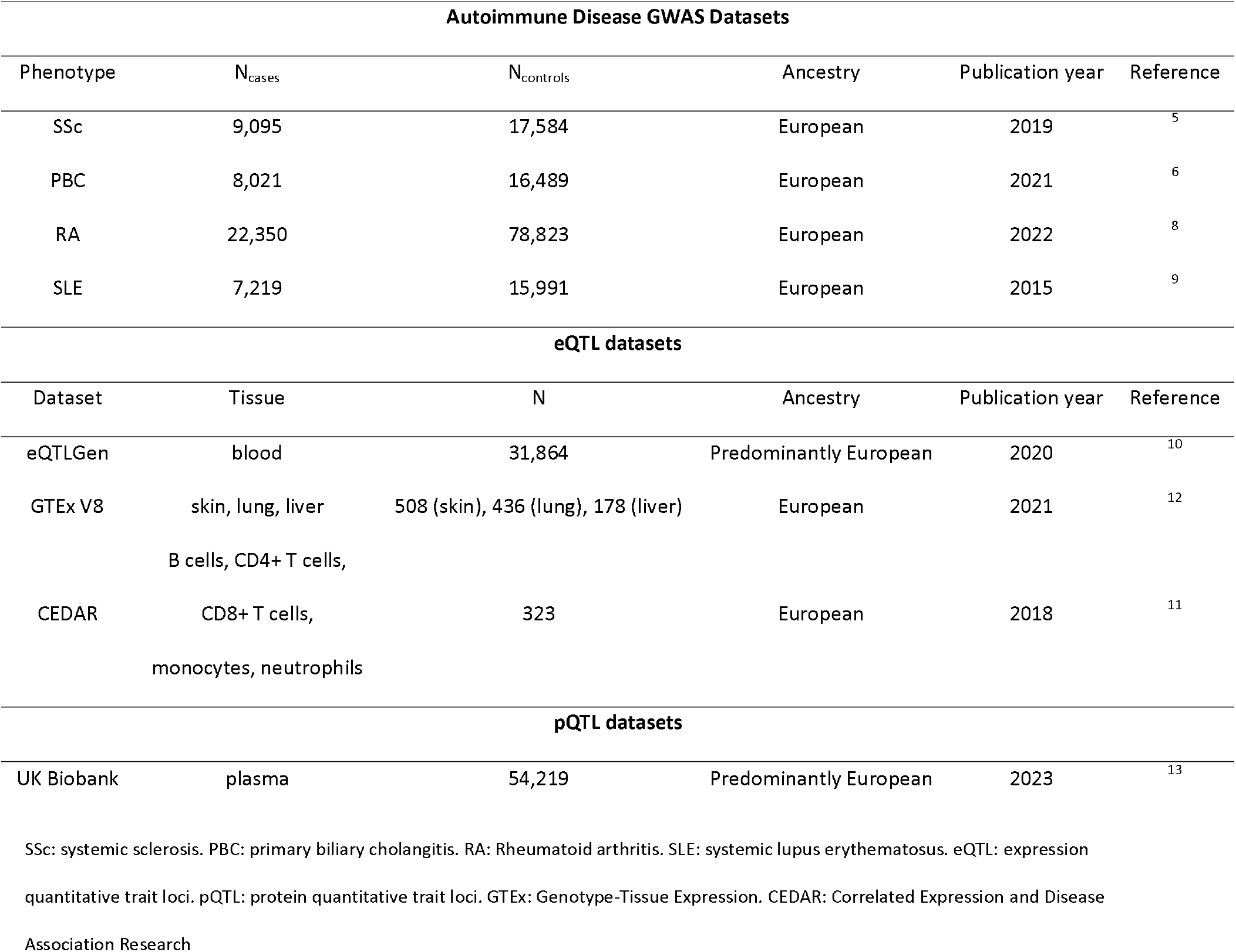
GWAS summary statistics included in this study.

### Genetic Correlation Analysis

To quantify the degree of shared genetic susceptibility, we used linkage disequilibrium score regression (LDSC) to estimate the global genetic correlation (r_g_), excluding the human leukocyte antigens (HLA) region, between each phenotype pair (SSc, PBC, RA, and SLE).^16,17^ A Bonferroni-corrected p-value of 8.3 × 10^-3^ was used as the significance threshold.

### Cross-Phenotype GWAS Meta-Analysis

We performed a cross-phenotype GWAS meta-analysis to identify pleiotropic loci shared between SSc and PBC. We combined the summary statistics of SSc and PBC using the fixed-effect model with effect size estimates and standard errors using METAL.^18^ Genomic control correction was applied to the summary statistics of each phenotype before the meta-analysis.^19^ After the meta-analysis, we excluded SNPs in the HLA region or with evidence of heterogeneity (heterogeneity p < 0.05). We used wANNOVAR to annotate the lead SNPs of the significant loci.^20^ We defined novel loci as those that were significant in the cross-phenotype meta-analysis but not significant in either SSc or PBC input GWAS. The loci were named based on the annotation of the lead SNPs from wANNOVAR, which relies on the distance to nearby genes.^20^

The fixed-effect model is limited in examining SNPs with heterogeneity of effects. Therefore, we performed a sensitivity analysis using the PLEIO (Pleiotropic Locus Exploration and Interpretation using Optimal test) method.^21^ PLEIO is designed for cross-phenotype meta-analysis and can account for heritability, genetic correlation, and sample overlap. There is no established method for extrapolating Z-scores from PLEIO statistics. Therefore, we used the fixed-effect model statistics for subsequent analyses.

### Fine-Mapping and Credible Set Analyses

We prioritized the most likely causal SNPs from the cross-phenotype meta-analysis statistics by calculating the 99% credible sets using CARMA (CAusal Robust Mapping method with Annotations).^22^ These sets represent the smallest sets of SNPs with the probability of including the causal variant exceeding 99%. CARMA is a novel Bayesian model for fine-mapping that can better account for the uneven measurement of SNPs in each GWAS study of a meta-analysis, as well as the discrepancies between summary statistics and LD from external reference panels. We incorporated functional annotation into CARMA using the prior causal probabilities based on the meta-analysis of 15 UK Biobank traits from PolyFun (POLYgenic FUNctionally-informed fine-mapping).^23^

### Tissue and Pathway Enrichment Analyses

We used two methods to prioritize the pathways and tissues that contribute to the pleiotropy of SSc and PBC: MAGMA (Multi-marker Analysis of GenoMic Annotation) and DEPICT (Data-driven Expression Prioritized Integration for Complex Traits).^24-26^ MAGMA and DEPICT perform enrichment analyses at pathway and tissue levels but use different approaches to associate loci with genes. MAGMA annotates SNPs based on their locations relative to genic regions (transcription start and stop sites +/-10 kb window). DEPICT prioritizes genes in a locus if genes in different loci have similar predicted functions. We reported the Bonferroni-corrected p values.

### Colocalization between SSc and PBC

We performed colocalization analyses between SSc and PBC in loci that are significant in the cross-phenotype meta-analysis with the fixed-effect model. Colocalization analyses infer the probability that a single genetic variant is causal to both traits of interest – SSc and PBC. We performed colocalization using the Wakefield’s method from the R package “coloc”.^27^ For loci with independent signals in the conditional analysis (p < 5 × 10^-8^), we performed additional colocalization analyses using statistics from the conditional analyses. Evidence of colocalization was defined as at least one signal with a colocalization probability (PP4) above 70%.

### Colocalization between Meta-Analysis Statistics and eQTL/pQTL Datasets

We prioritized loci that met the following criteria for additional colocalization analyses: (1) lead SNPs without evidence of heterogeneity (P_het_ ≥ 0.05); (2) significant in the fixed-effect cross-phenotype meta-analysis; and (3) colocalized between SSc and PBC. We performed colocalization analyses between the SSc-PBC meta-analysis statistics of these loci and cis-eQTL and cis-pQTL datasets to predict transcripts or proteins associated with the genomic signal. For the eQTL colocalization analyses, we included relevant tissues and cells, including blood, skin, lung, liver, and immune cells. We first screened genes using SNPs in that locus’s 99% credible set to query the eQTL databases. Next, we selected genes with significant eQTL signals for the colocalization analyses. For loci that colocalized with a cis-pQTL signal, we performed additional colocalization analyses between the meta-analysis statistics and trans-pQTLs measured at that locus to investigate the downstream effects of the candidate causal gene. A colocalization probability (PP4) above 70% was used as the significance threshold, and 50% as the suggestive threshold.

### Phenome-wide association studies (PheWAS)

We performed PheWAS for the lead SNP for each novel locus that colocalized between SSc and PBC. The PheWAS was performed within three biobanks: the Electronic Medical Records and Genomics III (eMERGE-III), All of Us, and the UK Biobank. Meta-PheWAS statistics were then calculated by fixed-effect meta-analysis of PheWAS results across the three biobanks.^28-30^ The details of the genotyping methods, imputation, quality control, ancestry inference, covariate adjustment, and phenotype identification were described in our previous studies.^31-33^

To further explore pleiotropic associations, we performed meta-PheWAS analyses on the polygenic risk scores (PRS) of the cross-phenotype meta-analysis (PRS-meta-PheWAS). We used PRS-CS (continuous shrinkage), a method based on high-dimensional Bayesian regression, to generate the weights for PRS.^34^ The HLA region was excluded. We set the Bonferroni-corrected statistical significance threshold for phenome-wide significance at 2.75x10^-5^ (0.05/1,817 phecodes tested). Lastly, we manually queried the top SNPs using PheWAS results from the Open Targets Genetics webpage^35,36^, which includes data from the GWAS Catalogue, UK Biobank, and FinnGen. We designated the effect allele as the GWAS risk allele in SSc-PBC cross-phenotype analysis.

### Integrative prioritization of novel candidate causal genes

For each novel locus, we prioritized candidate causal genes using an integrative approach. Each candidate causal gene was scored with the following criteria and we then calculated the number of the satisfied criteria (“priority score”): (1) genes most proximal to the lead SNP at the locus; (2) genes colocalizing with the locus in the examined cis-eQTL datasets; (3) genes colocalizing with the locus in the examined cis-pQTL datasets; (4) genes with a nonsynonymous coding variant in the credible set; (5) Genes prioritized by MAGMA (false discovery rate [FDR] q-value < 0.05)^24^; (6) genes prioritized by DEPICT (FDR q-value < 0.05)^26^; (7) genes receiving the top score from the Variant-to-Gene (V2G) pipeline on the Open Targets Genetics webpage^35,36^, using the fine-mapped SNPs (PIP > 10%); (8) genes whose predicted regulatory elements from the ENCODE-rE2G model^37^ intersected with the fine-mapped SNPs (PIP > 10%); (9) genes prioritized by the large language model (LLM) GPT-4, which was recently suggested as a systematic way to mine literature for candidate causal gene prioritization (input described in the **Supplemental Note**).^38^ We prioritized the genes with the highest priority scores within each locus. Patients or the public were not involved in the design, or conduct, or reporting, or dissemination plans of our research.

## Results

### Global Genetic Correlation

There was a strong global genetic correlation between SSc and PBC (rg = 0.84, p = 1.7 × 10^-5^), in which the effect estimate is comparable to the genetic correlation between SSc and SLE (rg = 0.84, p = 1.6 × 10^-15^). The pairwise comparison of global genetic correlation in SSc, PBC, RA and SLE is shown in Figure 2.

**Figure 2.**
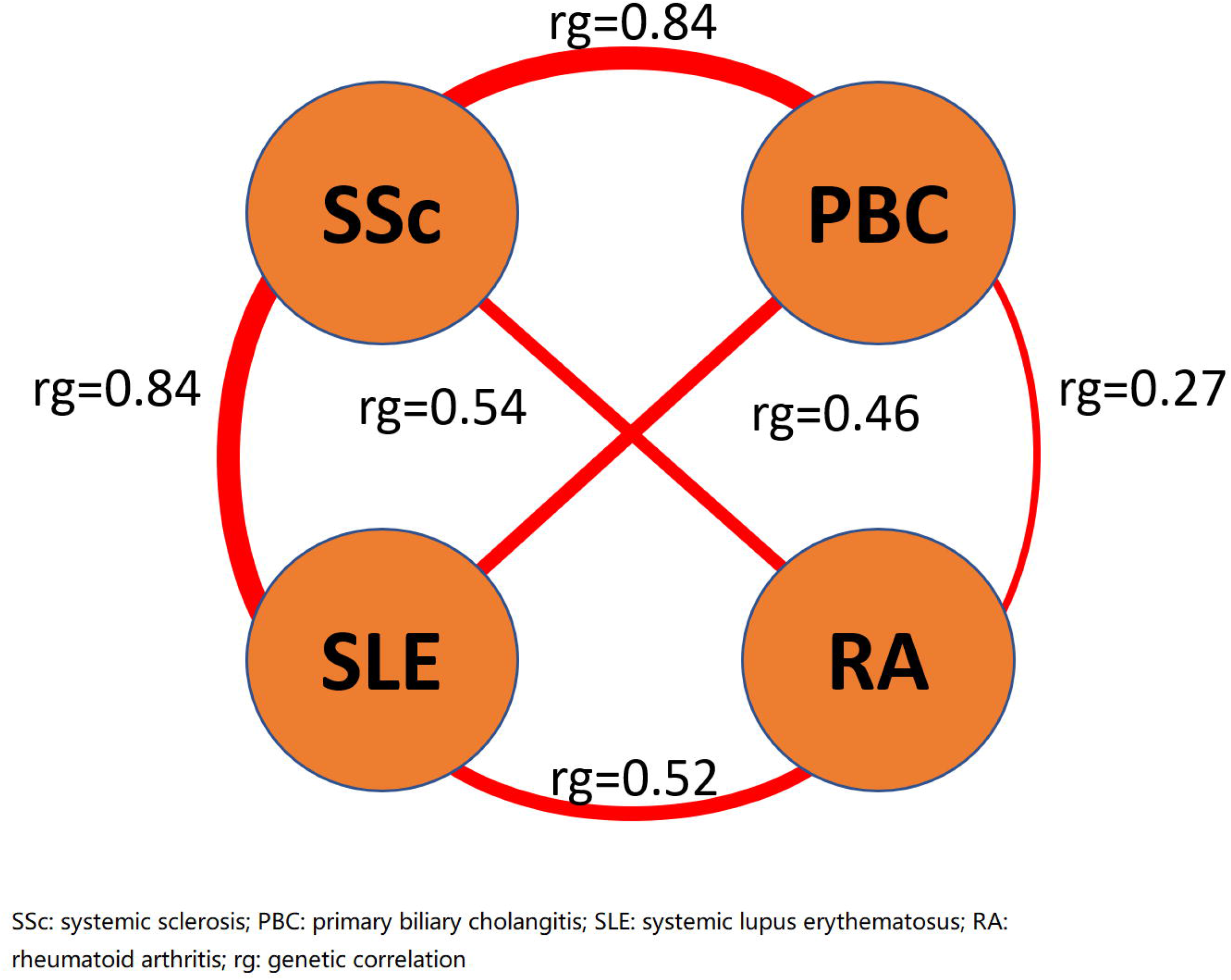
Pairwise genetic correlation in SSc, PBC, RA and SLE. We performed genetic correlation analyses using linkage disequilibrium score regression pairwise among SSc, PBC, RA and SLE. The effect estimates of genetic correlation between SSc and PBC, and between SSc and SLE, were both 0.84, higher than other pairwise estimates.

### Cross-Phenotype GWAS Meta-Analysis

We performed a cross-phenotype meta-analysis for SSc and PBC using the fixed-effect model. The Manhattan plot is shown in Figure 3. There were 44 non-HLA loci that reached genome-wide significance (p < 5 × 10^-8^, Supplemental Table 1). The genomic inflation factor (λ) was 1.065 and the LDSC intercept was 1.010 (standard error 0.012). In 16 out of the 44 significant loci (36%), there was evidence of heterogeneity in the lead SNPs (Phet < 0.05). However, these loci remained significant after removing SNPs with heterogeneity.

**Figure 3.**
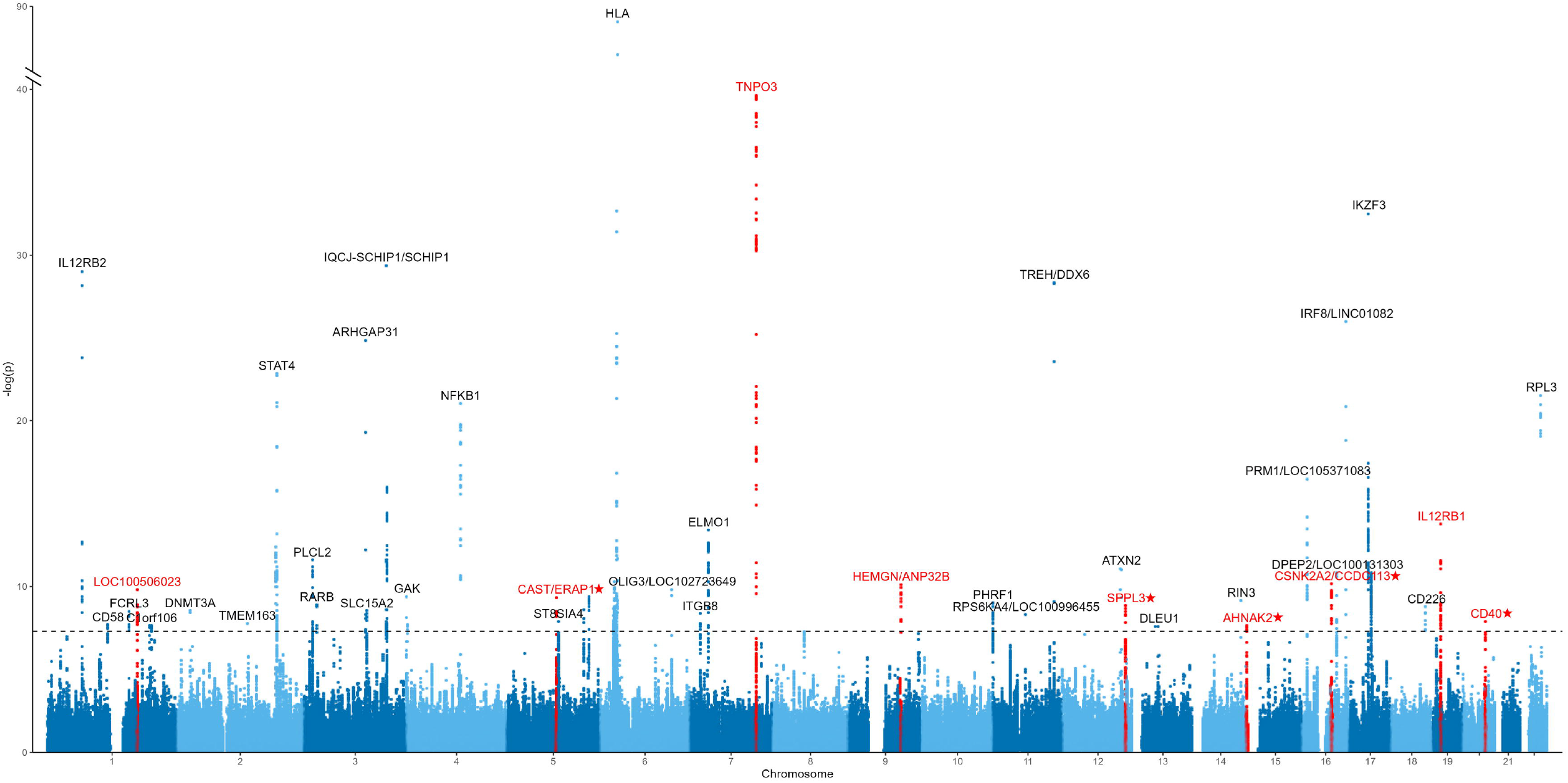
Manhattan plot of the cross-phenotype GWAS meta-analysis in SSc and PBC using the fixed-effect model. We performed a cross-phenotype GWAS meta-analysis in SSc and PBC using the fixed-effect model. We identified 44 significant genomic loci (p < 5 × 10^-8^). We found 9 loci that colocalized between SSc and PBC and did not show evidence of heterogeneity (indicated in red), among which five were novel (indicated with an asterisk). SNPs with evidence of heterogeneity (P_het_ < 0.05) were excluded.

Given the high proportion of loci with evidence of heterogeneity, we performed another cross-phenotype meta-analysis between SSc and PBC using PELIO as a sensitivity analysis. There were 58 non-HLA loci that reached genome-wide significance (p < 5 × 10^-8^, Supplemental Table 2). The genomic inflation factor (λ) was 1.091. Forty-one of the 44 loci (93%) that were significant in the fixed-effect model were also significant in PLEIO. Regarding the novel loci, five out of the seven (71%) identified in the fixed-effect model were also significant in PLEIO. The two novel loci that were only significant in the fixed-effect model, *CD40* and *AHNAK2*, had p-values of 7.37 × 10^-8^ and 3.37 × 10^-7^ in PLEIO. Two novel loci, NDFIP1 and PPHLN1, were significant only in PLEIO but not in the fixed-effect model. However, the evidence for the association of these loci with SSc was insufficient (lead SNP p values 3.93 × 10^-2^ and 1.33 × 10^-2^, respectively). Thus, no further analyses were performed for these loci.

### Tissue and Pathway Enrichment Analyses

We used two data-driven genome-wide methods, MAGMA and DEPICT, to explore pathway and tissue enrichment across SSc and PBC. Tissue enrichment analysis using MAGMA prioritized spleen (p = 1.21 × 10^-6^), whole blood (p = 1.59 × 10^-6^), Epstein-Barr Virus (EBV)-transformed lymphocytes (p = 4.16 × 10^-4^), lung (p = 2.81 × 10^-3^), and terminal ileum (p = 4.63 × 10^-3^) (**Figure 4a, Supplemental Table 3**).

**Figure 4.**
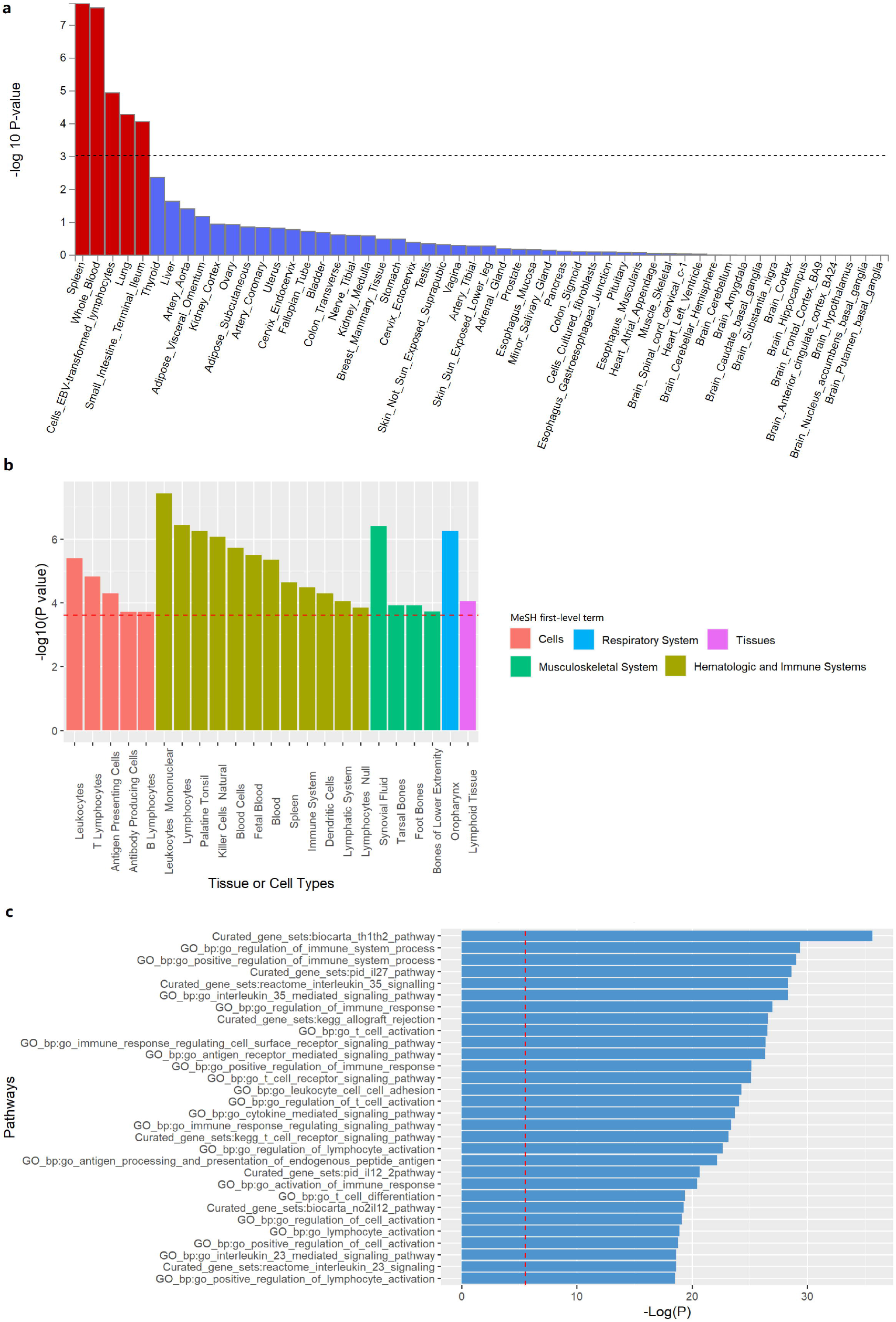
Tissue and pathway enrichment analyses. **a** Tissue enrichment analysis using MAGMA prioritized tissues related to the immune system (spleen, whole blood and EBV-transformed lymphocytes), respiratory system (lung) and digestive system (terminal ileum). **b** Tissue enrichment analysis using DEPICT prioritized multiple tissues and cells related to the immune system, respiratory system and musculoskeletal system. c Significant enrichment of multiple immune-related pathways associated with SSc and PBC using MAGMA.

Concurrently, the DEPICT method significantly enriched 20 tissues and cells, most related to the immune system or respiratory tract. The top-ranked tissues/cells were mononuclear leukocytes (p = 2.95 × 10^-5^), oropharynx (p = 3.80 × 10^-5^), palatine tonsil (p = 3.80 × 10^-5^), and synovial fluid (p = 2.63 × 10^-4^) (**Figure 4b, Supplemental Table 4**).

Pathway enrichment analysis using MAGMA revealed significant enrichment in 73 gene sets (Figure 4c, Supplemental Table 6). The top-ranked gene sets were “Th1/Th2 pathway” (p = 4.92 × 10^-12^), “positive regulation of immune system process” (p = 3.67 × 10^-9^), “IL-27 pathway” (p = 5.60 × 10^-9^) and “IL-35 pathway” (p = 7.63 × 10^-9^). A separate pathway enrichment analysis using DEPICT revealed significant enrichment in 263 gene sets, which similarly involved multiple aspects of the human immune system (**Supplemental Data 1**).

### Fine-Mapping and Credible Set Analyses

To infer the causal SNPs underlying the association signals, we performed fine-mapping and generated 99% credible sets. There were 53 predicted causal signals across the 44 significant loci. Eleven (21%) of the credible sets contained only one SNP. Twenty-six (49%) credible sets contained fewer than five SNPs. Thirty-nine (74%) credible sets contained fewer than ten SNPs. In 30 (68%) loci, the maximum posterior inclusion probability (PIP) in their credible sets contained the lead SNPs of that loci. There were no nonsynonymous coding variants in any of the credible sets. The credible SNP sets in each locus are summarized in **Supplemental Table 5**.

### Colocalization between SSc and PBC

Due to LD, significant SNPs within genomic loci may not necessarily be causal for the associated trait. Therefore, we conducted colocalization analyses to determine whether there was at least one shared causal variant between SSc and PBC in loci significant in the cross-phenotype meta-analysis. We identified 9 loci that colocalized between SSc and PBC (PP4 > 70%) and did not have evidence of heterogeneity (P_het_ ≥ 0.05), as detailed in the **Table 2** and **Supplemental Figure 1**. Notably, five were novel: *CSNK2A2*/*CCDC113, SPPL3, CAST*/*ERAP1, AHNAK2*, and *CD40*.

**Table 2.** Genomic loci significant in the cross-phenotype GWAS, colocalized between SSc and PBC and do not have evidence of heterogeneity.

| Locus | Lead SNP | CHR | BP | A1 | A2 | FE OR | FE p-values | PLEIO p-values | Colocalization PP4 | Novel loci |
| --- | --- | --- | --- | --- | --- | --- | --- | --- | --- | --- |
| *TNPO3* | rs17338998 | 7 | 128618559 | T | C | 1.52 | 2.37E-40 | 4.16E-73 | 98% | No |
| *CSNK2A2/CCDC113* | rs2731783 | 16 | 58253460 | A | G | 1.15 | 7.79E-11 | 1.50E-09 | 96% | Yes |
| *IL12RB1* | rs2305743 | 19 | 18193191 | A | G | 0.86 | 1.95E-14 | 7.42E-14 | 96% | No |
| *LOC100506023* | rs2022449 | 1 | 173238736 | T | G | 1.12 | 1.69E-10 | 2.82E-09 | 92% | No |
| *SPPL3* | rs551125 | 12 | 121203427 | C | T | 0.91 | 1.65E-09 | 2.59E-08 | 91% | Yes |
| *CAST/ERAP1* | rs27524 | 5 | 96101944 | A | G | 0.9 | 4.89E-10 | 6.04E-09 | 85% | Yes |
| *HEMGN/ANP32B* | rs4743150 | 9 | 100740124 | T | C | 0.88 | 8.29E-11 | 2.80E-10 | 85% | No |
| *AHNAK2* | rs10083496 | 14 | 105402786 | G | A | 1.09 | 2.56E-08 | 7.37E-08 | 80% | Yes |
| *CD40* | rs4810485 | 20 | 44747947 | T | G | 1.13 | 1.44E-08 | 7.37E-08 | 73% | Yes |

SSc: systemic sclerosis. PBC: primary biliary cholangitis. PLEIO: Pleiotropic Locus Exploration and Interpretation using Optimal test. FE: fixed-effect model. PP4: posterior probability of hypothesis 4 that there is one variant causal to both traits. A1: effect allele. A2: non-effect allele.

### Colocalization with Tissue- and Immune Cell-Specific cis-eQTL

In the nine loci that were significant in cross-phenotype meta-analysis without evidence of heterogeneity and colocalized with SSc and PBC, we further performed colocalization between SSc-PBC meta-analysis statistics and eQTL statistics in blood, skin, lung, liver and immune cells. In seven of the nine loci, the SSc-PBC meta-analysis statistics colocalized with at least one transcript in the examined eQTL statistics (PP4 > 50%), prioritizing candidate causal genes at these loci (Figure 5). The eQTL colocalized transcripts were *IRF5, TNPO3, ANP32B, IL12RB1, ERAP1, ERAP2, SPPL3, AKT1, PLD4, LINC00638* and *CD40*.

**Figure 5.**
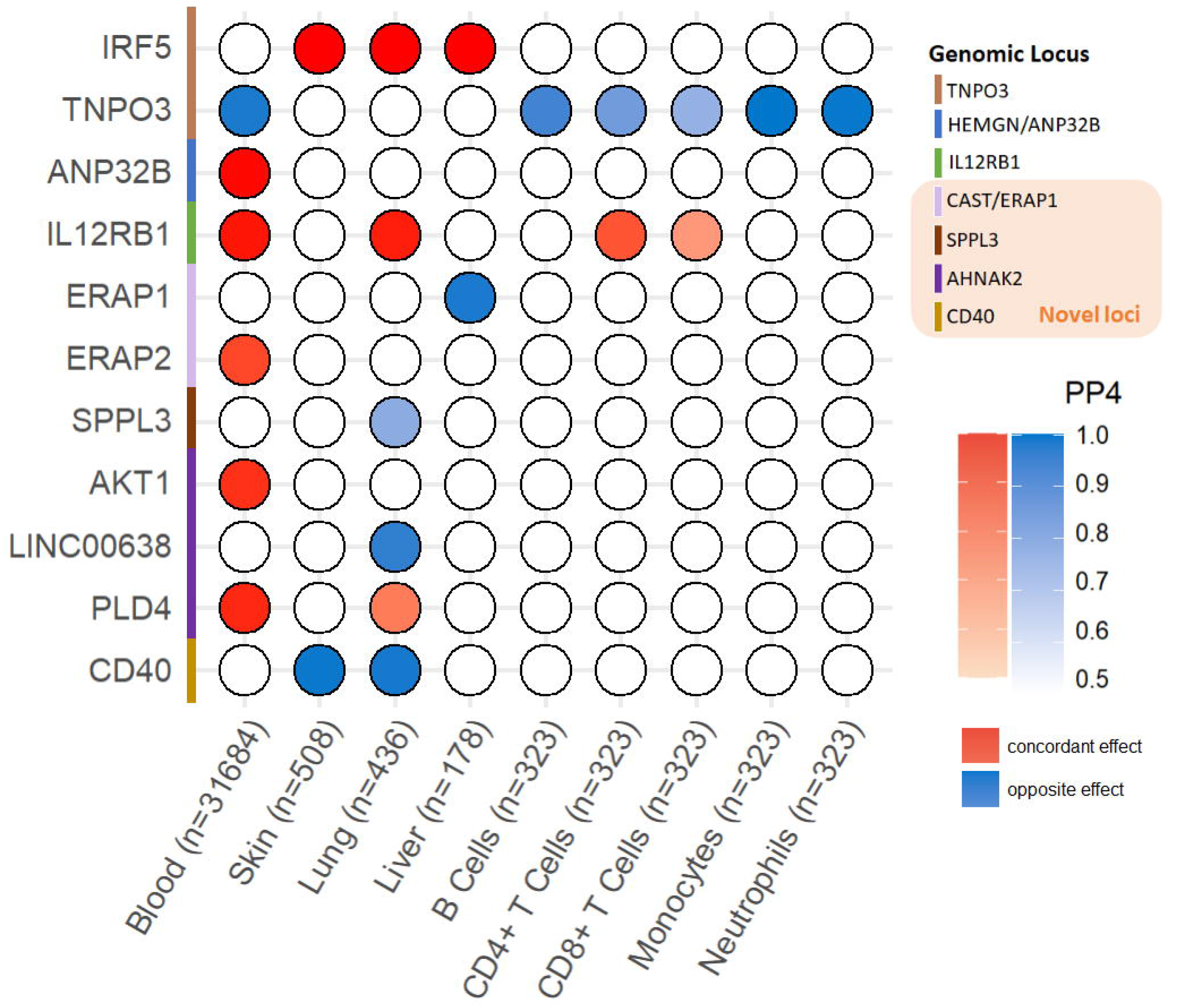
Colocalization analysis with expression quantitative trait loci (eQTL) in blood, skin, lung, liver and immune cells. Out of the nine loci that were significant in the fixed-model cross-phenotype meta-analysis, showed no evidence of heterogeneity, and colocalized between SSc and PBC, seven colocalized with expressed genes in at least one of the examined tissues.

### Colocalization with plasma cis- and trans-pQTL

Across the nine loci mentioned above, only the IL12RB1 and CD40 loci encode secreted proteins previously assessed in blood pQTL datasets. These plasma proteins include interleukin 12 receptor subunit beta 1 (IL12RB1) and CD40, respectively. The SSc-PBC meta-analysis statistics colocalized with cis-pQTL for CD40 protein levels (PP4 = 98%) but not with IL12RB1 protein levels (PP4 = 0.4%). The SSc-PBC risk allele was associated with lower plasma CD40 levels. Moreover, the SSc-PBC risk alleles at the CD40 locus colocalized in trans to reduced *BAFF* levels (PP4 = 99%) and increased levels of *CD40L* (PP4 = 99%), *FCER2* (PP4 = 99%), *CD22* (PP4 = 99%), *TRAF2* (PP4 = 97%), *FCLR1* (PP4 = 99%), and *TCL1A* (PP4 = 99%).

### PheWAS

We performed a meta-PheWAS for the five novel loci that colocalized between SSc and PBC. Three novel loci had significant associations with at least one phecode (Supplemental Figure 3). Rs10083496-G (ANNAK2 locus) was associated with “Systemic lupus erythematosus” (OR = 1.12, p = 2.05 × 10^-7^). Rs4810485-T (*CD40* locus) was associated with “Non-Hodgkins lymphoma” (OR = 1.11, p = 1.38 × 10^-6^) and “Anxiety disorders” (OR = 1.04, p = 2.86 × 10^-6^).

We subsequently performed an additional meta-PheWAS analysis to evaluate phenotypic associations of a genome-wide PRS based on the SSc-PBC meta-analysis statistics, excluding the HLA region (“SSc-PBC PRS”). We identified a total of 134 significant associations, the majority of which were related to immune dysregulation. As expected, SSc (OR = 1.78, p = 9.19 × 10^-30^) and PBC (OR = 3.01, p = 5.55 × 10^-86^) were top-ranked in their effect estimates associated with the SSc-PBC PRS, confirming that the PRS captures the risk for both diseases in the external datasets. Moreover, the PRS was associated with systemic autoimmune rheumatic diseases including “Rheumatoid arthritis” (OR = 1.24, p = 2.16 × 10^-55^), “Systemic lupus erythematosus” (OR = 1.75, p = 3.34 × 10^-64^), “Sicca syndrome” (OR = 1.58, p = 9.46 × 10^-43^), “Polyarteritis nodosa and allied conditions” (OR = 1.18, p = 6.72 × 10^-10^), as well as with non-rheumatic autoimmune diseases including “Hypothyroidism” (OR = 1.12, p = 4.42 × 10^-55^), “Multiple sclerosis” (OR = 1.30, p = 1.15 × 10^-23^), “Inflammatory bowel disease and other gastroenteritis and colitis” (OR = 1.14, p = 3.23 × 10^-21^) and “Idiopathic fibrosing alveolitis” (OR = 1.19, p = 2.00 × 10^-8^) (**Supplemental Figure 5 and Supplemental Table 8**).

Additionally, we manually queried the top SNP from the novel loci using PheWAS results from the Open Targets Genetics webpage. Rs4810485-T (CD40 locus) was associated with multiple autoimmune disorders, including “rheumatoid arthritis” (OR = 0.85, p = 5.7 × 10^-9^), “inflammatory bowel disease” (OR = 1.08, p = 4.6 × 10^-10^) and “multiple sclerosis” (OR = 1.08, p = 1.8 × 10^-5^). Rs27524-G (CAST/ERAP1 locus) was associated with “ankylosing spondylitis” (OR = 0.84, p = 5.4 × 10^-7^), “iridocyclitis” (OR = 0.88, p = 6.9 × 10^-8^) and “psoriasis” (OR = 0.85, p = 1.6 × 10^-6^).

### Integrative prioritization of novel candidate causal genes

We prioritized five candidate causal genes from the novel loci using a scoring approach that integrates nine *in silico* annotation methods (**Figure 6**. MAGMA, DEPICT and ENCODE-rE2G results in the **Supplemental Table 9-11**. GPT-4 output described in the **Supplemental Note**). CD40 received the highest priority score of 8, meeting all criteria except for credible sets containing a non-synonymous coding variant. *ERAP1* received a priority score of 6, followed by *PLD4* with a score of 5. *SPPL3* and *CCDC113* received priority scores of 4 and 3, respectively.

**Figure 6.**
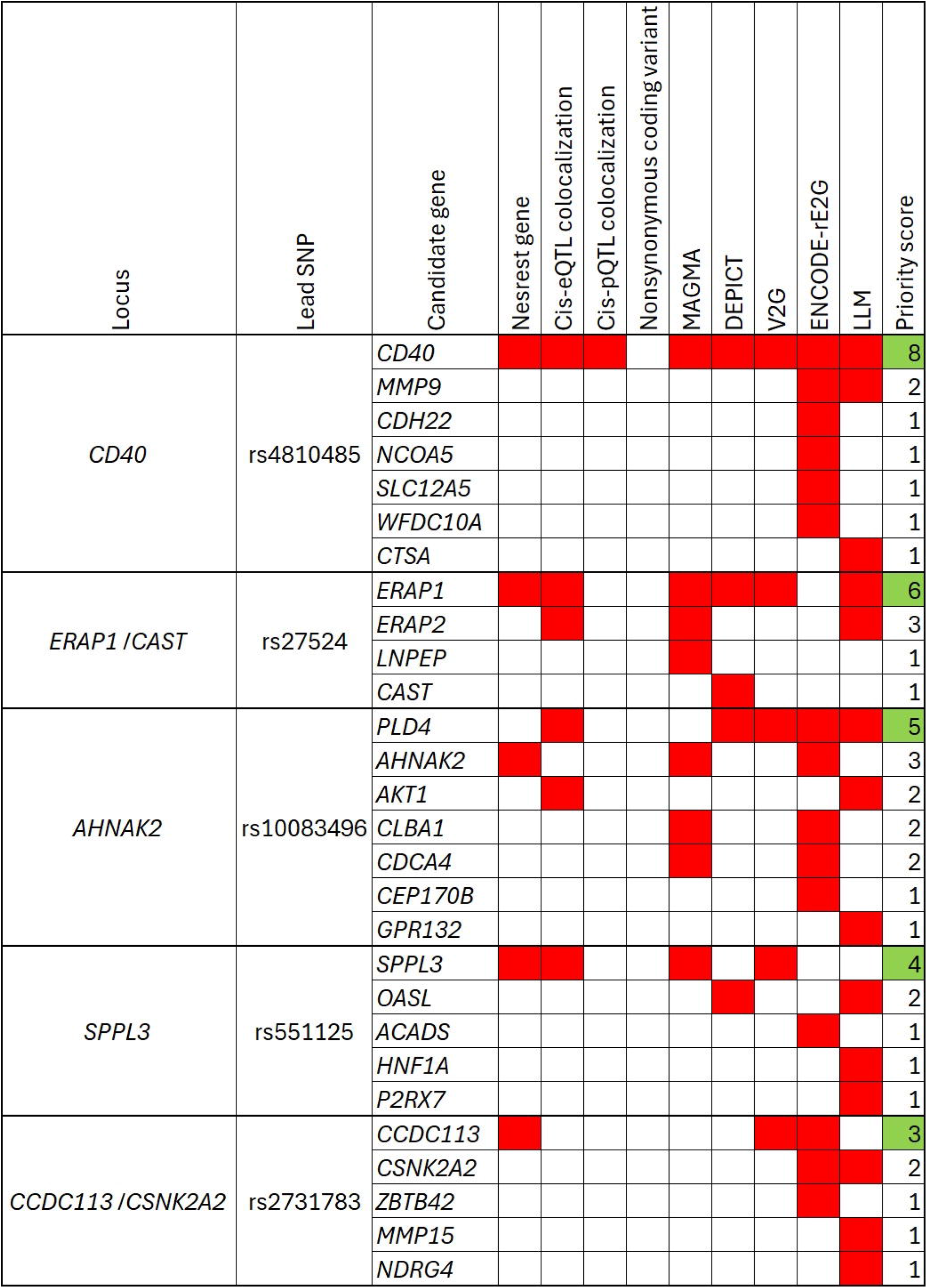
Integrative prioritization of novel candidate causal genes. We prioritized five novel candidate causal genes based on a priority score integrating nine criteria: CD40, ERAP1, PLD4, SPPL3 and CCDC113

## Discussion

Our study demonstrates a strong genetic correlation between SSc and PBC, with the correlation effect estimate comparable to that between SSc and SLE. The prevalence of PBC in SSc was 2-2.5%, lower than the 8.4-14.7% prevalence of SLE in SSc.^3,4,39-41^ Nevertheless, the prevalence of PBC in the general population, ranging from 19-402 per million persons, was also much lower than that of SLE, which ranges from 200-1500 per million persons.^42-46^ Consequently, compared to the general population, the relative risk (RR) of PBC and SLE in SSc likely mirrors their genetic correlation. Hence, our genetic correlation results, corroborated by the extent of phenotypic overlaps, support the existence of shared genetic susceptibilities and biological mechanisms between SSc and PBC.

We identified 44 significant non-HLA genomic loci in the fixed-model cross-phenotype GWAS meta-analysis. The robustness of our meta-analysis was supported by the PRS-meta-PheWAS analysis. In this analysis, using independent external datasets, SSc and PBC ranked among the top hits associated with the PRS, derived from our SSc-PBC meta-analysis statistics. Moreover, the SSc-PBC PRS demonstrated associations with a broad spectrum of autoimmune disorders, highlighting that the shared genetic susceptibilities between SSc and PBC captured by our cross-phenotype meta-analysis represent pleiotropic genomic regions.

We identified five novel loci that were significant in the cross-phenotype GWAS meta-analysis and colocalized between SSc and PBC: *CSNK2A2*/*CCDC113, SPPL3, CAST*/*ERAP1, AHNAK2*, and *CD40*. Three of the five loci were independently confirmed in a GWAS in another population: *CD40* and *AHANK2* loci were significant in a recent SSc GWAS meta-analysis that included the Japanese population,^47^ and the *CSNK2A2*/*CCDC113* locus was significant in a PBC GWAS from the Chinese population.^48^ Moreover, we found that the lead SNP in the *CD40, AHNAK2*, and *CAST*/*ERAP1* loci was associated with other autoimmune disorders in PheWAS. The pleiotropic effects observed at these loci underscore their potential role in promoting autoimmunity. We subsequently prioritized five novel candidate causal genes for SSc and PBC based on integrating nine analytic approaches: *CD40, ERAP1, PLD4, SPPL3*, and *CCDC113*.

At the *CD40* locus, the SSc-PBC meta-analysis statistics colocalized with not only the reduced transcript but also reduced plasma protein levels of CD40. Such associations have also been observed in other autoimmune diseases, including inflammatory bowel disease and multiple sclerosis, as well as malignancy in our PheWAS analysis.^49,50^ This seems paradoxical given CD40’s established role in promoting autoimmunity.^51^ However, CD40 deficiency, a rare monogenic disorder caused by bi-allelic loss-of-function variants in *CD40*, is characterized not only by humoral immunodeficiency but also by malignancy and autoimmunity, including sclerosing cholangitis and colitis.^52^ Thus, a causal relationship between reduced *CD40* expression due to polymorphisms and an increased risk of autoimmunity and malignancy is biologically plausible, mimicking the phenotypic manifestations of its monogenic disease counterpart. Interestingly, at the *CD40* locus, the SSc-PBC meta-analysis statistics also colocalized with increased levels of multiple plasma proteins involved in B cell functions, including *CD40L, FCER2, CD22, TRAF2*, and *TCL1A*. This suggests a potential compensatory response. Soluble CD40L (sCD40L), the circulating form measured in the proteomics assay, is the ligand of *CD40*, which also binds to other receptors on endothelial cells and promotes vascular pathology.^53^ Elevated soluble CD40L levels have been found in patients with SSc and are associated with its vascular manifestations.^54^ Overall, the complex B cell dysregulation mediating genetically determined reduced *CD40* expression in SSc warrants further investigation.

Our study also suggests that SSc and PBC may be associated with reduced major histocompatibility complex (MHC)-I-mediated immune response, potentially affecting cancer immunosurveillance. The SSc-PBC meta-analysis statistics in *CAST*/*ERAP1* and *SPPL3* loci colocalized with reduced transcripts of endoplasmic reticulum aminopeptidase 1 (ERAP1), increased transcripts of endoplasmic reticulum aminopeptidase 2 (ERAP2) and reduced transcripts of signal peptide peptidase-like 3 (SPPL3). These genes are implicated in the MHC-I-mediated immune response. ERAP1 and ERAP2 process and trim antigen peptides prior to their binding to MHC-I. Conditions referred to as “MHC-I-opathy”, including psoriasis and ankylosing spondylitis, were associated with *MHC-I* and *ERAP1* alleles.^10,36,55-57^ In contrast, both SSc and PBC are associated with *MHC-II* and the risk alleles in *CAST*/*ERAP1* identified in our study are in the opposite direction of ERAP1 and ERAP2 expression compared to those found in the MHC-I-opathy entities.^58,59^ Recent research has shown that reduced SPPL3 activity can dampen the MHC-I-mediated response and promote tumor immune evasion.^60^ Both SSc and PBC have been associated with an elevated risk of cancer, and recent findings indicate that SSc carries a higher burden of somatic mutations and genomic instability.^61-64^ It is possible that these changes could be related to diminished immunosurveillance from a dampened MHC-I-mediated response.

Our study has several limitations. First, we used summary statistics from published GWAS meta-analyses and were unable to perform standardized quality control with individual-level genotype data. Nevertheless, the GWAS studies included in our study were recent, comprised of large sample sizes from multiple cohorts, and led by international experts. Second, our study was performed in European ancestries; thus, our results may not be generalizable to other populations. Third, the sample size of SSc and PBC in our external datasets was limited, which precluded us from performing a replication GWAS to validate the newly discovered loci. Instead, we conducted a PRS-meta-PheWAS analysis to confirm that the PRS derived from our SSc-PBC meta-analysis captures the overall genetic risk of SSc and PBC in independent EHR-based datasets. Fourth, the sample size of GTEx and CEDAR was smaller than that of eQTLGen, which could limit statistical power in our eQTL colocalization analyses for the relevant tissues and cells. Finally, for our meta-PheWAS analyses, the diagnoses in the EHR-based datasets rely on administrative codes, which may have non-random missingness and low sensitivity for phenotype detection.^65^

In conclusion, our study revealed a strong genetic correlation between SSc and PBC and provided insights into their shared genetic susceptibility. We prioritized seven novel genes that were potentially involved in the common causal mechanisms between SSc and PBC. These discoveries prioritize therapeutic targets for both SSc and PBC. Moreover, our study advocates for heightened awareness among rheumatologists about the possibility of concurrent PBC in patients with SSc.

## Data Availability

All data produced in the present study are available upon reasonable request to the authors

## Acknowledgements and affiliations

Dr. Luo’s work was supported by the Rheumatology Research Foundation Scientist Development Award. Dr. Bernstein’s work was supported by the National Institute of Arthritis and Musculoskeletal and Skin Diseases (grant K23-AR-075112), the National Heart, Lung, and Blood Institute (grant R01-HL-164758), and the Department of Defense (grant W81XWH2210163).

Dr. Liu was supported by the National Institute of Diabetes and Digestive and Kidney Diseases (grant 1K01DK137031).

The work was previously presented at the 2023 ACR Convergence: Luo Y, Khan A, Perreault G, Liu L, Lee C, Gourh P, Pomenti S, Kiryluk K, Bernstein E. Shared Genetic Susceptibility Between Systemic Sclerosis and Primary Biliary Cholangitis: Analyses from Genome-Wide Association Studies [abstract]. Arthritis Rheumatol. 2023; 75 (suppl 9).

