## Supplemental Material for "Cross-Phenotype GWAS Supports Shared Genetic Susceptibility to Systemic Sclerosis and Primary Biliary Cholangitis"

**Supplemental Note**

**Reference datasets**

All the summary statistics datasets were harmonized against the 1000 Genomes (phase 3).^1^ Unless otherwise indicated, genotype data from the 1000 Genomes Project (phase 3) ^1^ European individuals was used for the LD reference panel for the analyses in our study. Allele frequency was calculated based on the assumption that the control individuals in the meta-analysis have the same allele frequency as 1000 Genomes (phase 3) European individuals.

**Loci definition**

We defined genomic loci using the following criteria implemented in FUMA (FUnctional Mapping and Annotation) ^2^: First, SNPs with p < 5 x 10^-8^ were considered significant SNPs. Next, SNPs in LD (r2 ≥ 0.2) within a 500 kb window with the significant SNPs were included to form an LD block. Finally, LD blocks within 500 kb of each other were merged into one locus.

**Fine-Mapping and Credible Set Analysis Using CARMA**

To screen multiple signals in loci with large window sizes, we first performed stepwise conditional analysis using GCTA-COJO ^3^ in the genomic region of a 1.5 Mb window centered on the lead SNPs of each locus. Next, we calculated 99% credible SNP sets for each genomic locus using CARMA (CAusal Robust Mapping method with Annotations). CARMA is an in-house developed Bayesian model for fine-mapping. Compared to other methods, CARMA can better account for uneven measurement of SNPs in each GWAS study of a meta-analysis, as well as the discrepancies between summary statistics and LD from external reference panels. We performed CARMA in the genomic region of a 500 kb window centered on the lead SNPs of independent signals from the conditional analysis. We incorporated functional annotation into CARMA using the prior causal probabilities based on a meta-analysis of 15 UK Biobank traits from PolyFun (POLYgenic FUNctionally-informed fine-mapping) ^4^. For loci with large window sizes and multiple independent signals in COJO, CARMA was performed in each independent signal separately, and the results were then merged into that locus.

**Large language model (LLM) candidate causal gene prioritization using GPT-4**

Large language models (LLMs) are deep learning models trained on vast amounts of text data and able to perform a large number of tasks, including answering biomedical questions.^5^ It has been recently found that GPT-4 can be used a systematic way to mine literature and identifying causal genes at GWAS loci and have achieved high performance, as their training datasets include scientific literature and other sources of information about genetics.^6^ We have used the input approach described by Shringarpure et al to perform candidate causal gene prioritization using GPT-4.^6^ All genes within 500 kb of the lead SNP at the locus were input as the candidate genes to prioritize.

**Instruction input:**

“*You are an expert in biology and genetics. Your task is to identify likely causal genes within a locus for a given GWAS phenotype based on literature evidence.*

*From the list, provide the likely causal gene (matching one of the given genes), evidence (0: very unsure to 1: very confident), and a brief reason (50 words or less) for your choice.*

*Identify the causal gene.*”

***ERAP1*/*CAST* locus Input:**

“*GWAS: systemic sclerosis and primary biliary cholangitis cross-phenotype meta-analysis*

*Genes in the locus: {PCSK1}	, {CAST}, {ERAP1}, {ERAP2}, {LNPEP}, {SETP22}, {LIX1}, {RIOK2}, {RNU1-73P}”*

***ERAP1*/*CAST* locus Output:**

“*To identify the likely causal gene within the locus for the phenotypes of systemic sclerosis and primary biliary cholangitis from the GWAS cross-phenotype meta-analysis, I will focus on genes that have been previously implicated in related immune or inflammatory pathways. Here's the evaluation based on available literature:*

*ERAP1 (Endoplasmic Reticulum Aminopeptidase 1)*

*Evidence Score: 0.8*

*Reason: ERAP1 is involved in antigen processing and presentation. Previous studies have implicated ERAP1 in autoimmune diseases through its role in modulating inflammation and immune responses, which are key components in both systemic sclerosis and primary biliary cholangitis.*

*ERAP2 (Endoplasmic Reticulum Aminopeptidase 2)*

*Evidence Score: 0.7*

*Reason: Similar to ERAP1, ERAP2 also plays a role in the processing of antigenic peptides, essential for immune regulation. Though less frequently associated with autoimmune conditions than ERAP1, its functional similarity suggests a potential role.*

*The other genes listed (PCSK1, CAST, LNPEP, SETP22, LIX1, RIOK2, RNU1-73P) have less direct connections to immune system modulation or are less studied in the context of these specific autoimmune diseases.*

*Based on the literature and functional relevance to the phenotypes in question, ERAP1 is the most likely causal gene in this locus for the GWAS phenotype of systemic sclerosis and primary biliary cholangitis.*”

***SPPL3* locus input:**

“*GWAS: systemic sclerosis and primary biliary cholangitis cross-phenotype meta-analysis*

*Genes in the locus: {PXN}, {NME2P1}, {RNU4-2}, {RNU4-1}, {SIRT4}, {RNU6-1088P}, {PLA2G1B}, {MSI1}, {RPS27P25}, {COX6A1}, {TRIAP1}, {GATC}, {RPL31P52}, {SRSF9}, {DYNLL1}, {DYNLL1-AS1}, {COQ5}, {RPL29P24}, {RNF10}, {POP5}, {RPL11P5}, {CABP1}, {MLEC}, {UNC119B}, {MIR4700}, {ACADS}, {SPPL3}, {ARF1P2}, {CLIC1P1}, {RPL12P33}, {HNF1A-AS1}, {HNF1A}, {C12orf43}, {OASL}, {P2RX7}, {P2RX4}, {CAMKK2}”*

***SPPL3* locus output:**

“*For the GWAS phenotype analysis of systemic sclerosis and primary biliary cholangitis, focusing on genes that are connected to immune function and inflammatory processes will be crucial. Here’s an evaluation of the genes in the provided locus based on their biological functions and relevance to these conditions:*

*HNF1A (Hepatocyte Nuclear Factor 1 Alpha)*

*Evidence Score: 0.85*

*Reason: HNF1A is a transcription factor important in liver function and has been implicated in autoimmune liver diseases. It influences immune response and metabolic pathways, which are central to the pathology of systemic sclerosis and primary biliary cholangitis.*

*OASL (2'-5'-Oligoadenylate Synthetase Like)*

*Evidence Score: 0.75*

*Reason: OASL is involved in antiviral responses and modulating immune signaling pathways. Given the role of viral infection and immune response in autoimmune diseases, OASL is a significant candidate.*

*P2RX7 (P2X Purinoceptor 7)*

*Evidence Score: 0.7*

*Reason: P2RX7 is a receptor involved in inflammation and cell death, processes that are enhanced in both systemic sclerosis and primary biliary cholangitis. It's linked to the regulation of immune cells and inflammatory cytokines.*

*The remaining genes, while potentially relevant, do not have as direct connections to immune modulation or inflammation in the context of systemic sclerosis and primary biliary cholangitis as HNF1A, OASL, and P2RX7. Thus, based on the functional implications and known associations, HNF1A is the most likely causal gene for this specific locus concerning the phenotypes in question.*”

***AHNAK2* locus input:**

“GWAS: systemic sclerosis and primary biliary cholangitis cross-phenotype meta-analysis

Genes in the locus: {CEND1P1}, {TMEM179}, {C14orf180}, {MIR4710}, {INF2}, {ADSSL1}, {SIVA1}, {AKT1}, {ZBTB42}, {LINC00638}, {RPS26P49}, {RPS2P4}, {CEP170B}, {PLD4}, {AHNAK2}, {C14orf79}, {CDCA4}, {GPR132}, {JAG2}, {NUDT14}, {BRF1}, {BTBD6}, {PACS2}, {RPS20P33}, {TEX22}, {MTA1}”

***AHNAK2* locus input:**

“*Given the autoimmune nature of systemic sclerosis and primary biliary cholangitis, the focus should be on genes implicated in immune regulation, cellular signaling, and inflammation. Here’s an evaluation of the most relevant genes from the provided list:*

*AKT1 (AKT Serine/Threonine Kinase 1)*

*Evidence Score: 0.85*

*Reason: AKT1 plays a crucial role in multiple cellular processes including metabolism, proliferation, cell survival, growth, and angiogenesis. It is also integral to immune cell signaling. Its dysregulation has been linked to several autoimmune and inflammatory diseases.*

*PLD4 (Phospholipase D4)*

*Evidence Score: 0.75*

*Reason: PLD4 is involved in the production of signaling molecules that play roles in inflammation and immune responses. It has been studied in the context of other autoimmune disorders, suggesting a possible role in systemic sclerosis and primary biliary cholangitis.*

*GPR132 (G Protein-Coupled Receptor 132)*

*Evidence Score: 0.7*

*Reason: GPR132 is known to be involved in immune system modulation and inflammation. It acts as a sensor for cellular damage and plays a role in the regulation of inflammatory responses.*

*While the other genes listed could potentially contribute to disease pathology through various pathways, AKT1, PLD4, and GPR132 are more directly implicated in pathways relevant to the phenotypes in question. Therefore, AKT1 is suggested as the most likely causal gene in this locus for the GWAS phenotype of systemic sclerosis and primary biliary cholangitis, based on its critical roles in cellular signaling and immune modulation.*”

***CCDC113*/*CSNK2A2* locus input:**

“*GWAS: systemic sclerosis and primary biliary cholangitis cross-phenotype meta-analysis*

*Genes in the locus: {CCDC135}, {KATNB1}, {KIFC3}, {RNU6-20P}, {CNGB1}, {TEPP}, {ZNF319}, {USB1}, {MMP15}, {C16orf80}, {CSNK2A2}, {RN7SL645P}, {CCDC113}, {PRSS54}, {GINS3}, {RNU6-269P}, {RNU6-1110P}, {NDRG4}, {RNU6-103P}, {SETD6}, {CNOT1}, {SNORA46}, {SNORA50}, {SLC38A7}, {GOT2}*”

***CCDC113*/*CSNK2A2* locus output:**

“*In evaluating the genes from the locus for systemic sclerosis and primary biliary cholangitis, the focus should again be on those with known implications in immune function, inflammation, and cellular integrity processes. Here’s an analysis of the most relevant genes:*

*MMP15 (Matrix Metallopeptidase 15)*

*Evidence Score: 0.8*

*Reason: MMP15 is part of the matrix metalloproteinase family that degrades extracellular matrix components. These enzymes play a role in tissue remodeling, inflammation, and fibrosis, which are central to the pathologies of systemic sclerosis and primary biliary cholangitis.*

*CSNK2A2 (Casein Kinase 2 Alpha 2)*

*Evidence Score: 0.75*

*Reason: CSNK2A2 is involved in various cellular processes including cell cycle control and DNA repair. It has been implicated in the signaling pathways that regulate immune responses and may influence the progression of autoimmune disorders.*

*NDRG4 (N-Myc Downstream Regulated Gene 4)*

*Evidence Score: 0.7*

*Reason: NDRG4 is known to be involved in cell growth and differentiation. It has potential roles in cellular stress responses and may be involved in inflammatory pathways, which are relevant to autoimmune diseases.*

*Among the other candidates, although several have functions that might intersect with disease mechanisms, MMP15, CSNK2A2, and NDRG4 have clearer connections to the specific pathological features of systemic sclerosis and primary biliary cholangitis. Therefore, MMP15 is suggested as the most likely causal gene due to its direct role in processes such as tissue remodeling and inflammation, critical in both conditions.*”

***CD40* locus input:**

“*GWAS: systemic sclerosis and primary biliary cholangitis cross-phenotype meta-analysis*

*Genes in the locus: {WFDC9}, {WFDC10A}, {RPS2P7}, {WFDC11}, {WFDC10B}, {WFDC13}, {MIR3617}, {SPINT4}, {HNRNPA1P3}, {WFDC3}, {SPINT5P}, {RNU6ATAC38P}, {DNTTIP1}, {UBE2C}, {TNNC2}, {SNX21}, {ACOT8}, {ZSWIM3}, {ZSWIM1}, {SPATA25}, {NEURL2}, {CTSA}, {PLTP}, {PCIF1}, {ZNF335}, {FTLP1}, {MMP9}, {SLC12A5}, {NCOA5}, {RPL13P2}, {CD40}, {CDH22}, {SLC35C2}, {ELMO2}, {ZNF663P}, {MKRN7P}, {ZNF840}, {ZNF334}, {OCSTAMP}, {SLC13A3}*”

***CD40* locus output:**

“*For the GWAS cross-phenotype meta-analysis of systemic sclerosis and primary biliary cholangitis, it's crucial to focus on genes that have known connections to immune regulation, inflammation, fibrosis, and cellular signaling. Here's a detailed look at the top relevant genes:*

*CD40 (CD40 Molecule)*

*Evidence Score: 0.9*

*Reason: CD40 is a costimulatory protein found on antigen-presenting cells and is essential for initiating specific immune responses. Its interaction with CD40L on T cells is crucial for the activation of both humoral and cellular immune responses, which are dysregulated in autoimmune diseases like systemic sclerosis and primary biliary cholangitis.*

*MMP9 (Matrix Metallopeptidase 9)*

*Evidence Score: 0.85*

*Reason: MMP9 is involved in the breakdown of extracellular matrix in normal physiological processes such as tissue remodeling, as well as in disease states including fibrosis and inflammation. This enzyme is therefore relevant to the fibrotic processes observed in systemic sclerosis and the inflammatory responses in primary biliary cholangitis.*

*CTSA (Cathepsin A)*

*Evidence Score: 0.75*

*Reason: CTSA encodes a lysosomal enzyme that plays a role in the degradation of proteins and the production of certain bioactive peptides. It is involved in immune system processes and may contribute to the pathology of lysosomal storage diseases, which have immunological components.*

*Given the significant roles of these genes in immune regulation and fibrosis, CD40 emerges as the most likely causal gene due to its critical involvement in immune cell activation and its established association with various autoimmune conditions. This makes CD40 a highly plausible candidate for influencing both systemic sclerosis and primary biliary cholangitis.”*

**Supplemental Table 1** Significant genomic loci in the coss-phenotype GWAS meta-analysis between SSc and PBC using the fixed-effect model

| Locus | Lead SNP | CHR | BP | A1 | A2 | Meta-analysis OR | Meta-analysis P-Values | Heterogeneity | | SSc OR | SSc p-values | PBC OR | PBC p-values | Novel loci | |
| --- | --- | --- | --- | --- | --- | --- | --- | --- | --- | --- | --- | --- | --- | --- | --- |
| *IL12RB2* | rs1874791 | 1 | 67806432 | A | G | 1.32 | 9.59E-30 | | Yes | 1.18 | 1.09E-02 | 1.35 | 1.87E-29 | | No |
| *CD58* | rs11588376 | 1 | 117072524 | C | T | 0.83 | 2.29E-08 | | No | 0.89 | 6.92E-02 | 0.81 | 5.15E-08 | | No |
| *FCRL3* | rs7528684 | 1 | 157670816 | G | A | 0.9 | 3.41E-09 | | No | 0.93 | 9.38E-03 | 0.89 | 2.12E-08 | | No |
| *LOC100506023* | rs2022449 | 1 | 173238736 | T | G | 1.12 | 1.69E-10 | | No | 1.15 | 6.28E-08 | 1.1 | 7.18E-05 | | No |
| *CRB1* | rs4915526 | 1 | 197331817 | C | T | 0.88 | 2.42E-08 | | Yes | 0.88 | 4.89E-02 | 0.88 | 1.27E-07 | | No |
| *C1orf106* | rs55838263 | 1 | 200874728 | G | A | 0.88 | 2.61E-08 | | No | 0.94 | 2.35E-01 | 0.87 | 1.79E-08 | | No |
| *DNMT3A* | rs75904251 | 2 | 25541953 | A | G | 0.81 | 3.24E-09 | | No | 0.93 | 3.94E-01 | 0.79 | 1.07E-09 | | No |
| *TMEM163* | rs842362 | 2 | 135341120 | A | G | 0.89 | 1.86E-08 | | No | 0.95 | 2.03E-01 | 0.87 | 9.99E-09 | | No |
| *STAT4* | rs11889341 | 2 | 191943742 | T | C | 1.27 | 1.64E-23 | | Yes | 1.34 | 1.84E-03 | 1.27 | 7.43E-22 | | No |
| *PLCL2* | rs11128810 | 3 | 16960181 | A | G | 1.14 | 2.91E-12 | | No | 1.1 | 2.45E-03 | 1.16 | 8.14E-11 | | No |
| *RARB* | rs12496365 | 3 | 25387754 | C | G | 1.14 | 1.43E-09 | | Yes | 1.08 | 2.91E-02 | 1.16 | 3.96E-09 | | No |
| *ARHGAP31* | rs9834901 | 3 | 119111870 | C | T | 0.77 | 1.42E-25 | | Yes | 0.81 | 1.30E-06 | 0.75 | 1.68E-21 | | No |
| *SLC15A2* | rs9843053 | 3 | 121617433 | A | G | 1.12 | 3.34E-09 | | No | 1.1 | 6.62E-04 | 1.14 | 4.49E-07 | | Yes |
| *IQCJ-SCHIP1/SCHIP1* | rs4680527 | 3 | 159568050 | T | A | 0.77 | 4.43E-30 | | Yes | 0.85 | 7.01E-02 | 0.76 | 5.17E-30 | | No |
| *GAK* | rs3755963 | 4 | 894255 | G | A | 1.11 | 4.34E-10 | | No | 1.12 | 9.74E-06 | 1.11 | 3.47E-06 | | No |
| *NFKB1* | rs230534 | 4 | 103449041 | T | C | 1.17 | 1.03E-21 | | No | 1.15 | 5.38E-09 | 1.19 | 2.55E-15 | | No |
| *CAST/ERAP1* | rs27524 | 5 | 96101944 | A | G | 0.9 | 4.89E-10 | | No | 0.87 | 4.60E-07 | 0.91 | 3.34E-05 | | Yes |
| *ST8SIA4* | rs3756353 | 5 | 100239187 | T | C | 1.1 | 1.45E-08 | | No | 1.08 | 1.10E-03 | 1.12 | 1.25E-06 | | Yes |
| *TNIP1* | rs1422673 | 5 | 150438988 | T | C | 1.12 | 2.87E-09 | | Yes | 1.16 | 1.94E-07 | 1.1 | 3.51E-04 | | No |
| *UBLCP1* | rs10042630 | 5 | 158710768 | A | T | 1.14 | 4.42E-10 | | No | 1.11 | 2.51E-04 | 1.16 | 1.25E-07 | | No |
| *OLIG3/LOC102723649* | rs62432712 | 6 | 137964697 | G | A | 1.15 | 1.68E-10 | | No | 1.09 | 1.96E-02 | 1.18 | 4.37E-10 | | No |
| *ITGB8* | rs6944997 | 7 | 20438370 | C | A | 1.12 | 5.09E-09 | | No | 1.09 | 6.81E-03 | 1.13 | 8.27E-08 | | No |
| *ELMO1* | rs16879645 | 7 | 37400469 | C | T | 1.24 | 4.46E-14 | | No | 1.16 | 2.06E-03 | 1.27 | 9.13E-13 | | No |
| *TNPO3* | rs17338998 | 7 | 128618559 | T | C | 1.52 | 2.37E-40 | | No | 1.4 | 6.03E-21 | 1.52 | 9.71E-41 | | No |
| *HEMGN/ANP32B* | rs4743150 | 9 | 100740124 | T | C | 0.88 | 8.29E-11 | | No | 0.89 | 1.80E-04 | 0.86 | 3.10E-08 | | No |
| *PHRF1* | rs7943546 | 11 | 612148 | C | T | 0.88 | 1.07E-09 | | Yes | 0.85 | 6.18E-06 | 0.89 | 7.80E-06 | | No |
| *RPS6KA4/LOC100996455* | rs475032 | 11 | 64140737 | G | C | 0.9 | 5.71E-09 | | Yes | 0.94 | 4.78E-02 | 0.88 | 4.86E-09 | | No |
| *TREH/DDX6* | rs77209083 | 11 | 118607569 | G | A | 0.74 | 4.88E-29 | | Yes | 0.82 | 8.76E-03 | 0.73 | 1.99E-28 | | No |
| *ATXN2* | rs598710 | 12 | 111973141 | C | A | 0.84 | 9.07E-12 | | Yes | 0.9 | 9.44E-02 | 0.82 | 1.12E-11 | | No |
| *SPPL3* | rs551125 | 12 | 121203427 | C | T | 0.91 | 1.65E-09 | | No | 0.9 | 5.42E-06 | 0.91 | 2.33E-05 | | Yes |
| *LINC02341* | rs9533117 | 13 | 43046812 | T | A | 0.87 | 2.88E-08 | | Yes | 0.95 | 6.82E-01 | 0.86 | 2.03E-08 | | No |
| *DLEU1* | rs74899623 | 13 | 50794228 | A | G | 0.73 | 2.78E-08 | | Yes | 0.84 | 9.13E-02 | 0.69 | 2.56E-08 | | No |
| *RIN3* | rs734206 | 14 | 93108464 | C | T | 0.87 | 7.77E-10 | | No | 0.94 | 2.59E-01 | 0.86 | 3.98E-10 | | No |
| *AHNAK2* | rs10083496 | 14 | 105402786 | G | A | 1.09 | 2.56E-08 | | No | 1.09 | 3.31E-05 | 1.09 | 7.75E-05 | | Yes |
| *PRM1/LOC105371083* | rs4451969 | 16 | 11383519 | T | C | 0.84 | 3.51E-17 | | Yes | 0.89 | 8.42E-03 | 0.83 | 1.98E-16 | | No |
| *CSNK2A2/CCDC113* | rs2731783 | 16 | 58253460 | A | G | 1.15 | 7.79E-11 | | No | 1.15 | 1.22E-06 | 1.15 | 4.00E-06 | | Yes |
| *DPEP2/LOC100131303* | rs7204192 | 16 | 68039309 | G | A | 1.21 | 1.58E-11 | | No | 1.13 | 1.01E-01 | 1.22 | 2.89E-11 | | No |
| *IRF8/LINC01082* | rs17445836 | 16 | 86017663 | A | G | 0.77 | 1.08E-26 | | No | 0.8 | 1.17E-06 | 0.77 | 2.65E-22 | | No |
| *IKZF3* | rs8071789 | 17 | 38006333 | C | T | 0.78 | 3.19E-33 | | Yes | 0.84 | 2.47E-03 | 0.77 | 3.83E-32 | | No |
| *KANSL1* | rs17660228 | 17 | 44166500 | A | G | 0.87 | 1.56E-11 | | No | 0.9 | 5.28E-04 | 0.85 | 9.88E-10 | | No |
| *CD226* | rs1788098 | 18 | 67542798 | T | C | 1.11 | 1.89E-09 | | No | 1.08 | 1.03E-02 | 1.13 | 1.53E-08 | | No |
| *IL12RB1* | rs2305743 | 19 | 18193191 | A | G | 0.86 | 1.95E-14 | | No | 0.83 | 4.64E-10 | 0.87 | 4.60E-07 | | No |
| *CD40* | rs4810485 | 20 | 44747947 | T | G | 1.13 | 1.44E-08 | | No | 1.18 | 3.02E-05 | 1.11 | 1.96E-05 | | Yes |
| *RPL3/SYNGR1* | rs5757622 | 22 | 39728861 | T | C | 1.24 | 3.29E-22 | | Yes | 1.16 | 7.23E-05 | 1.28 | 4.05E-20 | | No |

1. The lead SNP is the SNP with the smallest meta-analysis p-values after excluding all the SNPs with P_het_ < 0.05 2. The SNPs with the smallest meta-analysis p-values (prior to filtering on heterogeneity) had a P_het_ < 0.05. 3. If a locus had an independent signal in either SSc or PBC, additional colocalization analysis was performed using the conditional analysis statistics and the highest PP4 was reported. PP4: the posterior probability of hypothesis 4 that there is at least one variant causal to both traits. SSc: systemic sclerosis. PBC: primary biliary cholangitis. A1: effect allele. A2: non-effect allele.

**Supplemental Table 2** Significant genomic loci in the coss-phenotype GWAS meta-analysis between SSc and PBC using PLEIO

| Locus | Lead SNP | CHR | BP | A1 | A2 | PLEIO p-values | SSc OR | SSc p-values | PBC OR | PBC p-values | Significant in fixed-effect model | Novel loci |
| --- | --- | --- | --- | --- | --- | --- | --- | --- | --- | --- | --- | --- |
| *MMEL1* | rs3828154 | 1 | 2524943 | A | G | 3.10E-09 | 1.02 | 2.95E-01 | 1.14 | 4.16E-09 | No | No |
| *IL12RB2* | rs6679356 | 1 | 67820194 | C | T | 1.26E-86 | 1.17 | 4.80E-04 | 1.55 | 6.61E-63 | Yes | No |
| *CD58* | rs10924109 | 1 | 117079262 | C | T | 9.18E-09 | 0.91 | 9.18E-02 | 0.81 | 2.88E-08 | Yes | No |
| *FCRL3* | rs7528684 | 1 | 157670816 | G | A | 1.77E-09 | 0.93 | 9.38E-03 | 0.89 | 2.12E-08 | Yes | No |
| *CD247* | rs2056626 | 1 | 167420425 | G | T | 5.09E-09 | 0.81 | 1.31E-11 | 0.99 | 8.13E-01 | No | No |
| *LOC100506023* | rs2022449 | 1 | 173238736 | T | G | 2.82E-09 | 1.15 | 6.28E-08 | 1.10 | 7.18E-05 | Yes | No |
| *DENND1B/C1orf53* | rs12123169 | 1 | 197780966 | A | T | 4.00E-20 | 1.04 | 3.92E-01 | 1.24 | 9.75E-18 | Yes | No |
| *C1orf106* | rs55838263 | 1 | 200874728 | G | A | 1.08E-08 | 0.94 | 2.35E-01 | 0.87 | 1.79E-08 | Yes | No |
| *DNMT3A* | rs934613 | 2 | 25514896 | T | G | 4.59E-10 | 0.94 | 3.05E-01 | 0.79 | 5.57E-10 | Yes | No |
| *TMEM163* | rs859767 | 2 | 135341200 | G | A | 2.17E-09 | 0.98 | 6.50E-01 | 0.87 | 1.54E-09 | Yes | No |
| *STAT4* | rs3821236 | 2 | 191902758 | A | G | 3.53E-34 | 1.31 | 1.94E-23 | 1.19 | 9.75E-11 | Yes | No |
| *PLCL2* | rs11128810 | 3 | 16960181 | A | G | 1.11E-12 | 1.10 | 2.45E-03 | 1.16 | 8.14E-11 | Yes | No |
| *RARB* | rs11920829 | 3 | 25381124 | A | C | 1.13E-15 | 0.94 | 4.43E-02 | 1.18 | 4.47E-14 | Yes | No |
| *PXK* | rs4076852 | 3 | 58375286 | G | A | 6.47E-10 | 1.16 | 1.04E-10 | 1.07 | 3.57E-03 | No | No |
| *ARHGAP31* | rs9884090 | 3 | 119116150 | A | G | 1.80E-34 | 0.83 | 1.89E-10 | 0.75 | 1.46E-21 | Yes | No |
| *SLC15A2* | rs9843053 | 3 | 121617433 | A | G | 8.98E-09 | 1.10 | 6.62E-04 | 1.14 | 4.49E-07 | Yes | Yes |
| *IL12A-AS1* | rs589446 | 3 | 159733527 | T | G | 1.68E-82 | 0.86 | 1.95E-10 | 0.70 | 1.96E-55 | Yes | No |
| *GAK* | rs3755963 | 4 | 894255 | G | A | 5.53E-09 | 1.12 | 9.74E-06 | 1.11 | 3.47E-06 | Yes | No |
| *MANBA* | rs228611 | 4 | 103561709 | A | G | 1.38E-24 | 0.89 | 1.29E-03 | 0.82 | 3.36E-19 | Yes | No |
| *IL7R/CAPSL* | rs11742240 | 5 | 35881376 | T | G | 1.77E-21 | 1.00 | 8.81E-01 | 0.80 | 6.43E-19 | No | No |
| *CAST/ERAP1* | rs27524 | 5 | 96101944 | A | G | 6.04E-09 | 0.87 | 4.60E-07 | 0.91 | 3.34E-05 | Yes | Yes |
| *ST8SIA4/SLCO4C1* | rs17782250 | 5 | 100249342 | A | T | 2.52E-08 | 1.06 | 1.44E-02 | 1.12 | 2.36E-07 | Yes | Yes |
| *NDFIP1* | rs10062349 | 5 | 141509597 | G | A | 4.75E-08 | 1.05 | 3.93E-02 | 0.89 | 7.36E-08 | No | Yes |
| *TNIP1* | rs3792783 | 5 | 150455732 | G | A | 7.37E-12 | 1.20 | 2.42E-12 | 1.10 | 1.41E-03 | Yes | No |
| *UBLCP1* | rs10042630 | 5 | 158710768 | A | T | 1.42E-09 | 1.11 | 2.51E-04 | 1.16 | 1.25E-07 | Yes | No |
| *ATG5* | rs633724 | 6 | 106734040 | T | C | 2.98E-08 | 1.13 | 2.84E-09 | 1.06 | 1.20E-02 | No | No |
| *LOC102723649/LOC100507406* | rs11757201 | 6 | 138003822 | C | G | 3.21E-11 | 1.07 | 1.34E-02 | 1.17 | 3.46E-10 | Yes | No |
| *ITGB8* | rs6944997 | 7 | 20438370 | C | A | 5.73E-09 | 1.09 | 6.81E-03 | 1.13 | 8.27E-08 | Yes | No |
| *ELMO1* | rs16879645 | 7 | 37400469 | C | T | 1.89E-15 | 1.16 | 2.06E-03 | 1.27 | 9.13E-13 | Yes | No |
| *TNPO3* | rs34871361 | 7 | 128671086 | T | C | 4.16E-73 | 1.40 | 3.12E-21 | 1.52 | 1.03E-40 | Yes | No |
| *FAM167A/BLK* | rs2736340 | 8 | 11343973 | T | C | 6.72E-20 | 1.24 | 3.33E-21 | 1.01 | 7.93E-01 | No | No |
| *HEMGN/ANP32B* | rs4743150 | 9 | 100740124 | T | C | 2.80E-10 | 0.89 | 1.80E-04 | 0.86 | 3.10E-08 | Yes | No |
| *PHRF1* | rs702966 | 11 | 611919 | G | C | 1.34E-09 | 0.80 | 2.12E-07 | 0.90 | 1.30E-05 | Yes | No |
| *PRDX5* | rs627425 | 11 | 64087642 | T | C | 1.56E-11 | 0.94 | 1.13E-02 | 0.87 | 2.22E-10 | Yes | No |
| *POU2AF1* | rs11605031 | 11 | 111240607 | G | A | 2.66E-08 | 0.97 | 3.23E-01 | 0.88 | 3.55E-08 | No | No |
| *DDX6/CXCR5* | rs4936443 | 11 | 118740864 | C | T | 2.62E-50 | 0.82 | 3.05E-07 | 0.69 | 5.39E-35 | Yes | No |
| *TNFRSF1A* | rs1800693 | 12 | 6440009 | C | T | 5.25E-19 | 0.95 | 9.21E-02 | 1.20 | 1.19E-16 | No | No |
| *PPHLN1* | rs1796357 | 12 | 42815553 | C | T | 3.17E-08 | 1.05 | 1.33E-02 | 1.18 | 3.08E-07 | No | Yes |
| *NAA25* | rs17696736 | 12 | 112486818 | G | A | 1.44E-16 | 1.04 | 2.16E-01 | 1.18 | 5.88E-15 | No | No |
| *SPPL3* | rs551125 | 12 | 121203427 | C | T | 2.59E-08 | 0.90 | 5.42E-06 | 0.91 | 2.33E-05 | Yes | Yes |
| *LINC02341* | rs12430303 | 13 | 43032027 | T | C | 1.15E-13 | 1.05 | 6.48E-02 | 0.86 | 8.79E-13 | Yes | No |
| *DLEU1* | rs9591325 | 13 | 50811220 | C | T | 2.41E-23 | 0.90 | 4.68E-02 | 0.64 | 2.14E-19 | Yes | No |
| *RAD51B* | rs8008961 | 14 | 68752643 | T | C | 3.52E-19 | 0.96 | 1.80E-01 | 0.81 | 8.35E-17 | No | No |
| *RIN3* | rs72699846 | 14 | 93098617 | A | G | 5.37E-11 | 0.96 | 1.92E-01 | 0.83 | 9.43E-11 | Yes | No |
| *LOC107984640/EXOC3L4* | rs59643720 | 14 | 103564807 | C | A | 1.12E-50 | 0.89 | 3.43E-02 | 1.37 | 2.73E-38 | No | No |
| *CSK* | rs1378942 | 15 | 75077367 | C | A | 1.10E-12 | 1.18 | 1.84E-14 | 0.97 | 1.37E-01 | No | No |
| *CLEC16A* | rs12928537 | 16 | 11191400 | A | G | 4.66E-28 | 0.99 | 6.12E-01 | 0.79 | 1.55E-23 | Yes | No |
| *IL4R/IL21R* | rs1119132 | 16 | 27403469 | A | G | 4.66E-10 | 1.16 | 5.47E-02 | 0.82 | 6.58E-10 | No | No |
| *CSNK2A2/CCDC113* | rs2731783 | 16 | 58253460 | A | G | 1.50E-09 | 1.15 | 1.22E-06 | 1.15 | 4.00E-06 | Yes | Yes |
| *DPEP2/LOC100131303* | rs7204192 | 16 | 68039309 | G | A | 6.88E-12 | 1.13 | 1.01E-01 | 1.22 | 2.89E-11 | Yes | No |
| *IRF8/LINC01082* | rs17445836 | 16 | 86017663 | A | G | 8.75E-32 | 0.80 | 1.17E-06 | 0.77 | 2.65E-22 | Yes | No |
| *IKZF3* | rs9303277 | 17 | 37976469 | C | T | 5.76E-48 | 0.89 | 9.88E-08 | 0.77 | 5.31E-33 | Yes | No |
| *KANSL1* | rs17577094 | 17 | 44187492 | G | A | 2.47E-12 | 0.92 | 3.77E-03 | 0.84 | 1.13E-10 | Yes | No |
| *DOK6/CD226* | rs1808094 | 18 | 67526026 | T | C | 4.42E-10 | 1.06 | 2.69E-02 | 1.14 | 2.79E-09 | Yes | No |
| *TYK2* | rs2304256 | 19 | 10475652 | A | C | 3.93E-19 | 1.05 | 3.05E-01 | 0.81 | 4.43E-17 | No | No |
| *IL12RB1* | rs2305743 | 19 | 18193191 | A | G | 7.42E-14 | 0.83 | 4.64E-10 | 0.87 | 4.60E-07 | Yes | No |
| *SPIB* | rs3745516 | 19 | 50926742 | A | G | 6.81E-38 | 1.03 | 2.36E-01 | 1.32 | 2.65E-30 | No | No |
| *RPL3/SYNGR1* | rs137685 | 22 | 39739628 | T | C | 7.12E-30 | 0.91 | 4.19E-03 | 0.80 | 2.41E-23 | Yes | No |

PP4: the posterior probability of hypothesis 4 that there is at least one variant causal to both traits. SSc: systemic sclerosis. PBC: primary biliary cholangitis. A1: effect allele. A2: non-effect allele.

**Supplemental Table 3** Credible SNP set in the fine-mapped genomic loci

| CHR | | Credible set locus | Credible set | Number of SNPs in the credible set | Lead SNP | PIP | SNP PIP_Max_ | PIP_Max_ | SNP PIP_Max_ functional annotation | Functional annotation in the 99% credible set |
| --- | --- | --- | --- | --- | --- | --- | --- | --- | --- | --- |
| 1 | *IL12RB2* | | 1 | 2 | rs1874791 | 38% | rs6660836 | 62% | intronic | intronic |
|  |  |  | 2 | 7 | rs1874791 | 38% | rs6676606 | 34% | intronic | intronic |
| 1 | *CD58* | | 1 | 18 | rs11588376 | 15% | rs11588376 | 15% | intronic | intergenic, UTR3, intronic |
| 1 | *FCRL3* | | 1 | 7 | rs7528684 | 43% | rs7528684 | 43% | upstream | intergenic, intronic, upstream |
| 1 | *LOC100506023* | | 1 | 11 | rs2022449 | 0% | rs704840 | 33% | ncRNA_intronic | ncRNA_intronic |
| 1 | *CRB1* | | 1 | 4 | rs4915526 | 69% | rs4915526 | 69% | intronic | intronic |
| 1 | *C1orf106* | | 1 | 11 | rs55838263 | 16% | rs55838263 | 16% | intronic | intronic, intergenic |
| 2 | *DNMT3A* | | 1 | 2 | rs75904251 | 56% | rs75904251 | 56% | intronic | intronic |
| 2 | *TMEM163* | | 1 | 1 | rs842362 | 100% | rs842362 | 100% | intronic | intronic |
| 2 | *STAT4* | | 1 | 3 | rs11889341 | 18% | rs4853516 | 93% | intronic | intronic, intergenic |
|  |  |  | 1 | 5 | rs11889341 | 18% | rs1263128 | 65% | intergenic | intergenic |
|  |  |  | 1 | 2 | rs11889341 | 18% | rs3024886 | 88% | intronic | intronic |
| 3 | *PLCL2* | | 1 | 7 | rs11128810 | 35% | rs11128810 | 35% | intronic | intronic |
| 3 | *RARB* | | 1 | 5 | rs12496365 | 0% | rs12497123 | 34% | intronic | intronic |
| 3 | *ARHGAP31* | | 1 | 1 | rs9834901 | 100% | rs9834901 | 100% | intronic | intronic |
| 3 | *SLC15A2* | | 1 | 41 | rs9843053 | 33% | rs9843053 | 33% | intronic | intronic, exonic, downstream, intergenic |
| 3 | *IQCJ-SCHIP1/SCHIP1* | | 1 | 1 | rs4680527 | 100% | rs4680527 | 100% | intronic | intronic |
|  |  |  | 2 | 7 | rs4680527 | 100% | rs1366301 | 38% | ncRNA_intronic | ncRNA_intronic |
|  |  |  | 3 | 2 | rs4680527 | 100% | rs2243131 | 61% | ncRNA_exonic | ncRNA_intronic, ncRNA_exonic |
|  |  |  | 4 | 1 | rs4680527 | 100% | rs9880646 | 100% | ncRNA_intronic | ncRNA_intronic |
|  |  |  | 5 | 14 | rs4680527 | 100% | rs13062928 | 23% | intergenic | intergenic |
| 4 | *GAK* | | 1 | 6 | rs3755963 | 86% | rs3755963 | 86% | intronic | intronic |
| 4 | *NFKB1* | | 1 | 3 | rs230534 | 93% | rs230534 | 93% | intronic | intronic |
| 5 | *CAST/ERAP1* | | 1 | 12 | rs27524 | 46% | rs27524 | 46% | intronic | intronic, exonic |
| 5 | *ST8SIA4* | | 1 | 88 | rs3756353 | 0% | rs3756354 | 6% | upstream | intronic, UTR3, UTR5, upstream, intergenic |
| 5 | *TNIP1* | | 1 | 7 | rs1422673 | 52% | rs1422673 | 52% | intronic | exonic, intronic |
| 5 | *UBLCP1* | | 1 | 8 | rs10042630 | 24% | rs10042630 | 24% | intronic | intronic |
| 6 | *OLIG3/LOC102723649* | | 1 | 2 | rs62432712 | 79% | rs62432712 | 79% | intergenic | intergenic |
|  |  |  | 2 | 7 | rs62432712 | 79% | rs719150 | 46% | intronic | ncRNA_intronic, intronic |
| 7 | *ITGB8* | | 1 | 14 | rs6944997 | 50% | rs6944997 | 50% | intronic | intronic |
| 7 | *ELMO1* | | 1 | 35 | rs16879645 | 21% | rs16879645 | 21% | intronic | intronic |
| 7 | *TNPO3* | | 1 | 1 | rs17338998 | 6% | rs4728142 | 100% | intergenic | intergenic |
| 7 |  |  | 2 | 17 | rs17338998 | 6% | rs13236009 | 15% | intronic | UTR3, downstream, intronic |
| 9 | *HEMGN/ANP32B* | | 1 | 9 | rs4743150 | 32% | rs4743150 | 32% | intergenic | intergenic, intronic, downstream |
| 11 | *TREH/DDX6* | | 1 | 2 | rs77209083 | 47% | rs11217001 | 53% | intergenic | intergenic |
| 12 | *ATXN2* | | 1 | 3 | rs598710 | 34% | rs598711 | 35% | intronic | intronic |
| 12 | *SPPL3* | | 1 | 27 | rs551125 | 29% | rs551125 | 29% | intronic | intergenic, downstream, UTR3, intronic |
| 13 | *LINC02341* | | 1 | 1 | rs9533117 | 100% | rs9533117 | 100% | ncRNA_intronic | ncRNA_intronic |
| 13 | *DLEU1* | | 1 | 1 | rs74899623 | 99% | rs74899623 | 99% | ncRNA_intronic | ncRNA_intronic |
| 14 | *RIN3* | | 1 | 1 | rs734206 | 100% | rs734206 | 100% | intronic | intronic |
| 14 | *AHNAK2* | | 1 | 5 | rs10083496 | 0% | rs11851053 | 34% | exonic | downstream, exonic |
| 16 | *PRM1/LOC105371083* | | 1 | 2 | rs4451969 | 100% | rs7194305 | 96% | intronic | intronic |
|  |  |  | 2 | 1 | rs4451969 | 100% | rs4451969 | 100% | intergenic | intergenic |
| 16 | *CSNK2A2/CCDC113* | | 1 | 20 | rs2731783 | 19% | rs2731783 | 19% | intergenic | intergenic |
| 16 | *DPEP2/LOC100131303* | | 1 | 3 | rs7204192 | 24% | rs113704872 | 71% | intronic | intronic, intergenic |
| 16 | *IRF8/LINC01082* | | 1 | 19 | rs17445836 | 100% | rs880364 | 20% | intergenic | intronic, UTR3, downstream, intergenic |
|  |  |  | 2 | 1 | rs17445836 | 100% | rs17445836 | 100% | intergenic | intergenic |
| 17 | *KANSL1* | | 1 | 15 | rs17660228 | 0% | rs8071789 | 100% | intronic | intronic, ncRNA_intronic |
|  | *IKZF3* | | 2 | 1 | rs8071789 | 100% | rs8071789 | 100% | intronic | intronic |
| 18 | *CD226* | | 1 | 8 | rs1788098 | 32% | rs1788098 | 32% | intronic | intergenic, UTR3, intronic |
| 19 | *IL12RB1* | | 1 | 8 | rs2305743 | 92% | rs2305743 | 92% | intronic | intronic, UTR3 |
| 20 | *CD40* | | 1 | 8 | rs4810485 | 44% | rs4810485 | 44% | intronic | UTR3, intronic, intergenic, UTR5 |
| 22 | *RPL3/SYNGR1* | | 1 | 8 | rs5757622 | 0% | rs62228376 | 21% | intergenic | intergenic |

PIP: posterior inclusion probability.

**Supplemental Table 4** Tissue enrichment analysis of the GTEx V8 based on MAGMA analysis

| Tissue name | Beta | Beta SD | SE | P-values | Bonferroni-corrected p-values |
| --- | --- | --- | --- | --- | --- |
| Spleen | 0.050031 | 0.097375 | 0.00914 | 2.24E-08 | 1.21E-06 |
| Whole_Blood | 0.038281 | 0.067865 | 0.007056 | 2.94E-08 | 1.59E-06 |
| Cells_EBV-transformed_lymphocytes | 0.025418 | 0.055131 | 0.005999 | 1.14E-05 | 6.16E-04 |
| Lung | 0.04497 | 0.08526 | 0.011583 | 5.20E-05 | 2.81E-03 |
| Small_Intestine_Terminal_Ileum | 0.045461 | 0.081828 | 0.012094 | 8.57E-05 | 4.63E-03 |
| Thyroid | 0.032282 | 0.062752 | 0.012288 | 0.00431 | 2.33E-01 |
| Liver | 0.015592 | 0.027574 | 0.007798 | 0.022784 | 1 |
| Artery_Aorta | 0.022515 | 0.045266 | 0.012738 | 0.038582 | 1 |
| Adipose_Visceral_Omentum | 0.020842 | 0.039795 | 0.013815 | 0.065708 | 1 |
| Kidney_Cortex | 0.013654 | 0.023411 | 0.011309 | 0.11365 | 1 |
| Ovary | 0.014505 | 0.028948 | 0.012158 | 0.11644 | 1 |
| Adipose_Subcutaneous | 0.014528 | 0.028546 | 0.01331 | 0.13753 | 1 |
| Artery_Coronary | 0.015721 | 0.030774 | 0.014726 | 0.14286 | 1 |
| Uterus | 0.014106 | 0.028285 | 0.013559 | 0.1491 | 1 |
| Cervix_Endocervix | 0.014008 | 0.027214 | 0.014474 | 0.16659 | 1 |
| Fallopian_Tube | 0.013151 | 0.02506 | 0.014826 | 0.18754 | 1 |
| Bladder | 0.013372 | 0.025577 | 0.016297 | 0.20597 | 1 |
| Colon_Transverse | 0.010359 | 0.018654 | 0.014575 | 0.23863 | 1 |
| Nerve_Tibial | 0.008392 | 0.016582 | 0.012385 | 0.24903 | 1 |
| Kidney_Medulla | 0.007596 | 0.013718 | 0.011652 | 0.25724 | 1 |
| Breast_Mammary_Tissue | 0.007383 | 0.013946 | 0.015677 | 0.31884 | 1 |
| Stomach | 0.007062 | 0.012556 | 0.015102 | 0.32004 | 1 |
| Cervix_Ectocervix | 0.003544 | 0.006827 | 0.015061 | 0.40699 | 1 |
| Testis | 0.000917 | 0.0015506 | 0.006982 | 0.44774 | 1 |
| Skin_Not_Sun_Exposed_Suprapubic | 0.000622 | 0.0011782 | 0.009794 | 0.47469 | 1 |
| Vagina | -0.00015 | -0.00027439 | 0.013861 | 0.50421 | 1 |
| Artery_Tibial | -0.0008 | -0.0016179 | 0.012459 | 0.5255 | 1 |
| Skin_Sun_Exposed_Lower_leg | -0.00063 | -0.0011994 | 0.009815 | 0.52555 | 1 |
| Adrenal_Gland | -0.00414 | -0.007842 | 0.012375 | 0.63092 | 1 |
| Prostate | -0.0064 | -0.011897 | 0.015233 | 0.66282 | 1 |
| Esophagus_Mucosa | -0.00442 | -0.0083981 | 0.009629 | 0.67701 | 1 |
| Minor_Salivary_Gland | -0.00637 | -0.011616 | 0.012075 | 0.70099 | 1 |
| Pancreas | -0.00743 | -0.012134 | 0.010847 | 0.75333 | 1 |
| Colon_Sigmoid | -0.01224 | -0.023499 | 0.015561 | 0.78417 | 1 |
| Cells_Cultured_fibroblasts | -0.00658 | -0.013956 | 0.008102 | 0.79167 | 1 |
| Esophagus_Gastroesophageal_Junction | -0.01313 | -0.02536 | 0.016 | 0.79402 | 1 |
| Pituitary | -0.01042 | -0.018974 | 0.011302 | 0.82175 | 1 |
| Esophagus_Muscularis | -0.01512 | -0.02929 | 0.015467 | 0.83591 | 1 |
| Heart_Atrial_Appendage | -0.01382 | -0.024296 | 0.011774 | 0.87974 | 1 |
| Muscle_Skeletal | -0.01131 | -0.021147 | 0.00869 | 0.9035 | 1 |
| Brain_Spinal_cord_cervical_c-1 | -0.01454 | -0.025641 | 0.010666 | 0.91357 | 1 |
| Heart_Left_Ventricle | -0.0161 | -0.026278 | 0.011336 | 0.92222 | 1 |
| Brain_Cerebellar_Hemisphere | -0.01503 | -0.029716 | 0.007577 | 0.97636 | 1 |
| Brain_Cerebellum | -0.01634 | -0.031895 | 0.007842 | 0.9814 | 1 |
| Brain_Amygdala | -0.02469 | -0.041071 | 0.009942 | 0.99349 | 1 |
| Brain_Caudate_basal_ganglia | -0.0252 | -0.042529 | 0.0099 | 0.99453 | 1 |
| Brain_Substantia_nigra | -0.02754 | -0.046378 | 0.010772 | 0.99471 | 1 |
| Brain_Cortex | -0.02332 | -0.041569 | 0.008975 | 0.99531 | 1 |
| Brain_Hippocampus | -0.02634 | -0.043503 | 0.010103 | 0.99543 | 1 |
| Brain_Frontal_Cortex_BA9 | -0.02263 | -0.040833 | 0.008631 | 0.99562 | 1 |
| Brain_Anterior_cingulate_cortex_BA24 | -0.02394 | -0.041346 | 0.009108 | 0.99571 | 1 |
| Brain_Hypothalamus | -0.02767 | -0.046515 | 0.010227 | 0.99659 | 1 |
| Brain_Nucleus_accumbens_basal_ganglia | -0.02612 | -0.044306 | 0.009556 | 0.99686 | 1 |
| Brain_Putamen_basal_ganglia | -0.02764 | -0.045966 | 0.010004 | 0.99713 | 1 |

GTEx: the Genotype-Tissue Expression project. MAGMA: Multi-marker Analysis of GenoMic Annotation. SD: standard deviation. SE: standard error.

**Supplemental Table 5** Significantly enriched tissues and cell types based on DEPICT analysis

| MeSH.term | name | MeSH.first.level.term | MeSH.second.level.term | Nominal.p.values | Bonferroni-corrected.p.values |
| --- | --- | --- | --- | --- | --- |
| A15.145.229.637.555 | Leukocytes Mononuclear | Hematologic and Immune Systems | Blood | 1.41E-07 | 2.95E-05 |
| A04.623.603 | Oropharynx | Respiratory System | Pharynx | 1.82E-07 | 3.80E-05 |
| A15.382.520.604.800 | Palatine Tonsil | Hematologic and Immune Systems | Immune System | 1.82E-07 | 3.80E-05 |
| A02.835.583.443.800.800 | Synovial Fluid | Musculoskeletal System | Skeleton | 1.26E-06 | 0.00026334 |
| A15.382.490.555.567.537 | Killer Cells Natural | Hematologic and Immune Systems | Immune System | 1.67E-06 | 0.00034903 |
| A15.382.490.555.567 | Lymphocytes | Hematologic and Immune Systems | Immune System | 2.33E-06 | 0.00048697 |
| A15.145.300 | Fetal Blood | Hematologic and Immune Systems | Blood | 3.69E-06 | 0.00077121 |
| A15.145.229 | Blood Cells | Hematologic and Immune Systems | Blood | 3.78E-06 | 0.00079002 |
| A11.118.637 | Leukocytes | Cells | Blood Cells | 1.00E-05 | 0.00209 |
| A15.145 | Blood | Hematologic and Immune Systems | Blood | 1.05E-05 | 0.0021945 |
| A11.118.637.555.567.569 | T Lymphocytes | Cells | Blood Cells | 4.68E-05 | 0.0097812 |
| A15.382.520.604.700 | Spleen | Hematologic and Immune Systems | Immune System | 5.78E-05 | 0.0120802 |
| A15.382 | Immune System | Hematologic and Immune Systems | Immune System | 8.14E-05 | 0.0170126 |
| A11.066 | Antigen Presenting Cells | Cells | Antigen-Presenting Cells | 8.15E-05 | 0.0170335 |
| A15.382.812.260 | Dendritic Cells | Hematologic and Immune Systems | Immune System | 8.15E-05 | 0.0170335 |
| A10.549 | Lymphoid Tissue | Tissues | Lymphoid Tissue | 9.14E-05 | 0.0191026 |
| A15.382.520 | Lymphatic System | Hematologic and Immune Systems | Immune System | 9.14E-05 | 0.0191026 |
| A02.835.232.043.300.710 | Tarsal Bones | Musculoskeletal System | Skeleton | 0.000124 | 0.025916 |
| A02.835.232.043.300 | Foot Bones | Musculoskeletal System | Skeleton | 0.000124 | 0.025916 |
| A02.835.232.043 | Bones of Lower Extremity | Musculoskeletal System | Skeleton | 0.000195 | 0.040755 |

MeSH: Medical Subject Headings

**Supplemental Table 6** Significantly enriched pathways based on MAGMA analysis

| Pathway | Number of genes | Beta | Beta SD | SE | P-values | Bonferroni-corrected p-values |
| --- | --- | --- | --- | --- | --- | --- |
| Curated_gene_sets:biocarta_th1th2_pathway | 16 | 2.1629 | 0.069586 | 0.26728 | 3.18E-16 | 4.92E-12 |
| GO_bp:go_regulation_of_immune_system_process | 1150 | 0.21742 | 0.05708 | 0.029871 | 1.78E-13 | 2.75E-09 |
| GO_bp:go_positive_regulation_of_immune_system_process | 813 | 0.25489 | 0.056926 | 0.035208 | 2.37E-13 | 3.67E-09 |
| Curated_gene_sets:pid_il27_pathway | 22 | 1.7079 | 0.06442 | 0.23781 | 3.62E-13 | 5.60E-09 |
| Curated_gene_sets:reactome_interleukin_35_signalling | 11 | 2.3834 | 0.063589 | 0.33384 | 4.94E-13 | 7.63E-09 |
| GO_bp:go_interleukin_35_mediated_signaling_pathway | 10 | 2.5366 | 0.06453 | 0.35535 | 4.96E-13 | 7.67E-09 |
| GO_bp:go_regulation_of_immune_response | 763 | 0.25152 | 0.054511 | 0.036198 | 1.93E-12 | 2.98E-08 |
| Curated_gene_sets:kegg_allograft_rejection | 20 | 1.6613 | 0.059747 | 0.24106 | 2.89E-12 | 4.46E-08 |
| GO_bp:go_t_cell_activation | 352 | 0.37307 | 0.055681 | 0.054164 | 2.96E-12 | 4.57E-08 |
| GO_bp:go_immune_response_regulating_cell_surface_receptor_signaling_pathway | 307 | 0.38598 | 0.053879 | 0.056222 | 3.46E-12 | 5.36E-08 |
| GO_bp:go_antigen_receptor_mediated_signaling_pathway | 185 | 0.48528 | 0.052797 | 0.070768 | 3.66E-12 | 5.66E-08 |
| GO_bp:go_positive_regulation_of_immune_response | 614 | 0.26896 | 0.052554 | 0.040221 | 1.18E-11 | 1.83E-07 |
| GO_bp:go_t_cell_receptor_signaling_pathway | 150 | 0.52348 | 0.051342 | 0.078356 | 1.24E-11 | 1.91E-07 |
| GO_bp:go_leukocyte_cell_cell_adhesion | 253 | 0.41387 | 0.05254 | 0.063129 | 2.86E-11 | 4.43E-07 |
| GO_bp:go_regulation_of_t_cell_activation | 245 | 0.41985 | 0.052463 | 0.064332 | 3.49E-11 | 5.40E-07 |
| GO_bp:go_cytokine_mediated_signaling_pathway | 584 | 0.27112 | 0.051719 | 0.041886 | 4.98E-11 | 7.69E-07 |
| GO_bp:go_immune_response_regulating_signaling_pathway | 437 | 0.30396 | 0.050406 | 0.047318 | 6.87E-11 | 1.06E-06 |
| Curated_gene_sets:kegg_t_cell_receptor_signaling_pathway | 90 | 0.6656 | 0.050666 | 0.1042 | 8.68E-11 | 1.34E-06 |
| GO_bp:go_regulation_of_lymphocyte_activation | 319 | 0.35923 | 0.051096 | 0.056934 | 1.44E-10 | 2.23E-06 |
| GO_bp:go_antigen_processing_and_presentation_of_endogenous_peptide_antigen | 8 | 2.3646 | 0.053807 | 0.37928 | 2.34E-10 | 3.61E-06 |
| Curated_gene_sets:pid_il12_2pathway | 49 | 0.85462 | 0.048065 | 0.14259 | 1.05E-09 | 1.62E-05 |
| GO_bp:go_activation_of_immune_response | 471 | 0.27032 | 0.046484 | 0.045408 | 1.35E-09 | 2.08E-05 |
| GO_bp:go_t_cell_differentiation | 185 | 0.43256 | 0.047062 | 0.074834 | 3.81E-09 | 5.89E-05 |
| Curated_gene_sets:biocarta_no2il12_pathway | 13 | 1.6811 | 0.048755 | 0.29165 | 4.20E-09 | 6.49E-05 |
| GO_bp:go_regulation_of_cell_activation | 418 | 0.28561 | 0.04635 | 0.049796 | 4.96E-09 | 7.66E-05 |
| GO_bp:go_lymphocyte_activation | 504 | 0.25735 | 0.045728 | 0.045162 | 6.18E-09 | 9.54E-05 |
| GO_bp:go_positive_regulation_of_cell_activation | 255 | 0.35758 | 0.04557 | 0.062972 | 6.94E-09 | 0.000107 |
| Curated_gene_sets:reactome_interleukin_23_signaling | 8 | 2.2953 | 0.05223 | 0.40614 | 8.12E-09 | 0.000125 |
| GO_bp:go_interleukin_23_mediated_signaling_pathway | 8 | 2.2953 | 0.05223 | 0.40614 | 8.12E-09 | 0.000125 |
| GO_bp:go_positive_regulation_of_lymphocyte_activation | 209 | 0.39332 | 0.045448 | 0.069824 | 9.03E-09 | 0.000139 |
| GO_bp:go_lymphocyte_differentiation | 263 | 0.35062 | 0.045366 | 0.062352 | 9.56E-09 | 0.000148 |
| GO_bp:go_positive_regulation_of_leukocyte_cell_cell_adhesion | 166 | 0.43037 | 0.044381 | 0.076961 | 1.14E-08 | 0.000177 |
| Curated_gene_sets:kegg_autoimmune_thyroid_disease | 28 | 1.1638 | 0.049513 | 0.20893 | 1.30E-08 | 0.0002 |
| GO_bp:go_t_cell_mediated_immunity | 82 | 0.64063 | 0.046559 | 0.11716 | 2.32E-08 | 0.000358 |
| GO_bp:go_positive_regulation_of_leukocyte_mediated_immunity | 110 | 0.52923 | 0.044508 | 0.099032 | 4.62E-08 | 0.000713 |
| GO_cc:go_side_of_membrane | 388 | 0.27566 | 0.043142 | 0.051674 | 4.87E-08 | 0.000751 |
| GO_bp:go_regulation_of_cell_cell_adhesion | 305 | 0.30638 | 0.042631 | 0.057597 | 5.29E-08 | 0.000816 |
| GO_bp:go_positive_regulation_of_cell_cell_adhesion | 196 | 0.37495 | 0.041974 | 0.071098 | 6.79E-08 | 0.001047 |
| GO_bp:go_antigen_processing_and_presentation_of_endogenous_antigen | 14 | 1.6258 | 0.04893 | 0.30843 | 6.89E-08 | 0.001062 |
| GO_bp:go_alpha_beta_t_cell_activation | 107 | 0.51638 | 0.042835 | 0.099296 | 1.01E-07 | 0.001557 |
| Curated_gene_sets:biocarta_asbcell_pathway | 8 | 1.8461 | 0.042008 | 0.36437 | 2.05E-07 | 0.003168 |
| GO_bp:go_regulation_of_lymphocyte_differentiation | 131 | 0.44289 | 0.040619 | 0.087542 | 2.13E-07 | 0.003291 |
| GO_bp:go_regulation_of_leukocyte_proliferation | 171 | 0.39606 | 0.041447 | 0.078366 | 2.19E-07 | 0.00338 |
| GO_bp:go_response_to_cytokine | 902 | 0.16985 | 0.039832 | 0.033964 | 2.89E-07 | 0.004457 |
| GO_bp:go_fc_receptor_signaling_pathway | 153 | 0.38535 | 0.038167 | 0.078394 | 4.48E-07 | 0.006908 |
| GO_bp:go_regulation_of_leukocyte_mediated_immunity | 159 | 0.39767 | 0.040144 | 0.081304 | 5.07E-07 | 0.007817 |
| Curated_gene_sets:pid_cd8_tcr_pathway | 46 | 0.6996 | 0.038127 | 0.14404 | 6.03E-07 | 0.009296 |
| Curated_gene_sets:pid_il23_pathway | 32 | 0.88426 | 0.040212 | 0.183 | 6.83E-07 | 0.01053 |
| Curated_gene_sets:kegg_jak_stat_signaling_pathway | 115 | 0.47632 | 0.040951 | 0.098609 | 6.89E-07 | 0.010621 |
| Curated_gene_sets:biocarta_il12_pathway | 17 | 1.2637 | 0.041906 | 0.26272 | 7.62E-07 | 0.011751 |
| GO_mf:go_cytokine_receptor_activity | 75 | 0.56208 | 0.039077 | 0.11686 | 7.63E-07 | 0.011768 |
| GO_bp:go_positive_regulation_of_cytokine_production | 349 | 0.26234 | 0.038991 | 0.054657 | 8.03E-07 | 0.01237 |
| GO_bp:go_leukocyte_differentiation | 377 | 0.24922 | 0.038463 | 0.051957 | 8.15E-07 | 0.012562 |
| GO_bp:go_leukocyte_proliferation | 226 | 0.32302 | 0.038791 | 0.067402 | 8.33E-07 | 0.012831 |
| GO_bp:go_t_cell_antigen_processing_and_presentation | 3 | 2.5227 | 0.035159 | 0.52691 | 8.52E-07 | 0.013135 |
| GO_cc:go_external_side_of_plasma_membrane | 242 | 0.30946 | 0.038436 | 0.064751 | 8.89E-07 | 0.013698 |
| GO_bp:go_regulation_of_alpha_beta_t_cell_activation | 74 | 0.56513 | 0.039028 | 0.1188 | 9.92E-07 | 0.01529 |
| GO_bp:go_regulation_of_t_cell_differentiation | 111 | 0.45166 | 0.038156 | 0.095111 | 1.03E-06 | 0.015924 |
| GO_bp:go_cell_activation | 1058 | 0.14665 | 0.037048 | 0.031111 | 1.23E-06 | 0.018915 |
| Curated_gene_sets:pid_il12_stat4_pathway | 30 | 0.86184 | 0.03795 | 0.18319 | 1.28E-06 | 0.019783 |
| GO_bp:go_regulation_of_dendritic_cell_chemotaxis | 6 | 2.3295 | 0.045909 | 0.49692 | 1.39E-06 | 0.021476 |
| GO_bp:go_immune_effector_process | 909 | 0.15611 | 0.036744 | 0.033405 | 1.50E-06 | 0.023057 |
| Curated_gene_sets:reactome_death_receptor_signalling | 108 | 0.41196 | 0.034331 | 0.088314 | 1.56E-06 | 0.02403 |
| GO_bp:go_positive_regulation_of_lymphocyte_mediated_immunity | 86 | 0.54536 | 0.040585 | 0.11768 | 1.81E-06 | 0.027856 |
| GO_bp:go_regulation_of_t_cell_mediated_immunity | 54 | 0.66343 | 0.039164 | 0.14353 | 1.92E-06 | 0.029518 |
| Curated_gene_sets:reactome_fc_epsilon_receptor_fceri_signaling | 109 | 0.4184 | 0.035028 | 0.090523 | 1.92E-06 | 0.029534 |
| Curated_gene_sets:galindo_immune_response_to_enterotoxin | 54 | 0.59682 | 0.035231 | 0.13018 | 2.30E-06 | 0.03535 |
| GO_bp:go_regulation_of_cell_killing | 73 | 0.55941 | 0.038372 | 0.12237 | 2.44E-06 | 0.03761 |
| GO_bp:go_regulation_of_antigen_receptor_mediated_signaling_pathway | 51 | 0.62948 | 0.036116 | 0.13773 | 2.46E-06 | 0.037803 |
| GO_bp:go_regulation_of_lymphocyte_mediated_immunity | 119 | 0.44396 | 0.038823 | 0.097245 | 2.51E-06 | 0.038715 |
| GO_bp:go_positive_regulation_of_interferon_gamma_production | 49 | 0.68684 | 0.038629 | 0.1506 | 2.57E-06 | 0.039611 |
| GO_bp:go_negative_regulation_of_leukocyte_cell_cell_adhesion | 91 | 0.48249 | 0.036929 | 0.10652 | 2.98E-06 | 0.045914 |
| Curated_gene_sets:reactome_signaling_by_the_b_cell_receptor_bcr | 89 | 0.44425 | 0.033629 | 0.098156 | 3.03E-06 | 0.046673 |

MAGMA: Multi-marker Analysis of GenoMic Annotation. SD: standard deviation. SE: standard error.

**Supplemental Table 7** Colocalization analysis with eQTLs from blood, skin, lung, liver and immune cell types.

| Tissue or cell type | GWAS lead SNP | CHR | BP | PP4 | Gene | Direction |
| --- | --- | --- | --- | --- | --- | --- |
| blood | rs27524 | 5 | 96101944 | 87% | *ERAP2* | Concordant |
| blood | rs17338998 | 7 | 128618559 | 98% | *TNPO3* | Opposite |
| blood | rs4743150 | 9 | 100740124 | 99% | *ANP32B* | Concordant |
| blood | rs10083496 | 14 | 105402786 | 95% | *PLD4* | Concordant |
| blood | rs10083496 | 14 | 105402786 | 93% | *AKT1* | Concordant |
| blood | rs2305743 | 19 | 18193191 | 98% | *IL12RB1* | Concordant |
| Skin | rs17338998 | 7 | 128618559 | 100% | *IRF5* | Concordant |
| Skin | rs4810485 | 20 | 44747947 | 99% | *CD40* | Opposite |
| Lung | rs17338998 | 7 | 128618559 | 100% | *IRF5* | Concordant |
| Lung | rs551125 | 12 | 121203427 | 60% | *SPPL3* | Opposite |
| Lung | rs10083496 | 14 | 105402786 | 66% | *PLD4* | Concordant |
| Lung | rs10083496 | 14 | 105402786 | 94% | *LINC00638* | Opposite |
| Lung | rs2305743 | 19 | 18193191 | 97% | *IL12RB1* | Concordant |
| Lung | rs4810485 | 20 | 44747947 | 98% | *CD40* | Opposite |
| Liver | rs27524 | 5 | 96101944 | 98% | *ERAP1* | Opposite |
| Liver | rs17338998 | 7 | 128618559 | 100% | *IRF5* | Concordant |
| B cells | rs17338998 | 7 | 128618559 | 89% | *TNPO3* | Opposite |
| CD4+ T cells | rs17338998 | 7 | 128618559 | 72% | *TNPO3* | Opposite |
| CD4+ T cells | rs2305743 | 19 | 18193191 | 82% | *IL12RB1* | Concordant |
| CD8+ T cells | rs17338998 | 7 | 128618559 | 55% | *TNPO3* | Opposite |
| CD8+ T cells | rs2305743 | 19 | 18193191 | 53% | *IL12RB1* | Concordant |
| Monocytes | rs17338998 | 7 | 128618559 | 100% | *TNPO3* | Opposite |
| Neutrophils | rs17338998 | 7 | 128618559 | 100% | *TNPO3* | Opposite |

PP4: the posterior probability of hypothesis 4 that there is at least one variant causal to both traits

**Supplemental Table 8** Meta-phenome-wide association study (Meta-PheWAS) for the genome-wide polygenic

score for the SSc-PBC meta-analysis statistics across the eMERGE-III, All of Us and UKBB datasets.

| Phecode | Phenotype description | Phenotype group | Beta | OR | SE | P-values | N cases | N controls |
| --- | --- | --- | --- | --- | --- | --- | --- | --- |
| 571.6 | Primary biliary cirrhosis | digestive | 1.10 | 3.01 | 0.06 | 5.55E-86 | 636 | 575421 |
| 709 | Diffuse diseases of connective tissue | dermatologic | 0.43 | 1.53 | 0.02 | 1.20E-72 | 4789 | 565594 |
| 695.4 | Lupus (localized and systemic) | dermatologic | 0.55 | 1.74 | 0.03 | 4.50E-71 | 3059 | 566042 |
| 695.42 | Systemic lupus erythematosus | dermatologic | 0.56 | 1.76 | 0.03 | 3.34E-64 | 2663 | 566042 |
| 714 | Rheumatoid arthritis and other inflammatory polyarthropathies | musculoskeletal | 0.21 | 1.23 | 0.01 | 3.66E-58 | 13103 | 565992 |
| 714.1 | Rheumatoid arthritis | musculoskeletal | 0.22 | 1.24 | 0.01 | 2.16E-55 | 10939 | 565992 |
| 244.4 | Hypothyroidism NOS | endocrine/metabolic | 0.12 | 1.13 | 0.01 | 2.82E-55 | 39487 | 557418 |
| 244 | Hypothyroidism | endocrine/metabolic | 0.12 | 1.12 | 0.01 | 4.42E-55 | 43752 | 557418 |
| 709.2 | Sicca syndrome | dermatologic | 0.46 | 1.58 | 0.03 | 9.46E-43 | 2209 | 565594 |
| 709.3 | Systemic sclerosis | dermatologic | 0.58 | 1.78 | 0.05 | 9.19E-30 | 969 | 565594 |
| 443.1 | Raynaud's syndrome | circulatory system | 0.26 | 1.30 | 0.02 | 2.87E-26 | 3634 | 578147 |
| 695.41 | Cutaneous lupus erythematosus | dermatologic | 0.59 | 1.80 | 0.06 | 5.70E-25 | 834 | 566042 |
| 695 | Erythematous conditions | dermatologic | 0.14 | 1.15 | 0.01 | 4.42E-24 | 16263 | 569412 |
| 335 | Multiple sclerosis | neurological | 0.26 | 1.30 | 0.03 | 1.15E-23 | 3146 | 562788 |
| 443 | Peripheral vascular disease | circulatory system | 0.13 | 1.14 | 0.01 | 3.36E-23 | 15500 | 578147 |
| 571 | Chronic liver disease and cirrhosis | digestive | 0.13 | 1.14 | 0.01 | 5.02E-23 | 14632 | 575421 |
| 555 | Inflammatory bowel disease and other gastroenteritis and colitis | digestive | 0.13 | 1.14 | 0.01 | 3.23E-21 | 10058 | 473644 |
| 558 | Noninfectious gastroenteritis | digestive | 0.07 | 1.07 | 0.01 | 8.75E-18 | 27701 | 473644 |
| 285 | Other anemias | hematopoietic | 0.06 | 1.06 | 0.01 | 1.42E-15 | 52370 | 535342 |
| 418 | Nonspecific chest pain | circulatory system | 0.04 | 1.04 | 0.01 | 8.72E-15 | 88052 | 513185 |
| 785 | Abdominal pain | symptoms | 0.04 | 1.04 | 0.01 | 1.66E-14 | 102110 | 502851 |
| 296.2 | Depression | mental disorders | 0.05 | 1.06 | 0.01 | 4.44E-14 | 63016 | 504498 |
| 555.1 | Regional enteritis | digestive | 0.16 | 1.17 | 0.02 | 1.20E-13 | 5005 | 473644 |
| 550.2 | Diaphragmatic hernia | digestive | 0.05 | 1.05 | 0.01 | 1.27E-13 | 42888 | 538454 |
| 401 | Hypertension | circulatory system | 0.03 | 1.03 | 0.00 | 2.11E-13 | 198799 | 422498 |
| 296 | Mood disorders | mental disorders | 0.05 | 1.05 | 0.01 | 3.36E-13 | 67599 | 504498 |
| 401.1 | Essential hypertension | circulatory system | 0.03 | 1.03 | 0.00 | 4.10E-13 | 196898 | 422498 |
| 535 | Gastritis and duodenitis | digestive | 0.05 | 1.05 | 0.01 | 5.08E-13 | 48766 | 547049 |
| 709.7 | Unspecified diffuse connective tissue disease | dermatologic | 0.35 | 1.42 | 0.05 | 5.82E-13 | 1212 | 565594 |
| 480 | Pneumonia | respiratory | 0.06 | 1.06 | 0.01 | 1.52E-12 | 31620 | 564372 |
| 555.2 | Ulcerative colitis | digestive | 0.12 | 1.13 | 0.02 | 1.69E-12 | 6510 | 473644 |
| 530 | Diseases of esophagus | digestive | 0.04 | 1.04 | 0.01 | 4.19E-12 | 95046 | 496894 |
| 789 | Nausea and vomiting | symptoms | 0.05 | 1.05 | 0.01 | 1.37E-11 | 42695 | 566268 |
| 696.4 | Psoriasis | dermatologic | 0.12 | 1.13 | 0.02 | 1.84E-11 | 7403 | 558172 |
| 573.7 | Abnormal results of function study of liver | digestive | 0.11 | 1.11 | 0.02 | 2.36E-11 | 7331 | 575421 |
| 512 | Other symptoms of respiratory system | respiratory | 0.05 | 1.05 | 0.01 | 6.82E-11 | 82514 | 518532 |
| 504 | Other alveolar and parietoalveolar pneumonopathy | respiratory | 0.17 | 1.19 | 0.03 | 7.47E-11 | 2917 | 568291 |
| 530.1 | Esophagitis, GERD and related diseases | digestive | 0.03 | 1.04 | 0.01 | 7.72E-11 | 89042 | 496894 |
| 535.9 | Gastritis and duodenitis, NOS | digestive | 0.05 | 1.06 | 0.01 | 9.31E-11 | 25157 | 547049 |
| 300 | Anxiety disorders | mental disorders | 0.05 | 1.05 | 0.01 | 1.01E-10 | 51294 | 504498 |
| 571.51 | Cirrhosis of liver without mention of alcohol | digestive | 0.18 | 1.20 | 0.03 | 1.66E-10 | 3195 | 575421 |
| 696 | Psoriasis and related disorders | dermatologic | 0.11 | 1.12 | 0.02 | 1.95E-10 | 7957 | 558172 |
| 281.11 | Pernicious anemia | hematopoietic | 0.21 | 1.23 | 0.03 | 2.21E-10 | 1865 | 535342 |
| 317 | Alcohol-related disorders | mental disorders | 0.07 | 1.08 | 0.01 | 3.53E-10 | 17178 | 564822 |
| 317.1 | Alcoholism | mental disorders | 0.08 | 1.08 | 0.01 | 4.64E-10 | 13559 | 564822 |
| 480.1 | Bacterial pneumonia | respiratory | 0.07 | 1.08 | 0.01 | 4.78E-10 | 12735 | 564372 |
| 446 | Polyarteritis nodosa and allied conditions | circulatory system | 0.17 | 1.19 | 0.03 | 6.72E-10 | 3053 | 578147 |
| 300.1 | Anxiety disorder | mental disorders | 0.06 | 1.06 | 0.01 | 8.83E-10 | 41663 | 504498 |
| 564 | Functional digestive disorders | digestive | 0.04 | 1.04 | 0.01 | 1.09E-09 | 37198 | 473644 |
| 571.5 | Other chronic nonalcoholic liver disease | digestive | 0.09 | 1.09 | 0.01 | 1.09E-09 | 12682 | 575421 |
| 561 | Symptoms involving digestive system | digestive | 0.05 | 1.06 | 0.01 | 1.76E-09 | 22416 | 473644 |
| 279 | Disorders involving the immune mechanism | endocrine/metabolic | 0.12 | 1.13 | 0.02 | 1.84E-09 | 8791 | 613761 |
| 564.9 | Personal history of diseases of digestive system | digestive | 0.05 | 1.05 | 0.01 | 3.01E-09 | 21543 | 388186 |
| 714.2 | Juvenile rheumatoid arthritis | musculoskeletal | 0.39 | 1.47 | 0.07 | 3.77E-09 | 742 | 565992 |
| 279.7 | Other immunological findings | endocrine/metabolic | 0.24 | 1.27 | 0.04 | 3.98E-09 | 1644 | 500215 |
| 687.1 | Rash and other nonspecific skin eruption | dermatologic | 0.08 | 1.08 | 0.01 | 6.29E-09 | 16788 | 560555 |
| 480.11 | Pneumococcal pneumonia | respiratory | 0.07 | 1.08 | 0.01 | 6.70E-09 | 9760 | 564372 |
| 585 | Renal failure | genitourinary | 0.05 | 1.05 | 0.01 | 7.18E-09 | 40613 | 556055 |
| 446.9 | Arteritis NOS | circulatory system | 0.25 | 1.28 | 0.04 | 8.17E-09 | 1362 | 578147 |
| 172 | Skin cancer | neoplasms | -0.05 | 0.95 | 0.01 | 1.23E-08 | 32624 | 582525 |
| 507 | Pleurisy; pleural effusion | respiratory | 0.06 | 1.06 | 0.01 | 1.29E-08 | 22165 | 568291 |
| 279.2 | Autoimmune disease NEC | endocrine/metabolic | 0.30 | 1.35 | 0.05 | 1.68E-08 | 1254 | 200940 |
| 504.1 | Idiopathic fibrosing alveolitis | respiratory | 0.17 | 1.19 | 0.03 | 2.00E-08 | 1982 | 568291 |
| 696.41 | Psoriasis vulgaris | dermatologic | 0.11 | 1.12 | 0.02 | 2.64E-08 | 6060 | 558172 |
| 696.42 | Psoriatic arthropathy | dermatologic | 0.18 | 1.20 | 0.03 | 2.72E-08 | 1952 | 558172 |
| 276 | Disorders of fluid, electrolyte, and acid-base balance | endocrine/metabolic | 0.05 | 1.05 | 0.01 | 2.96E-08 | 42816 | 565879 |
| 172.2 | Other non-epithelial cancer of skin | neoplasms | -0.05 | 0.95 | 0.01 | 3.80E-08 | 27150 | 582525 |
| 414 | Other forms of chronic heart disease | circulatory system | 0.04 | 1.04 | 0.01 | 5.57E-08 | 38573 | 540049 |
| 512.9 | Other dyspnea | respiratory | 0.07 | 1.07 | 0.01 | 6.51E-08 | 28639 | 518532 |
| 496 | Chronic airway obstruction | respiratory | 0.05 | 1.05 | 0.01 | 9.21E-08 | 30997 | 523795 |
| 585.1 | Acute renal failure | genitourinary | 0.06 | 1.06 | 0.01 | 1.49E-07 | 19894 | 556055 |
| 591 | Urinary tract infection | genitourinary | 0.04 | 1.04 | 0.01 | 1.62E-07 | 41881 | 531251 |
| 411.8 | Other chronic ischemic heart disease, unspecified | circulatory system | 0.04 | 1.04 | 0.01 | 2.38E-07 | 28375 | 540049 |
| 550 | Abdominal hernia | digestive | 0.03 | 1.03 | 0.01 | 2.48E-07 | 79607 | 538454 |
| 562.1 | Diverticulosis | digestive | 0.03 | 1.03 | 0.01 | 2.73E-07 | 52068 | 473644 |
| 562 | Diverticulosis and diverticulitis | digestive | 0.03 | 1.03 | 0.01 | 3.78E-07 | 54593 | 473644 |
| 454 | Varicose veins | circulatory system | 0.05 | 1.05 | 0.01 | 4.07E-07 | 21919 | 515264 |
| 411 | Ischemic Heart Disease | circulatory system | 0.03 | 1.03 | 0.01 | 4.42E-07 | 70128 | 540049 |
| 709.6 | Other specified diffuse diseases of connective tissue | dermatologic | 0.40 | 1.49 | 0.08 | 4.51E-07 | 444 | 565594 |
| 454.1 | Varicose veins of lower extremity | circulatory system | 0.05 | 1.05 | 0.01 | 5.41E-07 | 20400 | 515264 |
| 578 | Gastrointestinal hemorrhage | digestive | 0.03 | 1.03 | 0.01 | 5.78E-07 | 44071 | 554087 |
| 428.2 | Heart failure NOS | circulatory system | 0.07 | 1.07 | 0.01 | 5.95E-07 | 12571 | 582024 |
| 428 | Congestive heart failure; nonhypertensive | circulatory system | 0.05 | 1.05 | 0.01 | 6.14E-07 | 27484 | 582024 |
| 557.1 | Celiac disease | digestive | 0.12 | 1.12 | 0.02 | 6.42E-07 | 3196 | 473644 |
| 415 | Pulmonary heart disease | circulatory system | 0.06 | 1.06 | 0.01 | 6.62E-07 | 15237 | 593617 |
| 519 | Other diseases of respiratory system, not elsewhere classified | respiratory | 0.05 | 1.05 | 0.01 | 6.66E-07 | 19676 | 599628 |
| 443.9 | Peripheral vascular disease, unspecified | circulatory system | 0.08 | 1.08 | 0.02 | 6.66E-07 | 10969 | 578147 |
| 573.2 | Liver replaced by transplant | digestive | 0.30 | 1.35 | 0.06 | 1.00E-06 | 724 | 575421 |
| 281.1 | Megaloblastic anemia | hematopoietic | 0.12 | 1.13 | 0.03 | 1.01E-06 | 3471 | 535342 |
| 414.2 | ASCVD | circulatory system | 0.04 | 1.04 | 0.01 | 1.31E-06 | 32422 | 540049 |
| 287 | Purpura and other hemorrhagic conditions | hematopoietic | 0.08 | 1.08 | 0.02 | 1.47E-06 | 9633 | 595630 |
| 770 | Myalgia and myositis unspecified | symptoms | 0.07 | 1.07 | 0.01 | 1.88E-06 | 16904 | 601052 |
| 276.1 | Electrolyte imbalance | endocrine/metabolic | 0.05 | 1.05 | 0.01 | 1.88E-06 | 29148 | 565879 |
| 681 | Superficial cellulitis and abscess | dermatologic | 0.04 | 1.04 | 0.01 | 1.96E-06 | 34519 | 569376 |
| 495.11 | Chronic obstructive asthma with exacerbation | respiratory | 0.08 | 1.09 | 0.02 | 2.06E-06 | 5730 | 523795 |
| 519.8 | Other diseases of respiratory system, NEC | respiratory | 0.05 | 1.05 | 0.01 | 2.25E-06 | 13890 | 599628 |
| 280 | Iron deficiency anemias | hematopoietic | 0.04 | 1.04 | 0.01 | 3.38E-06 | 26788 | 535342 |
| 578.8 | Hemorrhage of rectum and anus | digestive | 0.04 | 1.04 | 0.01 | 3.96E-06 | 21736 | 554087 |
| 281 | Other deficiency anemia | hematopoietic | 0.11 | 1.11 | 0.02 | 4.24E-06 | 4746 | 535342 |
| 717 | Polymyalgia Rheumatica | musculoskeletal | 0.12 | 1.12 | 0.03 | 4.33E-06 | 3113 | 628735 |
| 287.31 | Primary thrombocytopenia | hematopoietic | 0.18 | 1.20 | 0.04 | 4.64E-06 | 1419 | 595630 |
| 411.3 | Angina pectoris | circulatory system | 0.04 | 1.04 | 0.01 | 5.92E-06 | 28777 | 540049 |
| 216 | Benign neoplasm of skin | neoplasms | -0.04 | 0.96 | 0.01 | 7.23E-06 | 39632 | 577037 |
| 261 | Vitamin deficiency | endocrine/metabolic | 0.05 | 1.05 | 0.01 | 7.65E-06 | 26745 | 566979 |
| 245.2 | Chronic thyroiditis | endocrine/metabolic | 0.16 | 1.17 | 0.04 | 7.98E-06 | 2437 | 557418 |
| 960 | Poisoning by antibiotics | injuries & poisonings | 0.03 | 1.04 | 0.01 | 8.72E-06 | 27906 | 448582 |
| 245.21 | Chronic lymphocytic thyroiditis | endocrine/metabolic | 0.16 | 1.18 | 0.04 | 8.73E-06 | 2324 | 557418 |
| 716.1 | Unspecified polyarthropathy or polyarthritis | musculoskeletal | 0.19 | 1.20 | 0.04 | 9.05E-06 | 1334 | 567671 |
| 716 | Other arthropathies | musculoskeletal | 0.04 | 1.04 | 0.01 | 9.41E-06 | 30981 | 567671 |
| 783 | Fever of unknown origin | symptoms | 0.05 | 1.05 | 0.01 | 9.47E-06 | 23295 | 587991 |
| 709.4 | Polymyositis | dermatologic | 0.47 | 1.61 | 0.11 | 9.51E-06 | 233 | 565594 |
| 530.11 | GERD | digestive | 0.03 | 1.03 | 0.01 | 9.54E-06 | 60898 | 496894 |
| 509 | Respiratory failure, insufficiency, arrest | respiratory | 0.06 | 1.06 | 0.01 | 1.01E-05 | 14110 | 568291 |
| 278 | Overweight, obesity and other hyperalimentation | endocrine/metabolic | 0.03 | 1.03 | 0.01 | 1.05E-05 | 61107 | 549950 |
| 743.1 | Osteoporosis | musculoskeletal | 0.04 | 1.04 | 0.01 | 1.07E-05 | 28532 | 573225 |
| 580.31 | Nephritis and nephropathy in diseases classified elsewhere | genitourinary | 0.18 | 1.20 | 0.04 | 1.07E-05 | 1829 | 556055 |
| 535.6 | Duodenitis | digestive | 0.05 | 1.05 | 0.01 | 1.11E-05 | 10773 | 547049 |
| 411.4 | Coronary atherosclerosis | circulatory system | 0.04 | 1.04 | 0.01 | 1.30E-05 | 38772 | 540049 |
| 242 | Thyrotoxicosis with or without goiter | endocrine/metabolic | 0.08 | 1.09 | 0.02 | 1.35E-05 | 6471 | 557418 |
| 580.3 | Nephritis and nephropathy without mention of glomerulonephritis | genitourinary | 0.13 | 1.14 | 0.03 | 1.41E-05 | 3568 | 556055 |
| 509.8 | Dependence on respirator [Ventilator] or supplemental oxygen | respiratory | 0.16 | 1.17 | 0.04 | 1.48E-05 | 1789 | 568291 |
| 512.7 | Shortness of breath | respiratory | 0.06 | 1.06 | 0.01 | 1.52E-05 | 26845 | 110760 |
| 850 | Hemorrhage or hematoma complicating a procedure | injuries & poisonings | 0.06 | 1.06 | 0.01 | 1.53E-05 | 10115 | 591000 |
| 527.2 | Sialoadenitis | digestive | 0.19 | 1.21 | 0.04 | 1.53E-05 | 1172 | 605007 |
| 280.1 | Iron deficiency anemias, unspecified or not due to blood loss | hematopoietic | 0.04 | 1.04 | 0.01 | 1.57E-05 | 24000 | 535342 |
| 278.11 | Morbid obesity | endocrine/metabolic | 0.07 | 1.08 | 0.02 | 1.75E-05 | 14090 | 549950 |
| 939 | Atopic/contact dermatitis due to other or unspecified | dermatologic | 0.05 | 1.05 | 0.01 | 1.90E-05 | 28306 | 552695 |
| 455 | Hemorrhoids | circulatory system | 0.03 | 1.03 | 0.01 | 2.08E-05 | 54450 | 515264 |
| 496.3 | Bronchiectasis | respiratory | 0.09 | 1.09 | 0.02 | 2.16E-05 | 4778 | 523795 |
| 506 | Empyema and pneumothorax | respiratory | 0.11 | 1.11 | 0.03 | 2.33E-05 | 3919 | 568291 |
| 420.22 | Chronic pericarditis | circulatory system | 0.33 | 1.39 | 0.08 | 2.39E-05 | 442 | 608572 |
| 287.3 | Thrombocytopenia | hematopoietic | 0.07 | 1.08 | 0.02 | 2.41E-05 | 8436 | 595630 |
| 283.1 | Autoimmune hemolytic anemias | hematopoietic | 0.35 | 1.42 | 0.08 | 2.44E-05 | 300 | 535342 |
| 702.1 | Actinic keratosis | dermatologic | -0.06 | 0.94 | 0.01 | 2.49E-05 | 17311 | 585064 |

eMERGE: Electronic Medical Records and Genomics. UKBB: UK Biobank. OR: odds ratio. SE: standard error.

**Supplemental Table 9** Genes prioritized by MAGMA

| GENE | CHR | START | STOP | NSNPS | NPARAM | N | Z | P | SYMBOL | FDR Q_VALUE |
| --- | --- | --- | --- | --- | --- | --- | --- | --- | --- | --- |
| ENSG00000215912 | 1 | 2567415 | 2718286 | 2 | 1 | 52289 | 3.6623 | 1.25E-04 | TTC34 | 1.16E-02 |
| ENSG00000049239 | 1 | 9294834 | 9331396 | 29 | 3 | 52289 | 3.5775 | 1.73E-04 | H6PD | 1.46E-02 |
| ENSG00000001461 | 1 | 24742284 | 24799466 | 85 | 7 | 52289 | 3.1799 | 7.37E-04 | NIPAL3 | 4.05E-02 |
| ENSG00000168389 | 1 | 40420802 | 40435638 | 1 | 1 | 52289 | 3.2134 | 6.56E-04 | MFSD2A | 3.68E-02 |
| ENSG00000127124 | 1 | 41972036 | 42501596 | 566 | 31 | 52289 | 3.1428 | 8.37E-04 | HIVEP3 | 4.48E-02 |
| ENSG00000081985 | 1 | 67773047 | 67862583 | 23 | 5 | 52289 | 7.6975 | 6.94E-15 | IL12RB2 | 2.14E-11 |
| ENSG00000142864 | 1 | 67873493 | 67896098 | 2 | 1 | 52289 | 5.2819 | 6.39E-08 | SERBP1 | 2.15E-05 |
| ENSG00000162694 | 1 | 101337943 | 101361554 | 16 | 4 | 52289 | 3.4407 | 2.90E-04 | EXTL2 | 2.06E-02 |
| ENSG00000198758 | 1 | 110292702 | 110306649 | 2 | 1 | 52289 | 3.3645 | 3.83E-04 | EPS8L3 | 2.49E-02 |
| ENSG00000116815 | 1 | 117057157 | 117113661 | 14 | 1 | 52289 | 5.3555 | 4.27E-08 | CD58 | 1.50E-05 |
| ENSG00000169291 | 1 | 154442248 | 154474589 | 5 | 2 | 52289 | 3.2076 | 6.69E-04 | SHE | 3.73E-02 |
| ENSG00000160856 | 1 | 157644111 | 157670647 | 25 | 4 | 52289 | 3.7997 | 7.24E-05 | FCRL3 | 7.82E-03 |
| ENSG00000000460 | 1 | 169631245 | 169823221 | 543 | 18 | 52289 | 3.4696 | 2.61E-04 | C1orf112 | 1.95E-02 |
| ENSG00000188404 | 1 | 169659808 | 169680839 | 73 | 10 | 52289 | 3.1763 | 7.46E-04 | SELL | 4.07E-02 |
| ENSG00000007908 | 1 | 169691781 | 169733846 | 162 | 5 | 52289 | 3.3809 | 3.61E-04 | SELE | 2.37E-02 |
| ENSG00000171806 | 1 | 169761670 | 169764107 | 4 | 2 | 52289 | 3.7241 | 9.80E-05 | METTL18 | 9.64E-03 |
| ENSG00000000457 | 1 | 169818772 | 169863408 | 107 | 9 | 52289 | 3.5366 | 2.03E-04 | SCYL3 | 1.65E-02 |
| ENSG00000134376 | 1 | 197170592 | 197447585 | 102 | 14 | 52289 | 4.0909 | 2.15E-05 | CRB1 | 2.81E-03 |
| ENSG00000213047 | 1 | 197473878 | 197744826 | 13 | 3 | 52289 | 4.6163 | 1.95E-06 | DENND1B | 3.68E-04 |
| ENSG00000163362 | 1 | 200860176 | 200884863 | 8 | 1 | 52289 | 4.9697 | 3.35E-07 | C1orf106 | 8.78E-05 |
| ENSG00000136634 | 1 | 206940947 | 206945839 | 5 | 2 | 52289 | 5.1245 | 1.49E-07 | IL10 | 4.19E-05 |
| ENSG00000162892 | 1 | 207070788 | 207077484 | 10 | 2 | 52289 | 3.518 | 2.17E-04 | IL24 | 1.72E-02 |
| ENSG00000143748 | 1 | 224415036 | 224518089 | 98 | 3 | 52289 | 3.3527 | 4.00E-04 | NVL | 2.57E-02 |
| ENSG00000177800 | 1 | 229385383 | 229387557 | 2 | 1 | 52289 | 3.442 | 2.89E-04 | TMEM78 | 2.06E-02 |
| ENSG00000119772 | 2 | 25455845 | 25565459 | 22 | 5 | 52289 | 5.2995 | 5.81E-08 | DNMT3A | 1.99E-05 |
| ENSG00000143889 | 2 | 38789120 | 38830728 | 54 | 5 | 52289 | 3.4584 | 2.72E-04 | HNRNPLL | 1.99E-02 |
| ENSG00000125618 | 2 | 113973574 | 114036527 | 108 | 14 | 52289 | 3.2163 | 6.49E-04 | PAX8 | 3.67E-02 |
| ENSG00000198130 | 2 | 191054461 | 191208919 | 142 | 9 | 52289 | 3.4664 | 2.64E-04 | HIBCH | 1.95E-02 |
| ENSG00000151690 | 2 | 191273081 | 191373931 | 129 | 14 | 52289 | 4.8714 | 5.54E-07 | MFSD6 | 1.27E-04 |
| ENSG00000189362 | 2 | 191369068 | 191399448 | 33 | 5 | 52289 | 3.4305 | 3.01E-04 | TMEM194B | 2.10E-02 |
| ENSG00000138386 | 2 | 191511472 | 191557492 | 27 | 3 | 52289 | 4.3538 | 6.69E-06 | NAB1 | 1.03E-03 |
| ENSG00000138378 | 2 | 191894302 | 192016322 | 38 | 9 | 52289 | 7.8821 | 1.61E-15 | STAT4 | 9.72E-12 |
| ENSG00000115020 | 2 | 209130991 | 209223475 | 183 | 8 | 52289 | 3.4124 | 3.22E-04 | PIKFYVE | 2.19E-02 |
| ENSG00000154822 | 3 | 16844159 | 17132086 | 432 | 32 | 52289 | 5.7435 | 4.64E-09 | PLCL2 | 2.32E-06 |
| ENSG00000168329 | 3 | 39304985 | 39323226 | 7 | 3 | 52289 | 3.2367 | 6.05E-04 | CX3CR1 | 3.46E-02 |
| ENSG00000163687 | 3 | 58177984 | 58200424 | 19 | 6 | 52289 | 4.0553 | 2.50E-05 | DNASE1L3 | 3.21E-03 |
| ENSG00000114861 | 3 | 71003844 | 71633140 | 187 | 41 | 52289 | 4.2126 | 1.26E-05 | FOXP1 | 1.77E-03 |
| ENSG00000031081 | 3 | 119013220 | 119139561 | 168 | 28 | 52289 | 3.6136 | 1.51E-04 | ARHGAP31 | 1.30E-02 |
| ENSG00000180353 | 3 | 121350246 | 121379774 | 58 | 6 | 52289 | 3.5682 | 1.80E-04 | HCLS1 | 1.50E-02 |
| ENSG00000173230 | 3 | 121382046 | 121468602 | 116 | 11 | 52289 | 3.2704 | 5.37E-04 | GOLGB1 | 3.20E-02 |
| ENSG00000114013 | 3 | 121774213 | 121839983 | 58 | 9 | 52289 | 3.6291 | 1.42E-04 | CD86 | 1.25E-02 |
| ENSG00000114126 | 3 | 141663277 | 141868386 | 310 | 12 | 52289 | 3.296 | 4.90E-04 | TFDP2 | 2.99E-02 |
| ENSG00000250588 | 3 | 158680024 | 159615155 | 555 | 36 | 52289 | 5.0538 | 2.17E-07 | IQCJ-SCHIP1 | 5.97E-05 |
| ENSG00000151967 | 3 | 159557650 | 159615149 | 9 | 2 | 52289 | 7.0516 | 8.84E-13 | SCHIP1 | 1.37E-09 |
| ENSG00000168811 | 3 | 159706537 | 159713806 | 5 | 1 | 52289 | 5.4901 | 2.01E-08 | IL12A | 7.75E-06 |
| ENSG00000248710 | 3 | 159945241 | 160167617 | 69 | 9 | 52289 | 4.5094 | 3.25E-06 | RP11-432B6.3 | 5.58E-04 |
| ENSG00000068885 | 3 | 159974774 | 160117668 | 44 | 7 | 52289 | 4.5267 | 3.00E-06 | IFT80 | 5.36E-04 |
| ENSG00000113810 | 3 | 160117062 | 160152750 | 17 | 4 | 52289 | 3.4771 | 2.53E-04 | SMC4 | 1.93E-02 |
| ENSG00000213186 | 3 | 160150233 | 160203561 | 36 | 5 | 52289 | 3.5181 | 2.17E-04 | TRIM59 | 1.72E-02 |
| ENSG00000186432 | 3 | 160212783 | 160283376 | 43 | 5 | 52289 | 3.4256 | 3.07E-04 | KPNA4 | 2.12E-02 |
| ENSG00000179674 | 3 | 160394948 | 160396233 | 1 | 1 | 52289 | 6.6199 | 1.80E-11 | ARL14 | 2.13E-08 |
| ENSG00000163590 | 3 | 160473390 | 160796695 | 523 | 19 | 52289 | 3.2601 | 5.57E-04 | PPM1L | 3.27E-02 |
| ENSG00000180611 | 3 | 192514604 | 192635950 | 56 | 3 | 52289 | 3.4163 | 3.17E-04 | MB21D2 | 2.17E-02 |
| ENSG00000185619 | 4 | 699537 | 764428 | 103 | 13 | 52289 | 3.9659 | 3.66E-05 | PCGF3 | 4.41E-03 |
| ENSG00000178950 | 4 | 843064 | 926161 | 214 | 23 | 52289 | 3.3891 | 3.51E-04 | GAK | 2.32E-02 |
| ENSG00000127419 | 4 | 926175 | 952444 | 30 | 7 | 52289 | 4.9113 | 4.52E-07 | TMEM175 | 1.13E-04 |
| ENSG00000125386 | 4 | 2626988 | 2734292 | 151 | 13 | 52289 | 3.2098 | 6.64E-04 | FAM193A | 3.72E-02 |
| ENSG00000074966 | 4 | 48068410 | 48136273 | 88 | 11 | 52289 | 3.7316 | 9.51E-05 | TXK | 9.42E-03 |
| ENSG00000135605 | 4 | 48137800 | 48271881 | 324 | 22 | 52289 | 3.3963 | 3.42E-04 | TEC | 2.29E-02 |
| ENSG00000070190 | 4 | 100737990 | 100791311 | 50 | 7 | 52289 | 3.3158 | 4.57E-04 | DAPP1 | 2.83E-02 |
| ENSG00000109270 | 4 | 100799493 | 100815647 | 5 | 2 | 52289 | 4.2318 | 1.16E-05 | LAMTOR3 | 1.67E-03 |
| ENSG00000109320 | 4 | 103422486 | 103538459 | 19 | 5 | 52289 | 6.8211 | 4.52E-12 | NFKB1 | 5.91E-09 |
| ENSG00000109323 | 4 | 103552660 | 103682151 | 22 | 5 | 52289 | 7.2788 | 1.68E-13 | MANBA | 3.25E-10 |
| ENSG00000138778 | 4 | 104026963 | 104119566 | 90 | 5 | 52289 | 3.468 | 2.62E-04 | CENPE | 1.95E-02 |
| ENSG00000213949 | 5 | 52083730 | 52255040 | 180 | 30 | 52289 | 3.7354 | 9.37E-05 | ITGA1 | 9.34E-03 |
| ENSG00000164307 | 5 | 96096521 | 96143803 | 193 | 15 | 52289 | 4.8906 | 5.03E-07 | ERAP1 | 1.21E-04 |
| ENSG00000247121 | 5 | 96149731 | 96271513 | 306 | 20 | 52289 | 4.1979 | 1.35E-05 | CTD-2260A17.2 | 1.86E-03 |
| ENSG00000164308 | 5 | 96211643 | 96255420 | 64 | 7 | 52289 | 3.9576 | 3.78E-05 | ERAP2 | 4.53E-03 |
| ENSG00000113441 | 5 | 96271098 | 96373219 | 129 | 6 | 52289 | 3.3073 | 4.71E-04 | LNPEP | 2.91E-02 |
| ENSG00000113532 | 5 | 100142639 | 100238970 | 149 | 8 | 52289 | 4.7409 | 1.06E-06 | ST8SIA4 | 2.24E-04 |
| ENSG00000172869 | 5 | 118373467 | 118584833 | 99 | 13 | 52289 | 3.107 | 9.45E-04 | DMXL1 | 4.93E-02 |
| ENSG00000145901 | 5 | 150409506 | 150473138 | 117 | 18 | 52289 | 5.4166 | 3.04E-08 | TNIP1 | 1.12E-05 |
| ENSG00000145860 | 5 | 158584417 | 158637061 | 48 | 9 | 52289 | 3.2861 | 5.08E-04 | RNF145 | 3.07E-02 |
| ENSG00000164332 | 5 | 158690089 | 158713044 | 14 | 3 | 52289 | 5.9137 | 1.67E-09 | UBLCP1 | 8.93E-07 |
| ENSG00000113302 | 5 | 158741791 | 158757895 | 5 | 1 | 52289 | 3.5229 | 2.13E-04 | IL12B | 1.71E-02 |
| ENSG00000169223 | 5 | 176758563 | 176778853 | 10 | 1 | 52289 | 3.5426 | 1.98E-04 | LMAN2 | 1.63E-02 |
| ENSG00000169220 | 5 | 176784838 | 176799602 | 3 | 1 | 52289 | 3.9155 | 4.51E-05 | RGS14 | 5.32E-03 |
| ENSG00000010810 | 6 | 111981535 | 112194655 | 438 | 26 | 52289 | 3.5283 | 2.09E-04 | FYN | 1.68E-02 |
| ENSG00000118503 | 6 | 138188351 | 138204449 | 22 | 3 | 52289 | 4.5054 | 3.31E-06 | TNFAIP3 | 5.62E-04 |
| ENSG00000105855 | 7 | 20370325 | 20455377 | 150 | 16 | 52289 | 5.1285 | 1.46E-07 | ITGB8 | 4.18E-05 |
| ENSG00000155849 | 7 | 36893961 | 37488852 | 777 | 67 | 52289 | 6.1648 | 3.53E-10 | ELMO1 | 3.03E-07 |
| ENSG00000128595 | 7 | 128379346 | 128411861 | 28 | 1 | 52289 | 4.525 | 3.02E-06 | CALU | 5.36E-04 |
| ENSG00000128617 | 7 | 128412545 | 128415844 | 3 | 1 | 52289 | 3.6514 | 1.30E-04 | OPN1SW | 1.19E-02 |
| ENSG00000128604 | 7 | 128577666 | 128590089 | 7 | 2 | 52289 | 6.1094 | 5.00E-10 | IRF5 | 3.28E-07 |
| ENSG00000064419 | 7 | 128594948 | 128695198 | 81 | 8 | 52289 | 6.1094 | 5.00E-10 | TNPO3 | 3.28E-07 |
| ENSG00000146858 | 7 | 138710452 | 138720775 | 15 | 1 | 52289 | 4.1597 | 1.59E-05 | ZC3HAV1L | 2.14E-03 |
| ENSG00000253426 | 8 | 9009252 | 9025646 | 1 | 1 | 52289 | 3.4617 | 2.68E-04 | RP11-10A14.4 | 1.97E-02 |
| ENSG00000147471 | 8 | 37620111 | 37637283 | 1 | 1 | 52289 | 3.1607 | 7.87E-04 | PROSC | 4.26E-02 |
| ENSG00000104365 | 8 | 42128820 | 42189973 | 78 | 7 | 52289 | 3.4364 | 2.95E-04 | IKBKB | 2.06E-02 |
| ENSG00000104388 | 8 | 61429416 | 61536186 | 128 | 6 | 52289 | 4.9831 | 3.13E-07 | RAB2A | 8.48E-05 |
| ENSG00000104427 | 8 | 79578282 | 79632000 | 89 | 7 | 52289 | 3.6984 | 1.08E-04 | ZC2HC1A | 1.03E-02 |
| ENSG00000104432 | 8 | 79587978 | 79717758 | 196 | 14 | 52289 | 3.4777 | 2.53E-04 | IL7 | 1.93E-02 |
| ENSG00000137077 | 9 | 34709002 | 34710121 | 1 | 1 | 52289 | 3.8158 | 6.79E-05 | CCL21 | 7.38E-03 |
| ENSG00000197816 | 9 | 100000765 | 100140806 | 135 | 6 | 52289 | 3.7892 | 7.56E-05 | CCDC180 | 8.05E-03 |
| ENSG00000136938 | 9 | 100745643 | 100778225 | 8 | 3 | 52289 | 5.6033 | 1.05E-08 | ANP32B | 4.52E-06 |
| ENSG00000095380 | 9 | 100819021 | 100845357 | 12 | 2 | 52289 | 5.9406 | 1.42E-09 | NANS | 8.12E-07 |
| ENSG00000106785 | 9 | 100831557 | 100881494 | 17 | 3 | 52289 | 3.8682 | 5.48E-05 | TRIM14 | 6.27E-03 |
| ENSG00000134460 | 10 | 6052652 | 6104288 | 88 | 19 | 52289 | 3.4945 | 2.37E-04 | IL2RA | 1.83E-02 |
| ENSG00000107968 | 10 | 30722866 | 30750762 | 11 | 2 | 52289 | 3.6803 | 1.16E-04 | MAP3K8 | 1.09E-02 |
| ENSG00000128815 | 10 | 49892921 | 50191001 | 407 | 38 | 52289 | 3.3734 | 3.71E-04 | WDFY4 | 2.43E-02 |
| ENSG00000166507 | 10 | 75561669 | 75571589 | 5 | 2 | 52289 | 3.219 | 6.43E-04 | NDST2 | 3.65E-02 |
| ENSG00000174915 | 11 | 448268 | 491393 | 63 | 5 | 52289 | 3.7714 | 8.12E-05 | PTDSS2 | 8.47E-03 |
| ENSG00000070047 | 11 | 576486 | 612222 | 50 | 6 | 52289 | 5.365 | 4.05E-08 | PHRF1 | 1.45E-05 |
| ENSG00000177030 | 11 | 644233 | 706715 | 74 | 7 | 52289 | 4.7386 | 1.08E-06 | DEAF1 | 2.24E-04 |
| ENSG00000166788 | 11 | 18091482 | 18127638 | 71 | 5 | 52289 | 3.4206 | 3.12E-04 | SAAL1 | 2.15E-02 |
| ENSG00000149089 | 11 | 34874641 | 34938046 | 273 | 10 | 52289 | 3.6286 | 1.42E-04 | APIP | 1.25E-02 |
| ENSG00000110435 | 11 | 34937376 | 35042138 | 287 | 12 | 52289 | 4.23 | 1.17E-05 | PDHX | 1.67E-03 |
| ENSG00000165912 | 11 | 47199076 | 47207994 | 10 | 2 | 52289 | 3.2442 | 5.89E-04 | PACSIN3 | 3.39E-02 |
| ENSG00000134571 | 11 | 47352957 | 47374253 | 14 | 3 | 52289 | 3.5909 | 1.65E-04 | MYBPC3 | 1.41E-02 |
| ENSG00000219435 | 11 | 64067863 | 64072242 | 5 | 2 | 52289 | 3.6677 | 1.22E-04 | TEX40 | 1.14E-02 |
| ENSG00000110777 | 11 | 111222977 | 111326355 | 77 | 11 | 52289 | 3.5829 | 1.70E-04 | POU2AF1 | 1.44E-02 |
| ENSG00000170145 | 11 | 111473115 | 111601577 | 13 | 2 | 52289 | 3.5606 | 1.85E-04 | SIK2 | 1.54E-02 |
| ENSG00000160654 | 11 | 118215059 | 118225876 | 9 | 2 | 52289 | 3.1895 | 7.13E-04 | CD3G | 3.94E-02 |
| ENSG00000110344 | 11 | 118230300 | 118269926 | 42 | 6 | 52289 | 3.2885 | 5.04E-04 | UBE4A | 3.06E-02 |
| ENSG00000167283 | 11 | 118271869 | 118302211 | 41 | 5 | 52289 | 3.4932 | 2.39E-04 | ATP5L | 1.83E-02 |
| ENSG00000019144 | 11 | 118477155 | 118528741 | 16 | 3 | 52289 | 4.8701 | 5.58E-07 | PHLDB1 | 1.27E-04 |
| ENSG00000110367 | 11 | 118620034 | 118661858 | 2 | 1 | 52289 | 4.9765 | 3.24E-07 | DDX6 | 8.62E-05 |
| ENSG00000160683 | 11 | 118754475 | 118768508 | 2 | 1 | 52289 | 5.6029 | 1.05E-08 | CXCR5 | 4.52E-06 |
| ENSG00000186174 | 11 | 118764584 | 118796317 | 2 | 1 | 52289 | 4.0766 | 2.29E-05 | BCL9L | 2.97E-03 |
| ENSG00000135392 | 12 | 56214744 | 56224608 | 2 | 1 | 52289 | 3.5929 | 1.64E-04 | DNAJC14 | 1.40E-02 |
| ENSG00000257921 | 12 | 58166811 | 58180829 | 2 | 1 | 52289 | 4.0062 | 3.09E-05 | RP11-571M6.15 | 3.78E-03 |
| ENSG00000123297 | 12 | 58176372 | 58201854 | 3 | 1 | 52289 | 3.7642 | 8.35E-05 | TSFM | 8.59E-03 |
| ENSG00000175215 | 12 | 58213710 | 58240522 | 6 | 1 | 52289 | 3.395 | 3.43E-04 | CTDSP2 | 2.29E-02 |
| ENSG00000204842 | 12 | 111890018 | 112037480 | 3 | 1 | 52289 | 6.1094 | 5.00E-10 | ATXN2 | 3.28E-07 |
| ENSG00000111275 | 12 | 112204691 | 112247782 | 5 | 2 | 52289 | 4.0004 | 3.16E-05 | ALDH2 | 3.85E-03 |
| ENSG00000089022 | 12 | 112279782 | 112334343 | 7 | 2 | 52289 | 4.1112 | 1.97E-05 | MAPKAPK5 | 2.60E-03 |
| ENSG00000111300 | 12 | 112464500 | 112546826 | 10 | 3 | 52289 | 3.4983 | 2.34E-04 | NAA25 | 1.82E-02 |
| ENSG00000173064 | 12 | 112597992 | 112819896 | 40 | 5 | 52289 | 3.7613 | 8.45E-05 | HECTD4 | 8.59E-03 |
| ENSG00000157837 | 12 | 121200313 | 121342174 | 177 | 17 | 52289 | 4.4229 | 4.87E-06 | SPPL3 | 7.91E-04 |
| ENSG00000111325 | 12 | 123459127 | 123464590 | 3 | 1 | 52289 | 4.2711 | 9.72E-06 | OGFOD2 | 1.46E-03 |
| ENSG00000182196 | 12 | 123464607 | 123467456 | 1 | 1 | 52289 | 3.6456 | 1.33E-04 | ARL6IP4 | 1.21E-02 |
| ENSG00000090975 | 12 | 123468027 | 123634562 | 84 | 8 | 52289 | 3.7575 | 8.58E-05 | PITPNM2 | 8.66E-03 |
| ENSG00000051825 | 12 | 123636867 | 123728561 | 155 | 7 | 52289 | 3.452 | 2.78E-04 | MPHOSPH9 | 2.02E-02 |
| ENSG00000111364 | 12 | 124086624 | 124105488 | 67 | 7 | 52289 | 3.3971 | 3.41E-04 | DDX55 | 2.29E-02 |
| ENSG00000185344 | 12 | 124196865 | 124246302 | 69 | 5 | 52289 | 3.2385 | 6.01E-04 | ATP6V0A2 | 3.45E-02 |
| ENSG00000100599 | 14 | 92980118 | 93155339 | 59 | 13 | 52289 | 3.2133 | 6.56E-04 | RIN3 | 3.68E-02 |
| ENSG00000185567 | 14 | 105403581 | 105444694 | 43 | 4 | 52289 | 4.0253 | 2.85E-05 | AHNAK2 | 3.54E-03 |
| ENSG00000140104 | 14 | 105452112 | 105476819 | 51 | 5 | 52289 | 3.1422 | 8.38E-04 | C14orf79 | 4.48E-02 |
| ENSG00000170779 | 14 | 105475910 | 105487485 | 55 | 4 | 52289 | 3.266 | 5.45E-04 | CDCA4 | 3.23E-02 |
| ENSG00000172575 | 15 | 38780304 | 38857776 | 19 | 4 | 52289 | 3.1212 | 9.01E-04 | RASGRP1 | 4.74E-02 |
| ENSG00000103811 | 15 | 79213400 | 79241916 | 18 | 2 | 52289 | 3.6362 | 1.38E-04 | CTSH | 1.24E-02 |
| ENSG00000038532 | 16 | 11038345 | 11276046 | 61 | 13 | 52289 | 4.3299 | 7.46E-06 | CLEC16A | 1.13E-03 |
| ENSG00000175643 | 16 | 11343476 | 11445619 | 108 | 11 | 52289 | 6.5342 | 3.20E-11 | RMI2 | 3.53E-08 |
| ENSG00000149930 | 16 | 29984962 | 30003582 | 11 | 2 | 52289 | 3.372 | 3.73E-04 | TAOK2 | 2.43E-02 |
| ENSG00000149927 | 16 | 30016830 | 30034591 | 9 | 1 | 52289 | 3.7834 | 7.73E-05 | DOC2A | 8.18E-03 |
| ENSG00000149926 | 16 | 30035748 | 30064299 | 2 | 1 | 52289 | 3.6285 | 1.43E-04 | FAM57B | 1.25E-02 |
| ENSG00000169221 | 16 | 30368423 | 30381585 | 1 | 1 | 52289 | 3.1468 | 8.25E-04 | TBC1D10B | 4.44E-02 |
| ENSG00000180035 | 16 | 30389427 | 30411429 | 13 | 1 | 52289 | 3.3343 | 4.28E-04 | ZNF48 | 2.71E-02 |
| ENSG00000180096 | 16 | 30389454 | 30407312 | 11 | 1 | 52289 | 3.303 | 4.78E-04 | 1-Sep | 2.94E-02 |
| ENSG00000270466 | 16 | 30389755 | 30393863 | 4 | 1 | 52289 | 3.3215 | 4.48E-04 | 1-Sep | 2.81E-02 |
| ENSG00000169896 | 16 | 31271311 | 31344213 | 72 | 6 | 52289 | 3.5817 | 1.71E-04 | ITGAM | 1.44E-02 |
| ENSG00000124074 | 16 | 67696848 | 67701168 | 1 | 1 | 52289 | 4.4188 | 4.96E-06 | ENKD1 | 7.98E-04 |
| ENSG00000141098 | 16 | 67708434 | 67753324 | 9 | 1 | 52289 | 4.883 | 5.23E-07 | GFOD2 | 1.22E-04 |
| ENSG00000141084 | 16 | 67757005 | 67840555 | 28 | 3 | 52289 | 4.0285 | 2.81E-05 | RANBP10 | 3.52E-03 |
| ENSG00000102904 | 16 | 67840668 | 67866051 | 8 | 2 | 52289 | 3.5408 | 1.99E-04 | TSNAXIP1 | 1.63E-02 |
| ENSG00000141096 | 16 | 68009566 | 68014732 | 1 | 1 | 52289 | 4.798 | 8.01E-07 | DPEP3 | 1.72E-04 |
| ENSG00000167261 | 16 | 68021297 | 68034489 | 3 | 1 | 52289 | 6.1094 | 5.00E-10 | DPEP2 | 3.28E-07 |
| ENSG00000167264 | 16 | 68021649 | 68113223 | 16 | 2 | 52289 | 5.6953 | 6.16E-09 | DUS2 | 2.88E-06 |
| ENSG00000129910 | 16 | 89238175 | 89261900 | 24 | 1 | 52289 | 3.4737 | 2.57E-04 | CDH15 | 1.94E-02 |
| ENSG00000259803 | 16 | 89262406 | 89268072 | 1 | 1 | 52289 | 3.3891 | 3.51E-04 | SLC22A31 | 2.32E-02 |
| ENSG00000179593 | 17 | 7942335 | 7952452 | 14 | 3 | 52289 | 3.335 | 4.26E-04 | ALOX15B | 2.71E-02 |
| ENSG00000108278 | 17 | 34842473 | 34855154 | 5 | 2 | 52289 | 3.2549 | 5.67E-04 | ZNHIT3 | 3.29E-02 |
| ENSG00000108306 | 17 | 37415384 | 37558776 | 66 | 5 | 52289 | 3.6826 | 1.15E-04 | FBXL20 | 1.09E-02 |
| ENSG00000125686 | 17 | 37560538 | 37607539 | 36 | 3 | 52289 | 6.0625 | 6.70E-10 | MED1 | 4.14E-07 |
| ENSG00000131748 | 17 | 37793318 | 37819737 | 24 | 4 | 52289 | 5.1757 | 1.14E-07 | STARD3 | 3.65E-05 |
| ENSG00000173991 | 17 | 37820440 | 37822808 | 1 | 1 | 52289 | 7.713 | 6.15E-15 | TCAP | 2.14E-11 |
| ENSG00000141744 | 17 | 37824234 | 37826728 | 1 | 1 | 52289 | 7.4775 | 3.79E-14 | PNMT | 9.75E-11 |
| ENSG00000161395 | 17 | 37827375 | 37853050 | 28 | 2 | 52289 | 7.8622 | 1.89E-15 | PGAP3 | 9.72E-12 |
| ENSG00000141736 | 17 | 37844167 | 37886679 | 15 | 2 | 52289 | 5.7319 | 4.97E-09 | ERBB2 | 2.40E-06 |
| ENSG00000161405 | 17 | 37921198 | 38020441 | 166 | 1 | 52289 | 3.7438 | 9.06E-05 | IKZF3 | 9.09E-03 |
| ENSG00000204913 | 17 | 38097727 | 38101000 | 1 | 1 | 52289 | 7.3368 | 1.09E-13 | LRRC3C | 2.41E-10 |
| ENSG00000108344 | 17 | 38137050 | 38154213 | 23 | 4 | 52289 | 4.1801 | 1.46E-05 | PSMD3 | 1.97E-03 |
| ENSG00000008838 | 17 | 38175350 | 38217468 | 17 | 4 | 52289 | 4.0392 | 2.68E-05 | MED24 | 3.39E-03 |
| ENSG00000159314 | 17 | 43471275 | 43511787 | 17 | 3 | 52289 | 4.661 | 1.57E-06 | ARHGAP27 | 3.08E-04 |
| ENSG00000225190 | 17 | 43513266 | 43568115 | 17 | 1 | 52289 | 5.1297 | 1.45E-07 | PLEKHM1 | 4.18E-05 |
| ENSG00000120088 | 17 | 43699267 | 43913194 | 239 | 8 | 52289 | 6.293 | 1.56E-10 | CRHR1 | 1.41E-07 |
| ENSG00000185294 | 17 | 43922256 | 43924438 | 1 | 1 | 52289 | 5.9133 | 1.68E-09 | SPPL2C | 8.93E-07 |
| ENSG00000186868 | 17 | 43971748 | 44105700 | 28 | 3 | 52289 | 6.3615 | 9.99E-11 | MAPT | 9.64E-08 |
| ENSG00000120071 | 17 | 44107282 | 44302733 | 12 | 2 | 52289 | 3.8409 | 6.13E-05 | KANSL1 | 6.81E-03 |
| ENSG00000073969 | 17 | 44668035 | 44834830 | 25 | 3 | 52289 | 3.764 | 8.36E-05 | NSF | 8.59E-03 |
| ENSG00000108379 | 17 | 44839872 | 44910520 | 16 | 1 | 52289 | 5.1328 | 1.43E-07 | WNT3 | 4.18E-05 |
| ENSG00000141279 | 17 | 45600308 | 45700642 | 35 | 4 | 52289 | 3.6247 | 1.45E-04 | NPEPPS | 1.26E-02 |
| ENSG00000108424 | 17 | 45726842 | 45762871 | 36 | 6 | 52289 | 3.7128 | 1.02E-04 | KPNB1 | 9.88E-03 |
| ENSG00000198933 | 17 | 45771447 | 45789416 | 9 | 1 | 52289 | 3.6443 | 1.34E-04 | TBKBP1 | 1.21E-02 |
| ENSG00000073861 | 17 | 45810610 | 45823485 | 18 | 4 | 52289 | 3.3851 | 3.56E-04 | TBX21 | 2.35E-02 |
| ENSG00000005379 | 17 | 56378592 | 56406152 | 43 | 7 | 52289 | 3.2307 | 6.17E-04 | BZRAP1 | 3.52E-02 |
| ENSG00000108443 | 17 | 57970447 | 58027925 | 11 | 2 | 52289 | 3.4005 | 3.36E-04 | RPS6KB1 | 2.28E-02 |
| ENSG00000267318 | 17 | 58018269 | 58050462 | 1 | 1 | 52289 | 3.5579 | 1.87E-04 | RP11-178C3.1 | 1.54E-02 |
| ENSG00000125447 | 17 | 73232694 | 73258444 | 25 | 2 | 52289 | 3.7225 | 9.86E-05 | GGA3 | 9.64E-03 |
| ENSG00000125457 | 17 | 73262309 | 73267308 | 3 | 1 | 52289 | 3.8701 | 5.44E-05 | MIF4GD | 6.27E-03 |
| ENSG00000125454 | 17 | 73269073 | 73285591 | 3 | 1 | 52289 | 3.7626 | 8.41E-05 | SLC25A19 | 8.59E-03 |
| ENSG00000177885 | 17 | 73314157 | 73401790 | 29 | 3 | 52289 | 4.4234 | 4.86E-06 | GRB2 | 7.91E-04 |
| ENSG00000074695 | 18 | 56995055 | 57027194 | 56 | 4 | 52289 | 3.447 | 2.83E-04 | LMAN1 | 2.04E-02 |
| ENSG00000090339 | 19 | 10381511 | 10397291 | 5 | 2 | 52289 | 3.6268 | 1.43E-04 | ICAM1 | 1.25E-02 |
| ENSG00000076662 | 19 | 10444452 | 10450499 | 19 | 2 | 52289 | 4.7164 | 1.20E-06 | ICAM3 | 2.44E-04 |
| ENSG00000105401 | 19 | 10501810 | 10530797 | 25 | 3 | 52289 | 3.7175 | 1.01E-04 | CDC37 | 9.77E-03 |
| ENSG00000131351 | 19 | 17160539 | 17186435 | 1 | 1 | 52289 | 4.4689 | 3.93E-06 | HAUS8 | 6.60E-04 |
| ENSG00000096996 | 19 | 18169805 | 18209754 | 30 | 3 | 52289 | 6.8188 | 4.59E-12 | IL12RB1 | 5.91E-09 |
| ENSG00000099308 | 19 | 18208603 | 18262502 | 61 | 6 | 52289 | 4.703 | 1.28E-06 | MAST3 | 2.57E-04 |
| ENSG00000254858 | 19 | 18303992 | 18307758 | 5 | 2 | 52289 | 4.8086 | 7.60E-07 | MPV17L2 | 1.68E-04 |
| ENSG00000105650 | 19 | 18318771 | 18366229 | 35 | 5 | 52289 | 3.8915 | 4.98E-05 | PDE4C | 5.78E-03 |
| ENSG00000130518 | 19 | 18367908 | 18385319 | 21 | 5 | 52289 | 3.7972 | 7.32E-05 | KIAA1683 | 7.85E-03 |
| ENSG00000105186 | 19 | 33087913 | 33167503 | 285 | 13 | 52289 | 3.1363 | 8.56E-04 | ANKRD27 | 4.56E-02 |
| ENSG00000131941 | 19 | 33469499 | 33555794 | 41 | 4 | 52289 | 3.3007 | 4.82E-04 | RHPN2 | 2.96E-02 |
| ENSG00000076650 | 19 | 33571786 | 33621448 | 90 | 5 | 52289 | 3.2851 | 5.10E-04 | GPATCH1 | 3.07E-02 |
| ENSG00000105220 | 19 | 34850385 | 34893061 | 36 | 5 | 52289 | 3.5152 | 2.20E-04 | GPI | 1.73E-02 |
| ENSG00000266953 | 19 | 34887220 | 34900269 | 9 | 2 | 52289 | 3.5036 | 2.29E-04 | RP11-618P17.4 | 1.80E-02 |
| ENSG00000126249 | 19 | 34895289 | 34917073 | 18 | 3 | 52289 | 3.2752 | 5.28E-04 | PDCD2L | 3.16E-02 |
| ENSG00000126261 | 19 | 34919257 | 34960853 | 28 | 3 | 52289 | 3.2589 | 5.59E-04 | UBA2 | 3.27E-02 |
| ENSG00000105205 | 19 | 40221890 | 40228668 | 1 | 1 | 52289 | 3.1877 | 7.17E-04 | CLC | 3.96E-02 |
| ENSG00000105223 | 19 | 40854363 | 40886346 | 6 | 2 | 52289 | 3.3548 | 3.97E-04 | PLD3 | 2.57E-02 |
| ENSG00000105438 | 19 | 48885827 | 48894810 | 3 | 1 | 52289 | 3.2822 | 5.15E-04 | KDELR1 | 3.09E-02 |
| ENSG00000176920 | 19 | 49199228 | 49209207 | 1 | 1 | 52289 | 3.1098 | 9.36E-04 | FUT2 | 4.90E-02 |
| ENSG00000131408 | 19 | 50832910 | 50886239 | 15 | 2 | 52289 | 4.4255 | 4.81E-06 | NR1H2 | 7.91E-04 |
| ENSG00000105383 | 19 | 51728320 | 51747115 | 3 | 2 | 52289 | 3.6389 | 1.37E-04 | CD33 | 1.23E-02 |
| ENSG00000167562 | 19 | 53059075 | 53090427 | 2 | 1 | 52289 | 3.3295 | 4.35E-04 | ZNF701 | 2.74E-02 |
| ENSG00000180089 | 19 | 55738007 | 55741647 | 1 | 1 | 52289 | 3.267 | 5.43E-04 | TMEM86B | 3.23E-02 |
| ENSG00000132823 | 20 | 42825136 | 42839431 | 7 | 2 | 52289 | 3.4169 | 3.17E-04 | OSER1 | 2.17E-02 |
| ENSG00000101017 | 20 | 44746911 | 44758502 | 7 | 2 | 52289 | 4.5426 | 2.78E-06 | CD40 | 5.05E-04 |
| ENSG00000159110 | 21 | 34602206 | 34637980 | 7 | 2 | 52289 | 3.8219 | 6.62E-05 | IFNAR2 | 7.25E-03 |
| ENSG00000159128 | 21 | 34775202 | 34851655 | 19 | 2 | 52289 | 3.3168 | 4.55E-04 | IFNGR2 | 2.83E-02 |
| ENSG00000185651 | 22 | 21903736 | 21978323 | 43 | 1 | 52289 | 3.855 | 5.79E-05 | UBE2L3 | 6.52E-03 |
| ENSG00000161179 | 22 | 21982378 | 21984353 | 2 | 1 | 52289 | 3.1785 | 7.40E-04 | YDJC | 4.05E-02 |
| ENSG00000100024 | 22 | 24863206 | 24924358 | 51 | 4 | 52289 | 3.2556 | 5.66E-04 | UPB1 | 3.29E-02 |
| ENSG00000100330 | 22 | 30279144 | 30426855 | 120 | 12 | 52289 | 3.4693 | 2.61E-04 | MTMR3 | 1.95E-02 |
| ENSG00000182541 | 22 | 31608225 | 31676066 | 51 | 9 | 52289 | 3.1628 | 7.81E-04 | LIMK2 | 4.25E-02 |
| ENSG00000100335 | 22 | 39895437 | 39914137 | 5 | 1 | 52289 | 3.4367 | 2.94E-04 | MIEF1 | 2.06E-02 |
| ENSG00000128272 | 22 | 39915700 | 39918691 | 1 | 1 | 52289 | 3.3436 | 4.14E-04 | ATF4 | 2.64E-02 |
| ENSG00000100403 | 22 | 41697526 | 41756151 | 3 | 2 | 52289 | 3.6817 | 1.16E-04 | ZC3H7B | 1.09E-02 |
| ENSG00000167074 | 22 | 41763337 | 41795330 | 6 | 2 | 52289 | 4.0541 | 2.52E-05 | TEF | 3.21E-03 |
| ENSG00000183864 | 22 | 41829496 | 41843027 | 2 | 1 | 52289 | 3.154 | 8.05E-04 | TOB2 | 4.35E-02 |
| ENSG00000025770 | 22 | 50946645 | 50961901 | 3 | 1 | 52289 | 3.2587 | 5.60E-04 | NCAPH2 | 3.27E-02 |
| ENSG00000130489 | 22 | 50961997 | 50964868 | 1 | 1 | 52289 | 3.5349 | 2.04E-04 | SCO2 | 1.65E-02 |

MAGMA: “Multi-marker Analysis of GenoMic Annotation”. NSNPS: number of SNPs. NPARAM: number of relevant parameters used in the model. FDR: false discovery rate.

**Supplemental Table 10 Genes prioritized by DEPICT**

| Locus | N of genes | Chromosome and position | GWAS P value | Ensembl gene ID | Gene symbol | Nominal P value | FDR Q value |
| --- | --- | --- | --- | --- | --- | --- | --- |
| rs7528684 | 5 | chr1:157543539-157868046 | 3.41E-09 | ENSG00000163534 | FCRL1 | 2.71E-16 | <=0.01 |
| rs230534 | 1 | chr4:103422486-103538459 | 1.03E-21 | ENSG00000109320 | NFKB1 | 2.25E-11 | <=0.01 |
| rs7528684 | 5 | chr1:157543539-157868046 | 3.41E-09 | ENSG00000132704 | FCRL2 | 2.57E-11 | <=0.01 |
| rs1422673 | 1 | chr5:150409506-150473138 | 2.87E-09 | ENSG00000145901 | TNIP1 | 3.17E-11 | <=0.01 |
| rs2305743 | 1 | chr19:18170371-18197697 | 1.95E-14 | ENSG00000096996 | IL12RB1 | 3.86E-11 | <=0.01 |
| rs56410675;rs8071789 | 3 | chr17:37913968-38077313 | 3.19E-33 | ENSG00000161405 | IKZF3 | 4.32E-11 | <=0.01 |
| rs7528684 | 5 | chr1:157543539-157868046 | 3.41E-09 | ENSG00000160856 | FCRL3 | 7.00E-11 | <=0.01 |
| rs11588376 | 2 | chr1:117035645-117113661 | 2.29E-08 | ENSG00000116815 | CD58 | 1.58E-10 | <=0.01 |
| rs117710929;rs145809305;rs70600 | 13 | chr17:43471275-44896082 | 1.56E-11 | ENSG00000159314 | ARHGAP27 | 2.01E-10 | <=0.01 |
| rs9843053 | 7 | chr3:121311966-121741051 | 3.34E-09 | ENSG00000180353 | HCLS1 | 2.20E-10 | <=0.01 |
| rs4451969 | 5 | chr16:11348262-11445619 | 3.51E-17 | ENSG00000185338 | SOCS1 | 2.26E-10 | <=0.01 |
| rs12706861;rs4728142 | 2 | chr7:128577666-128695198 | 2.37E-40 | ENSG00000128604 | IRF5 | 2.54E-10 | <=0.01 |
| rs1874791;rs6676606 | 2 | chr1:67773047-67896098 | 9.59E-30 | ENSG00000081985 | IL12RB2 | 5.18E-10 | <=0.01 |
| rs1001674;rs9834901 | 5 | chr3:119013220-119278449 | 1.42E-25 | ENSG00000121594 | CD80 | 1.25E-09 | <=0.01 |
| rs598711;rs7311681 | 14 | chr12:111843752-112947717 | 9.06E-12 | ENSG00000135148 | TRAFD1 | 1.62E-09 | <=0.01 |
| rs7528684 | 5 | chr1:157543539-157868046 | 3.41E-09 | ENSG00000163518 | FCRL4 | 2.31E-09 | <=0.01 |
| rs734206 | 1 | chr14:92980118-93155339 | 7.77E-10 | ENSG00000100599 | RIN3 | 3.12E-09 | <=0.01 |
| rs11889341;rs16833239 | 1 | chr2:191894302-192016322 | 1.64E-23 | ENSG00000138378 | STAT4 | 7.03E-09 | <=0.01 |
| rs17445836;rs7202472 | 1 | chr16:85932409-85956197 | 1.08E-26 | ENSG00000140968 | IRF8 | 7.46E-09 | <=0.01 |
| rs2243131;rs79153365;rs9880646 | 2 | chr3:159631189-159943086 | 6.81E-13 | ENSG00000168811 | IL12A | 9.30E-09 | <=0.01 |
| rs10187120;rs1263128;rs1558471 | 1 | chr2:191511472-191557492 | 1.57E-21 | ENSG00000138386 | NAB1 | 9.54E-09 | <=0.01 |
| rs56410675;rs8071789 | 3 | chr17:37913968-38077313 | 3.19E-33 | ENSG00000073605 | GSDMB | 1.15E-08 | <=0.01 |
| rs475032 | 8 | chr11:64018995-64139687 | 5.71E-09 | ENSG00000168071 | CCDC88B | 1.19E-08 | <=0.01 |
| rs9533117 | 1 | chr13:43136872-43182149 | 2.88E-08 | ENSG00000120659 | TNFSF11 | 2.94E-08 | <=0.01 |
| rs7943546 | 10 | chr11:537527-640706 | 1.07E-09 | ENSG00000185507 | IRF7 | 3.30E-08 | <=0.01 |
| rs10042630 | 3 | chr5:158584417-158757895 | 4.42E-10 | ENSG00000113302 | IL12B | 3.33E-08 | <=0.01 |
| rs1788098 | 2 | chr18:67068284-67624160 | 1.89E-09 | ENSG00000150637 | CD226 | 3.51E-08 | <=0.01 |
| rs2022449 | 1 | chr1:173152873-173176452 | 1.68E-10 | ENSG00000117586 | TNFSF4 | 3.81E-08 | <=0.01 |
| rs598711;rs7311681 | 14 | chr12:111843752-112947717 | 9.06E-12 | ENSG00000111252 | SH2B3 | 3.99E-08 | <=0.01 |
| rs8057040 | 2 | chr16:11022748-11276046 | 2.52E-13 | ENSG00000038532 | CLEC16A | 4.62E-08 | <=0.01 |
| rs10042630 | 3 | chr5:158584417-158757895 | 4.42E-10 | ENSG00000145860 | RNF145 | 5.80E-08 | <=0.01 |
| rs7204192 | 28 | chr16:67679030-68482591 | 1.58E-11 | ENSG00000159753 | RLTPR | 7.95E-08 | <=0.01 |
| rs27524 | 3 | chr5:95865525-96271513 | 4.89E-10 | ENSG00000164307 | ERAP1 | 7.98E-08 | <=0.01 |
| rs13062928 | 6 | chr3:159974774-160396233 | 1.12E-16 | ENSG00000179674 | ARL14 | 1.12E-07 | <=0.01 |
| rs4810485 | 3 | chr20:44650329-44758502 | 1.44E-08 | ENSG00000101017 | CD40 | 1.37E-07 | <=0.01 |
| rs10083496 | 2 | chr14:105391153-105444694 | 2.56E-08 | ENSG00000166428 | PLD4 | 1.53E-07 | <=0.01 |
| rs148647102 | 2 | chr2:191054461-191236391 | 1.09E-10 | ENSG00000151689 | INPP1 | 1.63E-07 | <=0.01 |
| rs7204192 | 28 | chr16:67679030-68482591 | 1.58E-11 | ENSG00000205220 | PSMB10 | 1.73E-07 | <=0.01 |
| rs551125 | 5 | chr12:121200313-121477045 | 1.65E-09 | ENSG00000135114 | OASL | 2.07E-07 | <=0.01 |
| rs9843053 | 7 | chr3:121311966-121741051 | 3.34E-09 | ENSG00000145103 | ILDR1 | 2.63E-07 | <=0.01 |
| rs7204192 | 28 | chr16:67679030-68482591 | 1.58E-11 | ENSG00000141086 | CTRL | 5.74E-07 | <=0.01 |
| rs11128810 | 1 | chr3:16844159-17132094 | 2.91E-12 | ENSG00000154822 | PLCL2 | 9.94E-07 | <=0.01 |
| rs16879645 | 1 | chr7:36893961-37488852 | 4.46E-14 | ENSG00000155849 | ELMO1 | 9.97E-07 | <=0.01 |
| rs1001674;rs9834901 | 5 | chr3:119013220-119278449 | 1.42E-25 | ENSG00000031081 | ARHGAP31 | 1.04E-06 | <=0.01 |
| rs117710929;rs145809305;rs70600 | 13 | chr17:43471275-44896082 | 1.56E-11 | ENSG00000256762 | STH | 2.10E-06 | <=0.01 |
| rs27524 | 3 | chr5:95865525-96271513 | 4.89E-10 | ENSG00000247121 | - | 2.26E-06 | <=0.01 |
| rs6944997 | 1 | chr7:20370325-20450419 | 5.09E-09 | ENSG00000105855 | ITGB8 | 3.31E-06 | <=0.01 |
| rs75904251 | 1 | chr2:25455845-25565459 | 3.24E-09 | ENSG00000119772 | DNMT3A | 7.18E-06 | <=0.01 |
| rs11588376 | 2 | chr1:117035645-117113661 | 2.29E-08 | ENSG00000224950 | - | 7.66E-06 | <=0.01 |
| rs9843053 | 7 | chr3:121311966-121741051 | 3.34E-09 | ENSG00000145088 | EAF2 | 1.27E-05 | <=0.01 |
| rs3756353 | 1 | chr5:100145357-100238970 | 1.45E-08 | ENSG00000113532 | ST8SIA4 | 1.36E-05 | <=0.01 |
| rs55838263 | 4 | chr1:200860176-201081694 | 2.60E-08 | ENSG00000229191 | - | 1.78E-05 | <=0.01 |
| rs55838263 | 4 | chr1:200860176-201081694 | 2.60E-08 | ENSG00000116852 | KIF21B | 1.83E-05 | <=0.01 |
| rs27524 | 3 | chr5:95865525-96271513 | 4.89E-10 | ENSG00000153113 | CAST | 3.53E-05 | <=0.01 |
| rs475032 | 8 | chr11:64018995-64139687 | 5.71E-09 | ENSG00000173264 | GPR137 | 6.86E-05 | <=0.01 |
| rs7528684 | 5 | chr1:157543539-157868046 | 3.41E-09 | ENSG00000073754 | CD5L | 7.20E-05 | <=0.01 |
| rs7204192 | 28 | chr16:67679030-68482591 | 1.58E-11 | ENSG00000159761 | C16orf86 | 9.65E-05 | <=0.01 |
| rs475032 | 8 | chr11:64018995-64139687 | 5.71E-09 | ENSG00000162302 | RPS6KA4 | 2.08E-04 | <=0.01 |
| rs117710929;rs145809305;rs70600 | 13 | chr17:43471275-44896082 | 1.56E-11 | ENSG00000204650 | - | 4.55E-04 | <=0.01 |
| rs4915526 | 2 | chr1:197170592-197744826 | 2.42E-08 | ENSG00000213047 | DENND1B | 6.60E-04 | <=0.01 |
| rs3755963 | 2 | chr4:843064-952444 | 4.34E-10 | ENSG00000178950 | GAK | 1.10E-03 | <=0.01 |
| rs7204192 | 28 | chr16:67679030-68482591 | 1.58E-11 | ENSG00000072736 | NFATC3 | 1.37E-03 | <=0.01 |
| rs1001674;rs9834901 | 5 | chr3:119013220-119278449 | 1.42E-25 | ENSG00000163389 | POGLUT1 | 2.32E-03 | <0.05 |
| rs4451969 | 5 | chr16:11348262-11445619 | 3.51E-17 | ENSG00000175643 | RMI2 | 7.14E-03 | <0.05 |
| rs7204192 | 28 | chr16:67679030-68482591 | 1.58E-11 | ENSG00000167264 | DUS2L | 7.33E-03 | <0.05 |
| rs7204192 | 28 | chr16:67679030-68482591 | 1.58E-11 | ENSG00000167261 | DPEP2 | 0.01 | <0.05 |
| rs598711;rs7311681 | 14 | chr12:111843752-112947717 | 9.06E-12 | ENSG00000179295 | PTPN11 | 0.01 | <0.05 |
| rs56410675;rs8071789 | 3 | chr17:37913968-38077313 | 3.19E-33 | ENSG00000186075 | ZPBP2 | 0.01 | <0.05 |

DIPECT: “Data-driven Expression Prioritized Integration for Complex Traits”. GWAS: genome-wide association study. FDR: false discovery rate.

**Supplemental Table 11** Predicted regulatory element-target gene pairs by ENCODE-rE2G intersecting with fine-mapped SNPs (PIP > 10%) in novel loci

| Locus | SNP | SNP CHR | SNP BP | PIP | Region Start | Region End | Class | Target Gene | Target Gene Ensembl ID | Cell Type |
| --- | --- | --- | --- | --- | --- | --- | --- | --- | --- | --- |
| SPPL3 | rs551125 | 12 | 121203427 | 0.29 | 121203355 | 121204489 | genic | ACADS | ENSG00000122971 | OCI-LY7 |
|  |  |  |  |  | 121203100 | 121204671 | genic | ACADS | ENSG00000122971 | mucosa_of_descending_colon |
| AHNAK2 | rs10149193 | 14 | 105403474 | 0.27 | 105400953 | 105403825 | genic | AHNAK2 | ENSG00000185567 | GM20000 |
|  |  |  |  |  | 105400953 | 105403825 | genic | PLD4 | ENSG00000166428 | GM20000 |
|  |  |  |  |  | 105403136 | 105403636 | genic | PLD4 | ENSG00000166428 | naive |
|  |  |  |  |  | 105400512 | 105404538 | genic | AHNAK2 | ENSG00000185567 | Panc1 |
|  |  |  |  |  | 105400512 | 105404538 | genic | C14orf79 | ENSG00000140104 | Panc1 |
|  |  |  |  |  | 105400512 | 105404538 | genic | ZBTB42 | ENSG00000179627 | Panc1 |
|  |  |  |  |  | 105401643 | 105404522 | genic | C14orf79 | ENSG00000140104 | Karpas-422 |
|  |  |  |  |  | 105401643 | 105404522 | genic | CEP170B | ENSG00000099814 | Karpas-422 |
|  |  |  |  |  | 105401643 | 105404522 | genic | PLD4 | ENSG00000166428 | Karpas-422 |
|  |  |  |  |  | 105403055 | 105403555 | intergenic | PLD4 | ENSG00000166428 | CMK |
|  | rs28454709 | 14 | 105405942 | 0.32 | 105405918 | 105406418 | genic | PLD4 | ENSG00000166428 | coronary_artery |
| CCDC113/CSNK2A2 | rs2731783 | 16 | 58253460 | 0.19 | 58253272 | 58253772 | intergenic | CCDC113 | ENSG00000103021 | foreskin_melanocyte |
|  |  |  |  |  | 58253290 | 58253790 | intergenic | CCDC113 | ENSG00000103021 | placenta |
|  |  |  |  |  | 58252439 | 58253769 | intergenic | CCDC113 | ENSG00000103021 | trophoblast_cell |
|  |  |  |  |  | 58252439 | 58253769 | intergenic | CSNK2A2 | ENSG00000070770 | trophoblast_cell |
| CD40 | rs1569723 | 20 | 44742064 | 0.10 | 44741460 | 44742905 | intergenic | CD40 | ENSG00000101017 | RWPE2 |
|  |  |  |  |  | 44741561 | 44742192 | intergenic | CD40 | ENSG00000101017 | breast_epithelium |
|  | rs4810485 | 20 | 44747947 | 0.44 | 44746042 | 44748355 | promoter | CD40 | ENSG00000101017 | upper_lobe_of_left_lung |
|  |  |  |  |  | 44746458 | 44748615 | promoter | CD40 | ENSG00000101017 | memory |
|  |  |  |  |  | 44746264 | 44748354 | promoter | CD40 | ENSG00000101017 | common_myeloid_progenitor,_CD34-positive |
|  |  |  |  |  | 44746313 | 44748302 | promoter | CD40 | ENSG00000101017 | ACHN |
|  |  |  |  |  | 44747874 | 44748374 | genic | CD40 | ENSG00000101017 | T-helper_9_cell |
|  |  |  |  |  | 44746624 | 44748004 | promoter | CD40 | ENSG00000101017 | GM10266 |
|  |  |  |  |  | 44746315 | 44748590 | promoter | CD40 | ENSG00000101017 | GM12878 |
|  |  |  |  |  | 44746315 | 44748590 | promoter | MMP9 | ENSG00000100985 | GM12878 |
|  |  |  |  |  | 44746315 | 44748590 | promoter | NCOA5 | ENSG00000124160 | GM12878 |
|  |  |  |  |  | 44746315 | 44748590 | promoter | WFDC10A | ENSG00000180305 | GM12878 |
|  |  |  |  |  | 44747670 | 44748603 | genic | CD40 | ENSG00000101017 | GM19240 |
|  |  |  |  |  | 44746506 | 44748245 | promoter | CD40 | ENSG00000101017 | spleen |
|  |  |  |  |  | 44746256 | 44748139 | promoter | CD40 | ENSG00000101017 | CD4-positive,_alpha-beta_T_cell |
|  |  |  |  |  | 44746398 | 44748033 | promoter | CD40 | ENSG00000101017 | GM20000 |
|  |  |  |  |  | 44746154 | 44748289 | promoter | CD40 | ENSG00000101017 | GM12878 |
|  |  |  |  |  | 44746154 | 44748289 | promoter | NCOA5 | ENSG00000124160 | GM12878 |
|  |  |  |  |  | 44746642 | 44748346 | promoter | CD40 | ENSG00000101017 | RCC_7860 |
|  |  |  |  |  | 44746433 | 44748588 | promoter | CD40 | ENSG00000101017 | OCI-LY7 |
|  |  |  |  |  | 44747670 | 44748170 | genic | CD40 | ENSG00000101017 | liver |
|  |  |  |  |  | 44747549 | 44748138 | genic | CD40 | ENSG00000101017 | GM12891 |
|  |  |  |  |  | 44746191 | 44748193 | promoter | CD40 | ENSG00000101017 | HK-2 |
|  |  |  |  |  | 44747648 | 44748148 | genic | CD40 | ENSG00000101017 | activated_regulatory_T_cell |
|  |  |  |  |  | 44747524 | 44748024 | genic | CD40 | ENSG00000101017 | naive |
|  |  |  |  |  | 44747524 | 44748024 | genic | CDH22 | ENSG00000149654 | naive |
|  |  |  |  |  | 44747524 | 44748024 | genic | MMP9 | ENSG00000100985 | naive |
|  |  |  |  |  | 44747524 | 44748024 | genic | NCOA5 | ENSG00000124160 | naive |
|  |  |  |  |  | 44747524 | 44748024 | genic | SLC12A5 | ENSG00000124140 | naive |
|  |  |  |  |  | 44746100 | 44748027 | promoter | CD40 | ENSG00000101017 | B_cell |
|  |  |  |  |  | 44746077 | 44748497 | promoter | CD40 | ENSG00000101017 | GM12865 |
|  |  |  |  |  | 44746286 | 44748062 | promoter | CD40 | ENSG00000101017 | KBM-7 |
|  |  |  |  |  | 44746292 | 44748141 | promoter | CD40 | ENSG00000101017 | GM12892 |
|  |  |  |  |  | 44746311 | 44748035 | promoter | CD40 | ENSG00000101017 | midbrain |
|  |  |  |  |  | 44747794 | 44748294 | genic | CD40 | ENSG00000101017 | stimulated_activated_CD8-positive,_alpha-beta_memory_T_cell |
|  |  |  |  |  | 44746377 | 44747963 | promoter | CD40 | ENSG00000101017 | RKO |
|  |  |  |  |  | 44747538 | 44748038 | genic | CD40 | ENSG00000101017 | thymus |
|  |  |  |  |  | 44746642 | 44748758 | promoter | CD40 | ENSG00000101017 | NAMALWA |
|  |  |  |  |  | 44746020 | 44748150 | promoter | CD40 | ENSG00000101017 | suprapubic_skin |
|  |  |  |  |  | 44746265 | 44748044 | promoter | CD40 | ENSG00000101017 | GM12864 |
|  |  |  |  |  | 44746326 | 44748157 | promoter | CD40 | ENSG00000101017 | kidney_tubule_cell |
|  |  |  |  |  | 44747834 | 44748334 | genic | CD40 | ENSG00000101017 | CD8-positive,_alpha-beta_memory_T_cell |
|  |  |  |  |  | 44746029 | 44748346 | promoter | CD40 | ENSG00000101017 | lower_lobe_of_left_lung |
|  |  |  |  |  | 44747715 | 44748215 | genic | CD40 | ENSG00000101017 | tibial |
|  |  |  |  |  | 44747761 | 44748353 | genic | CD40 | ENSG00000101017 | RWPE2 |
|  |  |  |  |  | 44747797 | 44748297 | genic | CD40 | ENSG00000101017 | small_intestine |
|  |  |  |  |  | 44746573 | 44748335 | promoter | CD40 | ENSG00000101017 | kidney_glomerular_epithelial_cell |
|  |  |  |  |  | 44746573 | 44748335 | promoter | MMP9 | ENSG00000100985 | kidney_glomerular_epithelial_cell |
|  |  |  |  |  | 44746573 | 44748335 | promoter | NCOA5 | ENSG00000124160 | kidney_glomerular_epithelial_cell |
|  |  |  |  |  | 44747846 | 44748346 | genic | CD40 | ENSG00000101017 | ureter |
|  |  |  |  |  | 44747832 | 44748332 | genic | CD40 | ENSG00000101017 | stimulated_activated_naive_CD4-positive,_alpha-beta_T_cell |
|  |  |  |  |  | 44746421 | 44748040 | promoter | CD40 | ENSG00000101017 | Karpas-422 |
|  |  |  |  |  | 44747860 | 44748360 | genic | CD40 | ENSG00000101017 | central_memory_CD4-positive,_alpha-beta_T_cell |
|  |  |  |  |  | 44747676 | 44748176 | genic | CD40 | ENSG00000101017 | effector_memory_CD8-positive,_alpha-beta_T_cell |
|  |  |  |  |  | 44747815 | 44748315 | genic | CD40 | ENSG00000101017 | DND-41 |
|  |  |  |  |  | 44746588 | 44748308 | promoter | CD40 | ENSG00000101017 | glomerular_visceral_epithelial_cell |
|  |  |  |  |  | 44746588 | 44748308 | promoter | CDH22 | ENSG00000149654 | glomerular_visceral_epithelial_cell |
|  |  |  |  |  | 44746588 | 44748308 | promoter | MMP9 | ENSG00000100985 | glomerular_visceral_epithelial_cell |
|  |  |  |  |  | 44746588 | 44748308 | promoter | NCOA5 | ENSG00000124160 | glomerular_visceral_epithelial_cell |
|  |  |  |  |  | 44746596 | 44748124 | promoter | CD40 | ENSG00000101017 | kidney_epithelial_cell |
|  |  |  |  |  | 44746624 | 44748004 | promoter | CD40 | ENSG00000101017 | germinal |
|  |  |  |  |  | 44747365 | 44748267 | promoter | CD40 | ENSG00000101017 | renal_cortical_epithelial_cell |
|  |  |  |  |  | 44747543 | 44748043 | genic | CD40 | ENSG00000101017 | GM13977 |
|  |  |  |  |  | 44747865 | 44748365 | genic | CD40 | ENSG00000101017 | CD8-positive,_alpha-beta_T_cell |
|  |  |  |  |  | 44746574 | 44748250 | promoter | CD40 | ENSG00000101017 | RCC |
|  |  |  |  |  | 44746574 | 44748250 | promoter | CDH22 | ENSG00000149654 | RCC |
|  |  |  |  |  | 44746574 | 44748250 | promoter | NCOA5 | ENSG00000124160 | RCC |
|  |  |  |  |  | 44747658 | 44748158 | genic | CD40 | ENSG00000101017 | stimulated_activated_effector_memory_CD8-positive,_alpha-beta_T_cell |

CHR: chromosome. BP: base pair.

**Supplemental Figure 1** LocusZoom plots for the none genomic loci that were significant in the cross-phenotype GWAS meta-analysis, without evidence of heterogeneity, and colocalized between SSc and PBC. The fixed-effect model p-values were used for the meta-analysis statistics in the LocusZoom plot.

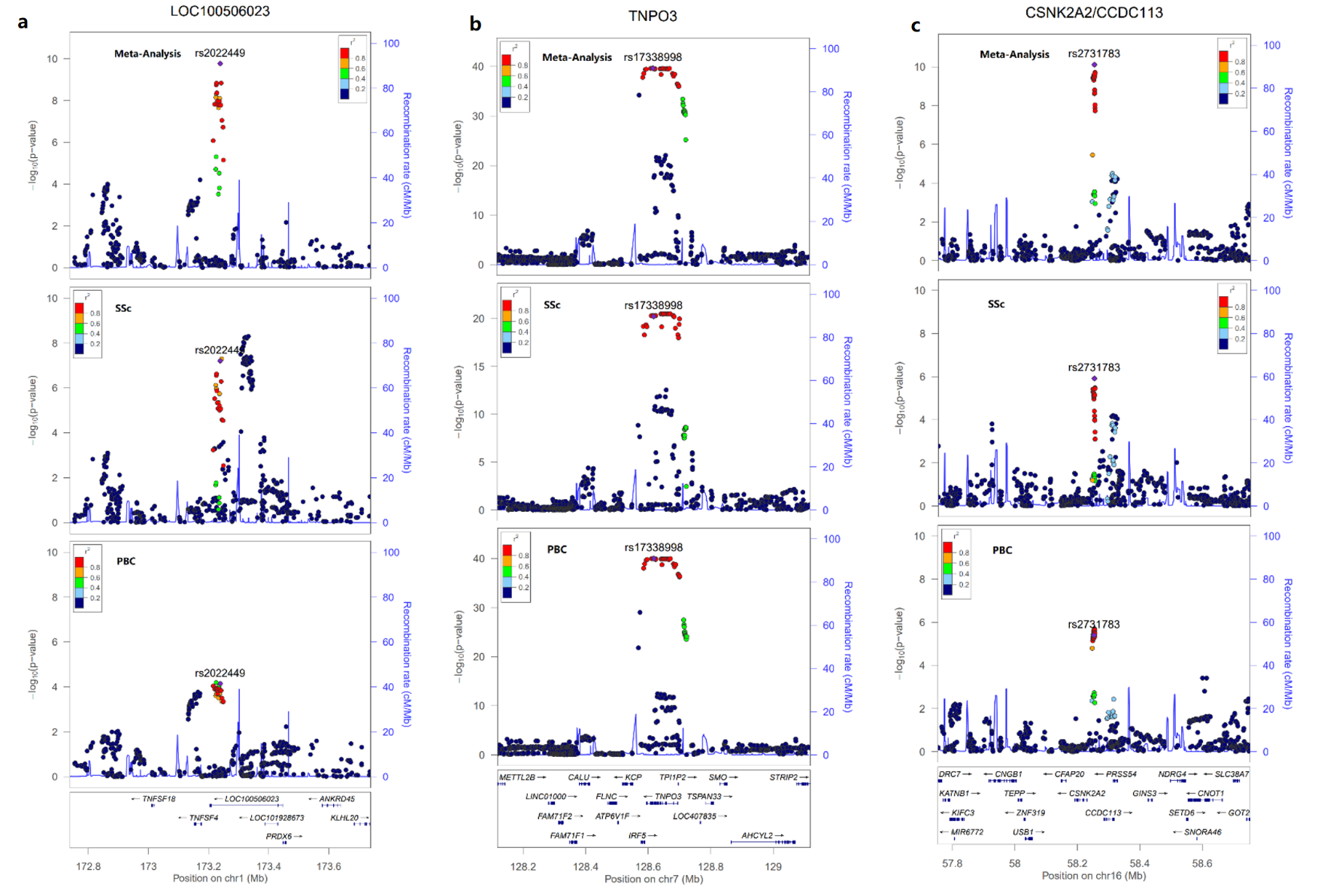

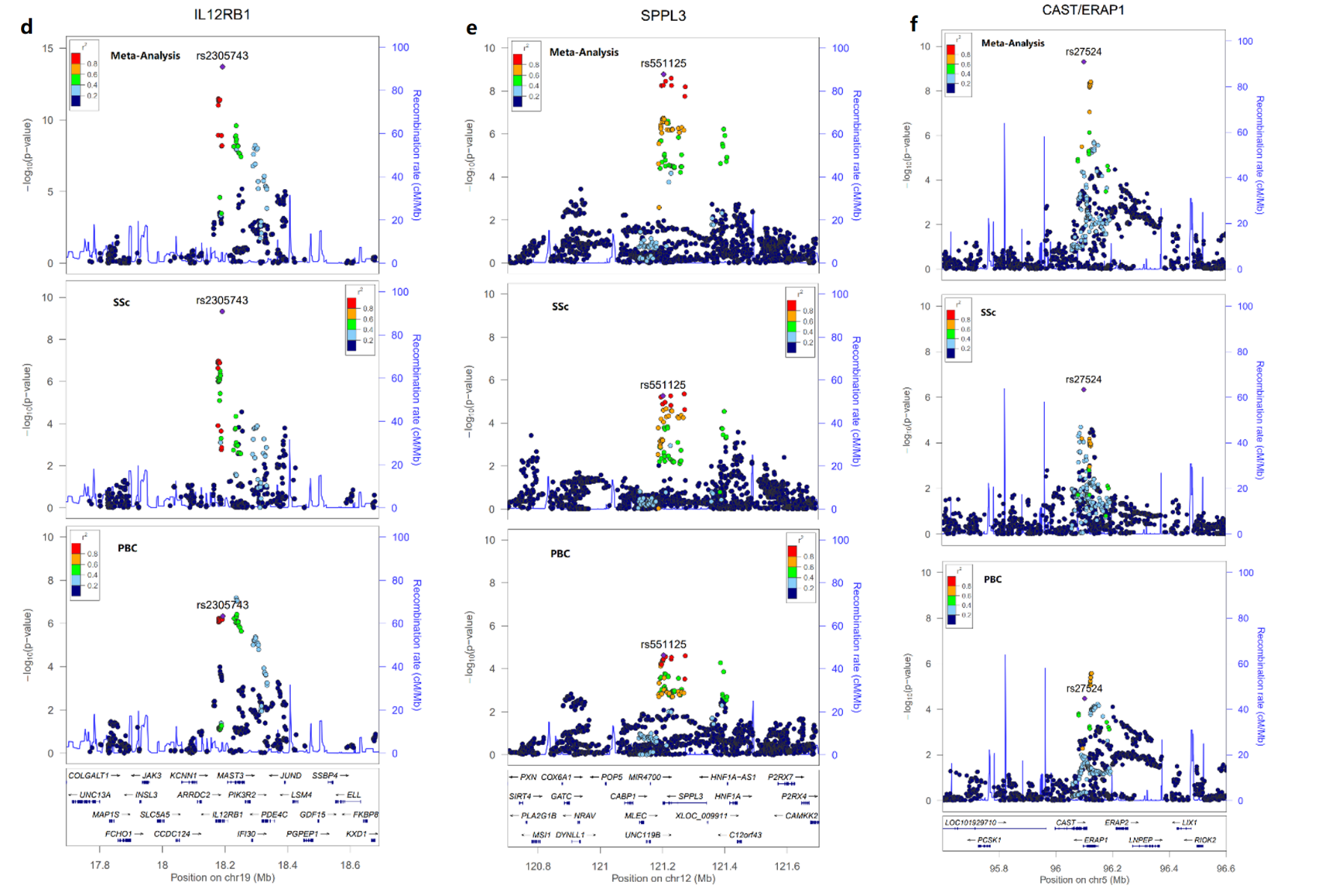

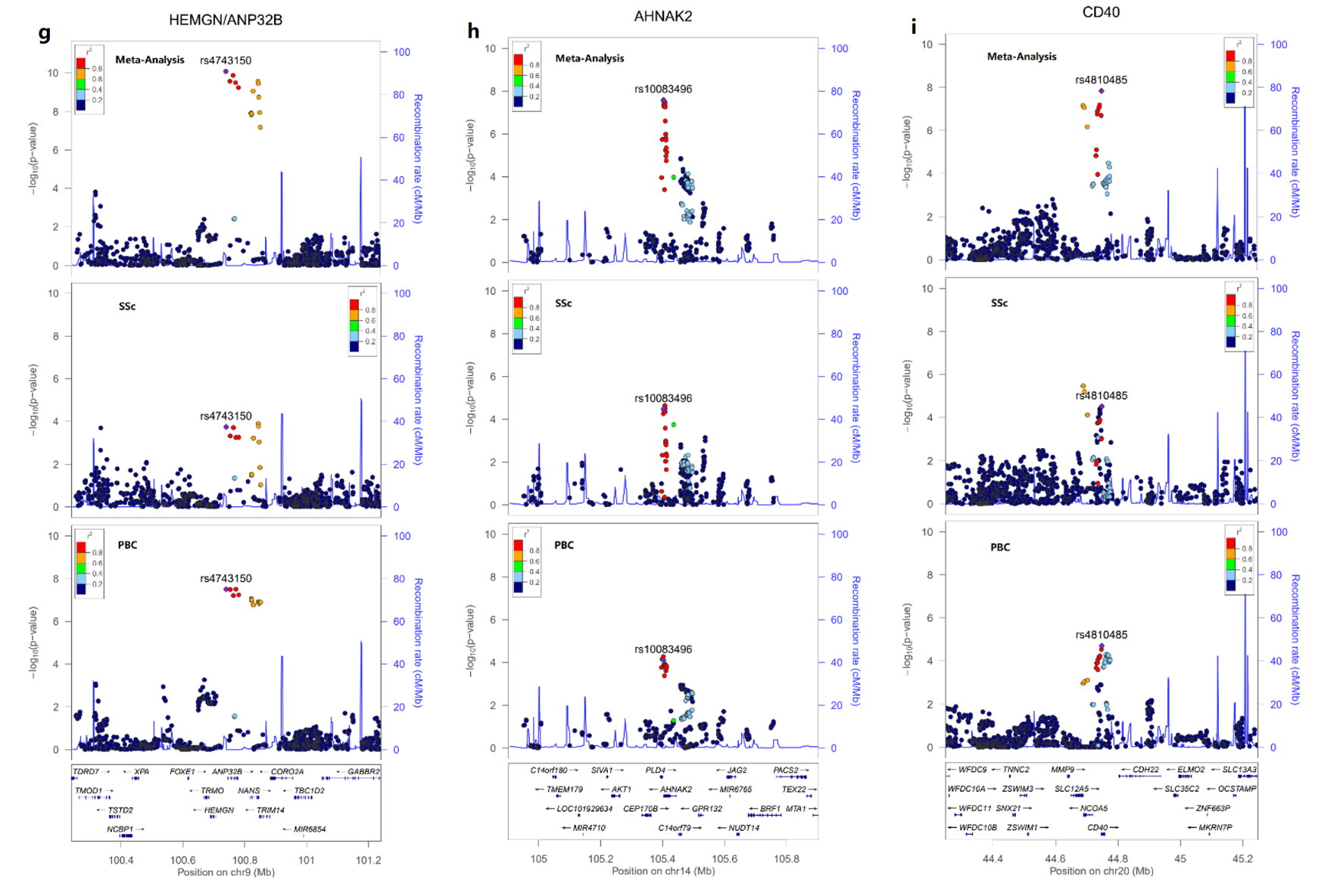

SSc: systemic sclerosis. PBC: primary biliary cholangitis

**Supplemental Figure 2** LocusZoom plots for the four genomic loci that were significant in the cross-phenotype GWAS meta-analysis, with evidence of heterogeneity, and colocalized between SSc and PBC. The PLEIO p-values were used for the meta-analysis statistics in the LocusZoom plot.

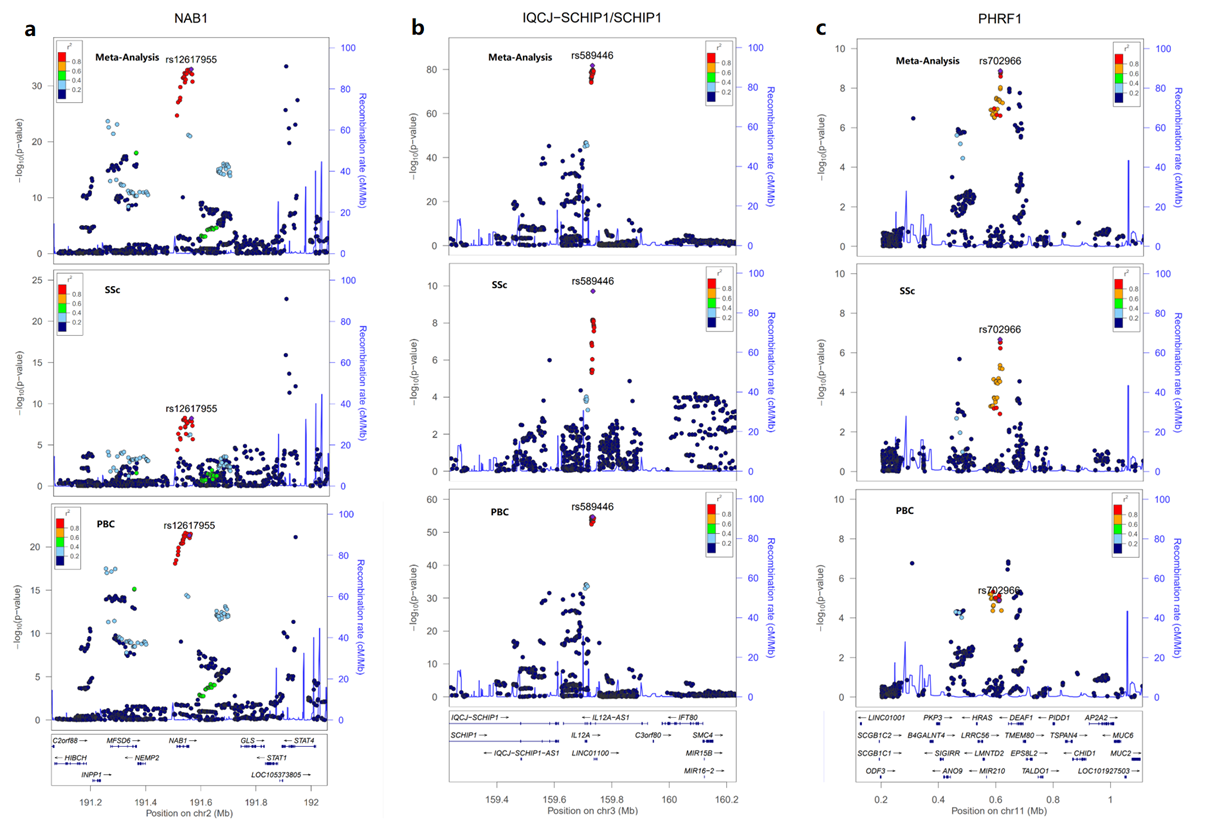

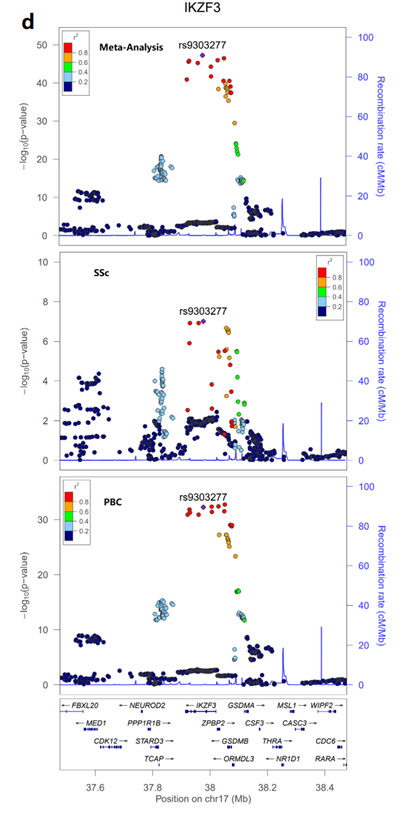

SSc: systemic sclerosis. PBC: primary biliary cholangitis

**Supplemental Figure 3** Meta-PheWAS analyses on the lead SNP of the novel candidate loci using eMERGE, All of Us and UK Biobank.

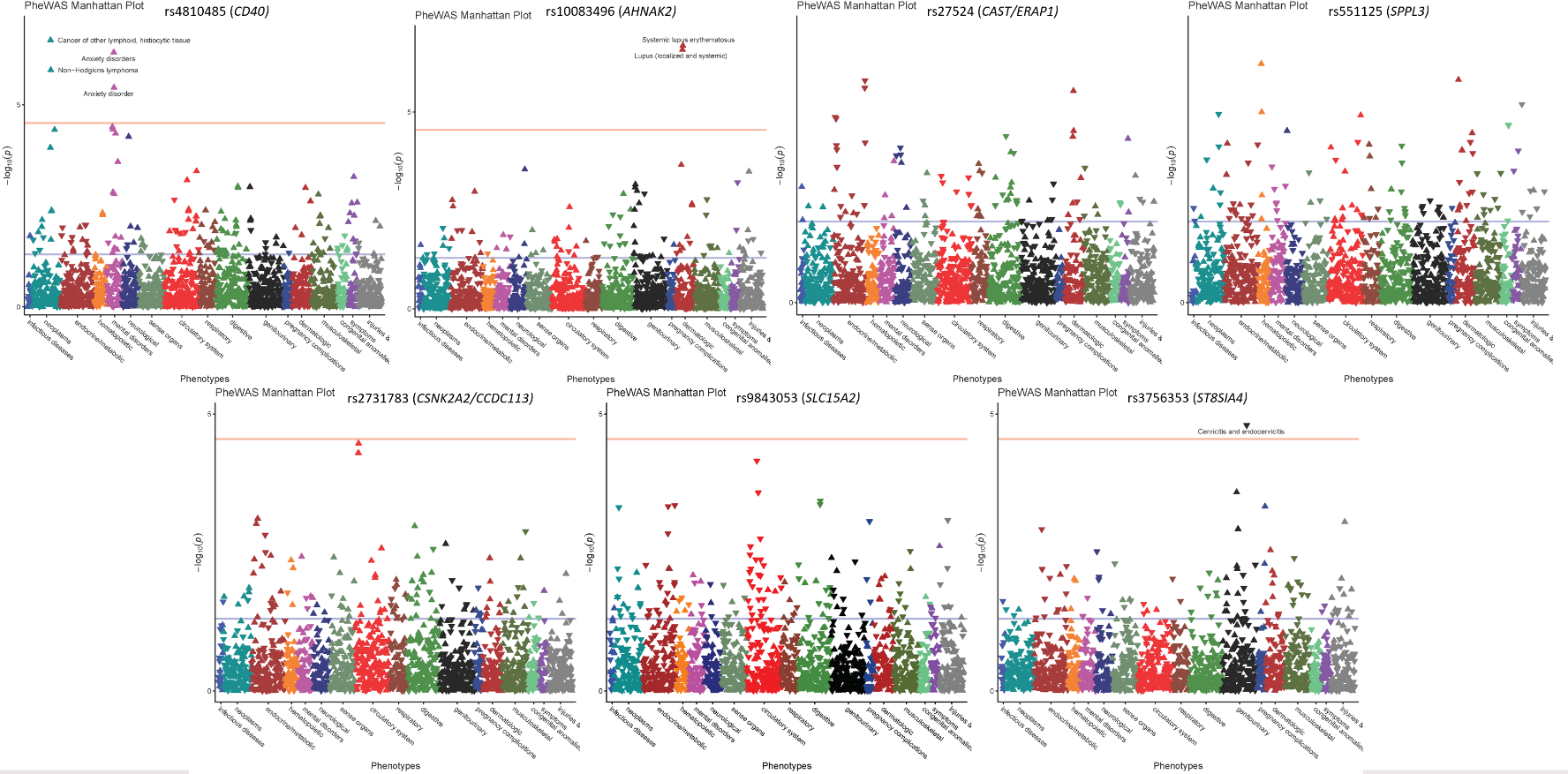
